# The Medical Impact of Emergent Banning of N-nitrosodimethylamine (NDMA)- contaminated Antihypertensive Drug: A nationwide longitudinal cohort study

**DOI:** 10.1101/2023.11.01.23297897

**Authors:** Juhee Ahn, Sungho Won, Jong-Heon Park, Daichi Shimbo, Hae-Young Lee

## Abstract

**OBJECTIVE:** To study the association of NDMA-contaminated generic valsartan with cancer incidence.

**DESIGN:** Nationwide longitudinal observational cohort study.

**SETTING:** Claims data from the National Health Insurance Service of South Korea was analyzed using 1:1:1 pairwise propensity score matching (PSM) of NDMA-uncontaminated original, NDMA-contaminated generic, and initially-suspended-but-finally confirmed as NDMA-uncontaminated generic valsartan user-groups. Time-dependent Cox models and dose-response evaluation were used to evaluate carcinogenicity.

**PARTICIPANTS:** A total of 3,231,212 participants with satisfactory minimal adherence, followed up from January 1, 2013 until December 31, 2020.

**INTERVENTIONS:** At least one tablet of valsartan.

**MAIN OUTCOME MEASURES:** The primary outcome was primary cancer incidence and secondary outcome was 12 prevalent organ-specific cancer incidences. All-cause and cardiovascular mortality risks were estimated before and after valsartan withdrawal.

**RESULTS:** Among participants (mean [standard deviation] age 59.5 [13.1] years; male, 53.5%), new users had adjusted hazard ratios and 95% confidence intervals (CI) of 1.069 (1.054 to 1.085) and 1.142 (1.100 to 1.186) for any cancer in the NDMA-exposed period (versus NDMA-unexposed) before and after 1:1:1 PSM, respectively. Regardless of PSM, lung and prostate cancer risks increased significantly during the NDMA-exposed period whereas, post-PSM, cancer risk did not increase in the eventually-NDMA-uncontaminated group, even in new users. All-cause and cardiovascular mortality did not differ significantly with NDMA exposure before and after emergent banning.

**CONCLUSIONS:** NDMA-contaminated valsartan increased cancer risk, especially of lung and prostate cancers. All-cause and cardiovascular mortality did not evidently increase following banning. This supports emergent health-policy action against potentially carcinogenic drugs.

## Introduction

Pharmacological treatment of non-communicable diseases requires long-term use. Therefore, it is existential threat that even a very small amount of carcinogen causes substantial problems through long-term accumulation. Hence, concerns about the possibility of cancer associated with antihypertensive drugs are continuously raised and resolved. Representative controversies have occurred with antihypertensive medications and overall cancers,^1^ angiotensin receptor blockers and lung cancer,^2^ nifedipine with breast and gastrointestinal tract cancers,^3–5^ and hydrochlorothiazide and skin cancer.^6^

Recent concerns include the potential carcinogenic effect of impurities generated during manufacturing processes rather than the main drug ingredient. The European Medicines Agency withdrew medicines containing the active substance, valsartan, which was supplied by Huahai Pharmaceuticals, China, from the market in July 2018 because of contamination with N-nitrosodimethylamine (NDMA).^7^ The United States Food and Drug Administration followed this action after confirming its contamination. On July 7, 2018, the Korea Ministry of Food and Drug Safety immediately suspended 219 generic valsartan products potentially contaminated with NDMA, and finally withdrew 115 generic valsartan products on July 9, 2018.^8^ Investigations reported that Huahai Pharmaceuticals probably brought the new manufacturing process contaminating NDMA around 2012.^9^ Shortly after the withdrawal action in 2018, Pottegård et al. reported no significant increase in overall cancer (adjusted hazard ratio [aHR] 1.09, 95% confidence interval [CI] 0.85-1.41) and no dose-response relation based on the Danish health registry during 4-year interval.^10^ Gomm et al., in 2021, based on German health insurance data, reported a statistically significant association between exposure to NDMA- contaminated valsartan and hepatic cancer (aHR 1.16; 95% CI 1.03-1.31); however, no association was found with the overall cancer risk during a 3-year interval.^7^ In contrast, observational study based on the French National Health Data System reported that the use of the NDMA-contaminated valsartan did not increase the overall risk of cancer, but suggested the increased risk of liver cancer and melanoma for a median follow-up of 4 years.^11^

In Korea, as an immediate administrative measure, >90% of related valsartan generic prescriptions were suspended after the government announcement, and most were automatically replaced with the original valsartan. Korea’s health policy mandates an immediate withdrawal from the market if drugs have potential risks before confirming the substantiality of the risks. However, there may be unintended consequences of a sudden suspension of antihypertensive drugs on to public health including a possibility that adherence to antihypertensive drugs might be reduced due to fear of cancer occurrence and overall distrust of medications; regarding cancer incidence, many health checkups might be conducted due to increased health concerns of patients with an increase in prostate, thyroid and breast cancer incidence, because they are frequently diagnosed during screenings; and a possibility that cardiovascular events might increase due to a decrease in adherence to antihypertensive medications.

Therefore, this study evaluated the association of NDMA-contaminated generic valsartan with cancer incidence and compared all-cause and cardiovascular deaths before and after valsartan withdrawal.

## Methods Data source

We conducted a long-term observation analysis using nationwide claims data from the National Health Insurance Service (NHIS)-National Health Information Database (NHID), Korea. The NHIS is a single, mandatory medical care system that covers approximately 50 million people residing in Korea. We also linked the Cause of Death Statistics by Korea Statistics to NHIS-NHID via the Microdata Integrated Service (http://mdis.kostat.go.kr). This study was exempted from review by the institutional review board (E-2103-057-1203) because of the anonymized data. Informed consent requirement was unattainable, because the NHIS- provided data were de-identified. The NHIS approved the use of the released data in 2022.

### Study population

We enrolled patients treated with at least one valsartan prescription between January 1, 2013 and December 31, 2020. Eligible patients had received a type of valsartan with prescription days ≥20% during a 3-month period and were aged ≥30 years. The index date was set to January 1, 2013, and patients were followed up until the date of death or December 31, 2020. We excluded patients with a history of cancer before the index date.

### Exposure

Valsartan was grouped into three by the NHIS as follows: NDMA-uncontaminated original valsartan (Original valsartan), NDMA-contaminated generic valsartan (NDMA-contaminated valsartan), and initially-suspended but finally confirmed as NDMA-uncontaminated generic valsartan (eventually NDMA-uncontaminated valsartan). In Korea, of the initially withdrawn 219 valsartan products on July 7, 2018, the policy cleared 104 generic valsartan products on July 9, 2018, and these were confirmed as ‘eventually NDMA-uncontaminated’ generic valsartan. However, patients prescribed with ‘eventually NDMA-uncontaminated’ generic valsartan received same recall messages; thus, they might show similar health behavior as the NDMA-contaminated valsartan group. Therefore, we included them in the analysis as an internal control group.

To avoid immortal time bias, valsartan was considered as a time-dependent exposure. The period from January, 2013 to the first observed valsartan prescription or follow-up among patients who received original valsartan was defined as the NDMA-unexposed period. The follow-up time of patients who received NDMA-contaminated valsartan was grouped into the NDMA-unexposed and NDMA-exposed periods. For the eventually NDMA-uncontaminated valsartan, patients had NDMA-unexposed and eventually NDMA-uncontaminated valsartan- exposed periods.

### Endpoints

The primary outcome was any primary cancer development. We also considered the incidence of 12 prevalent organ-specific cancers in Korea based on ICD-10 codes (Supplemental Table 1). The onset date was defined as the first diagnosis of any or organ-specific cancers. To evaluate the safety, all-cause and cardiovascular death were considered as secondary outcomes. We divided the year 2018 from January 1 to July 6 (before withdrawal) and July 7 to December 31 (after withdrawal) to compare the incidence of safety outcomes. The ICD10-codes of fatal cardiovascular events included hypertensive disease (I10-16), ischemic heart disease (I20-25), myocarditis/cardiomyopathy (I40-43), arrhythmias/heart failure (I46-52), cerebrovascular disease (I60-69), and aortic aneurysms (I70-73).

### Statistical analysis

To mitigate confounding effects, we performed 1:1 or 1:1:1 pairwise propensity score matching (PSM) with age, sex, Charlson’s comorbidity index (CCI), prevalent user of antihypertensive medication, drinking status, presence of disability, and level of healthcare provider from primary to tertiary hospitals (Supplemental Table 2). We compared participants’ baseline characteristics before and after PSM by using an analysis of variance (ANOVA) or Welch’s ANOVA for continuous variables and a χ^2^ test for categorical variables. Crude incidences and their 95% CIs of our primary and secondary outcomes were estimated under the assumption that each event follows the Poisson distribution. Time-dependent Cox models were used to estimate aHRs considering age, sex, CCI, prevalent user, and the year of the first observed valsartan prescription. The dose-response relationship was evaluated using the same model. The risks of all-cause and cardiovascular deaths were estimated in the same manner. We replicated these analyses in valsartan new users. We also performed same analysis after excluding patients with events within 6 months, 1 year, and 2 years after the first observed valsartan prescription; then, the best model was selected using Akaike information criterion (AIC). And, three sensitivity analyses were implemented: Subgroup analyses in lung cancer toward sex and smoking status, and prostate cancer towards ≤65 or > >65 years old; Patients with any cancer during the first year of follow-up were excluded; Patients with receiving valsartan prescriptions since 2015.

The significance level for all statistical analyses was set at 0.05, and *P* <.05 was considered significant. Statistical analysis was performed using SAS Enterprise Guide (version 7.13; SAS Institute), R software (version 4.1.3; R project), and Rex (version 3.5.3; RexSoft).

## Results

### Baseline Characteristics

Original and NDMA-contaminated generic valsartan were used from the study onset (January 1, 2013), while the eventually NDMA-uncontaminated valsartan was first claimed in September 2013. All prescriptions of valsartan invariably decreased after July, 2018, while those for NDMA-contaminated valsartan were totally eliminated. For the eventually NDMA- uncontaminated valsartan, although the temporary withdrawal was dismissed two days after initial suspension, the claim amount decreased in July–October, 2018, but maintained at the end of study period (December 31, 2020) (Supplemental Figure 1).

We identified 3,231,212 participants who received at least one tablet of valsartan, satisfying minimal adherence to prescription. Table 1 shows the baseline characteristics of the entire cohort and by valsartan groups. Compared with the original valsartan group, the NDMA- contaminated and eventually NDMA-uncontaminated generic valsartan groups reported hypertensive medication history more. In contrast, patients in the original valsartan group had higher comorbidity (high CCI) and were more likely to be treated in secondary and tertiary hospitals.

**Table 1.** Baseline characteristics of study population before propensity score matching.

| Characteristics | Total<br>(*n*=3,231,212) | Original valsartan<br>(*n* = 2,340,729) | Eventually NDMA-uncontaminated<br>valsartan<br>(*n* = 392,604) | NDMA-contaminated<br>valsartan<br>(*n* = 497,879) | *P* |
| --- | --- | --- | --- | --- | --- |
| *Age, years — Mean (SD)* | 59.5 (13.1) | 59.6 (13.1) | 58.4 (13.0) | 60.2 (13.0) | < 0.001<sup>‡</sup> |
| *Sex — n (%)* |  |  |  |  |  |
| Male | 1,729,185 (53.5) | 1,251,572 (53.5) | 220,216 (56.1) | 257,397 (51.7) | < 0.001 |
| Female | 1,502,027 (46.5) | 1,089,157 (46.5) | 172,388 (43.9) | 240,482 (48.3) |  |
| *CCI — n (%)* |  |  |  |  | < 0.001 |
| 0 | 1,117,080 (34.6) | 795,896 (34.0) | 152,286 (38.8) | 168,898 (33.9) |  |
| 1 | 811,326 (25.1) | 585,361 (25.0) | 98,847 (25.2) | 127,118 (25.5) |  |
| 2 | 545,323 (16.9) | 396,944 (17.0) | 62,529 (15.9) | 85,850 (17.2) |  |
| ≥ 3 | 757,483 (23.4) | 562,528 (24.0) | 78,942 (20.1) | 116,013 (23.2) |  |
| *Being a prevalent user — n (%)* |  |  |  |  | < 0.001 |
| No | 2,580,766 (79.9) | 1,726,616 (73.8) | 392,548 (99.9) | 461,599 (92.7) |  |
| Yes | 650,446 (20.1) | 614,110 (26.2) | 56 (0.1) | 36,280 (7.3) |  |
| *Drinking status — n (%)* |  |  |  |  | < 0.001<sup>§</sup> |
| No | 580,727 (18.0) | 423,135 (18.1) | 64,843 (16.5) | 92,749 (18.6) |  |
| Yes | 525,191 (16.3) | 377,406 (16.1) | 67,279 (17.1) | 80,506 (16.2) |  |
| Missing | 2,125,294 (65.8) | 1,540,188 (65.8) | 260,482 (66.4) | 324,624 (65.2) |  |
| *Smoking status — n (%)* |  |  |  |  | < 0.001 |
| Never | 633,278 (19.6) | 458,070 (19.6) | 72,978 (18.6) | 102,230 (20.5) |  |
| Ever | 214,149 (6.6) | 158,567 (6.8) | 24,657 (6.3) | 30,925 (6.2) |  |
| Current | 258,198 (8.0) | 183,691 (7.9) | 34,437 (8.8) | 40,070 (8.1) |  |
| Missing | 2,125,587 (65.8) | 1,540,401 (65.8) | 260,532 (66.4) | 324,654 (65.2) |  |
| **Disability — n (%)** |  |  |  |  | < 0.001 |
| No | 366,137 (11.3) | 266,799 (11.4) | 41,883 (10.7) | 57,455 (11.5) |  |
| Yes | 2,865,075 (88.7) | 2,073,930 (88.6) | 350,721 (89.3) | 440,424 (88.5) |  |
| **Level of healthcare provider — n (%)** |  |  |  |  | < 0.001<sup>§</sup> |
| Tertiary | 275,395 (8.5) | 272,014 (11.6) | 680 (0.2) | 2,701 (0.5) |  |
| Secondary | 777,139 (24.1) | 636,297 (27.2) | 59,218 (15.1) | 81,624 (16.4) |  |
| Primary | 1,966,594 (60.9) | 1,313,803 (56.1) | 297,279 (75.7) | 355,512 (71.4) |  |
| Healthcare center | 185,545 (5.7) | 100,059 (4.3) | 31,432 (8.0) | 54,063 (10.9) |  |
| Others<sup>†</sup> | 24,665 (0.8) | 17,128 (0.7) | 3,749 (1.0) | 3,788 (0.8) |  |
| Missing | 1,874 (0.1) | 1,428 (0.1) | 255 (0.1) | 191 (0.1) |  |
| **Follow-up duration, year - median (IQR)** |  | 8.0 (8.0-8.0) | 3.4 (1.8-5.1) | 4.3 (3.0-5.9) | < 0.001 |

Abbreviations: CCI, Charlson's comorbidity index; IQR, interquartile range; NDMA, N-nitrosodimethylamine; SD, standard deviation
<sup>†</sup>Others' healthcare providers included dentistry, psychiatry, and oriental medicine.
<sup>‡</sup>Welch's *t*-test was performed for unequal population variances.
<sup>§</sup>Chi-square test was performed including missing value.

All covariates before PSM differed according to the exposure (valsartan). Therefore, we performed PSM for the comparison between NDMA-uncontaminated original valsartan and NDMA-contaminated generic valsartan groups (Supplemental Table 2). After 1:1 PSM, 167,042 of NDMA-contaminated and 167,042 of original valsartan users were identified (Supplemental Table 3). Next, we performed 1:1:1 PSM for the comparison of the three groups (Supplemental Table 4). After 1:1:1 pairwise-PSM, 127,053 NDMA-contaminated, 127,053 eventually NDMA-uncontaminated, and 127,053 of original valsartan users were identified. The baseline characteristics between these exposure groups were moderated after PSM (Supplemental Tables 3 and 4). The flow diagram of the study is presented in Figure 1.

**Figure 1.**
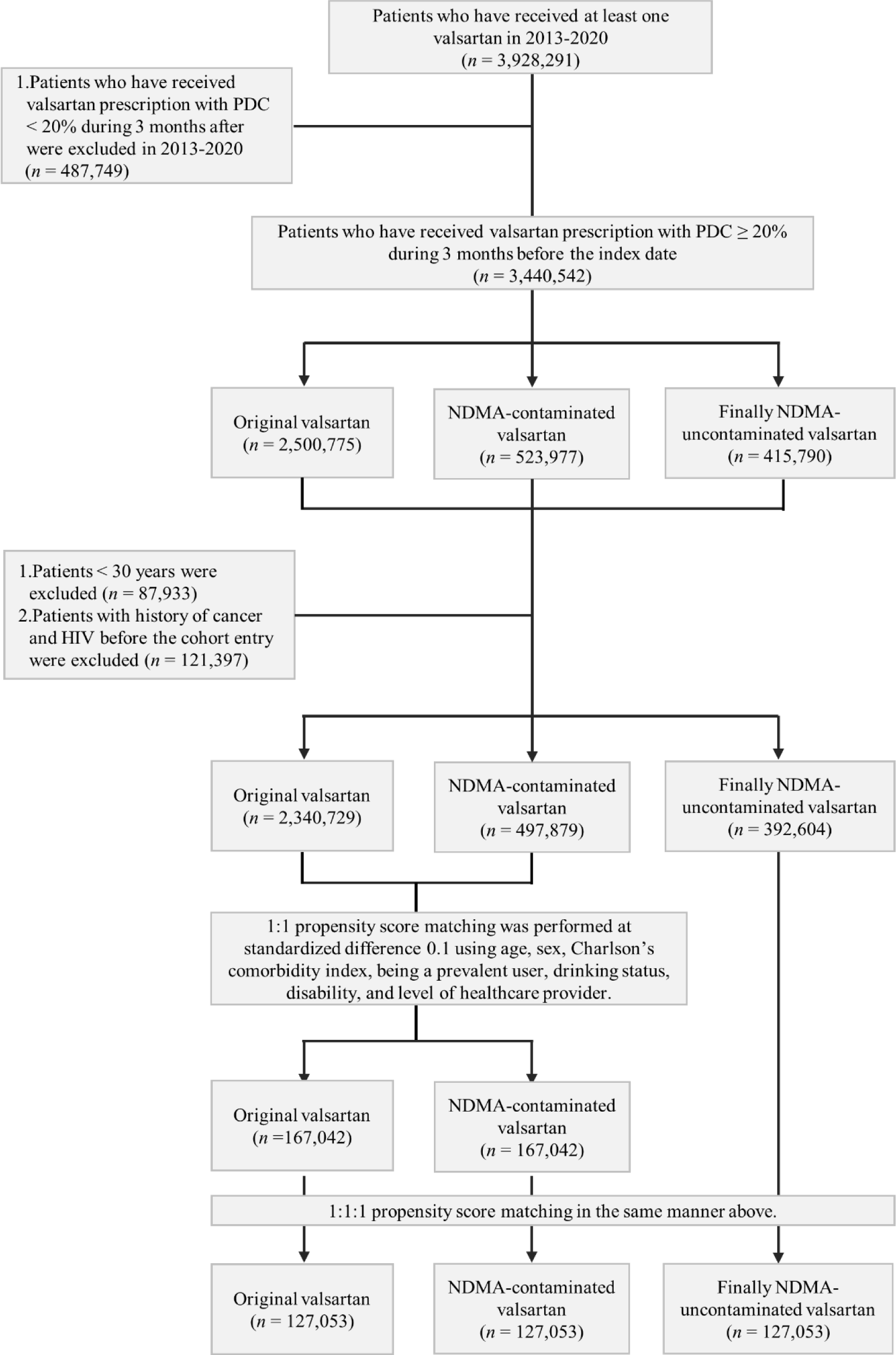
Flowchart of this study. HIV, human immunodeficiency virus; NDMA, N- Nitrosodimethylamine; PDC, prescription of days covered.

Before PSM, the mean follow-up years (standard deviation [SD]) of each exposure group were 7.2 (1.9), 7.1 (1.7), and 7.2 (1.6) for the original, NDMA- contaminated, and eventually NDMA-uncontaminated valsartan groups, respectively. The median follow-up years (interquartile range) according to exposure groups were shown in for study population before PSM (Table 1) and after PSM (Supplemental Tables 3 and 4).

### Incidence of Primary Cancers

The crude incidences rate (case per 1,000 person-years) and their 95% CIs of any cancer before and after PSM were 21.54 (21.47-21.61) and 19.87 (19.59-20.16) for original valsartan; 19.77 (19.62-19.91) and 19.61 (19.33-19.89) for NDMA-contaminated valsartan; 17.49 (17.35-17.65) and 18.27 (18.00-18.54) for eventually NDMA-uncontaminated valsartan, respectively (Supplemental Table 5).

The aHRs of any and 12 organ-specific cancers in original population were estimated according to different lag criteria (6 months and 1 and 2 years) (Supplemental Table 6). For each clinical outcome, the lowest AICs were detected when participants who experienced events within 2 years were excluded; therefore, we adopted it to all results. The risks (95% CI) of any cancer for the NDMA-exposed period (compared to those for the NDMA-unexposed) in new user population, were 1.098 (95% CI 1.084-1.113) for before PSM; 1.120 (95% CI 1.088-1.153) for 1:1 PSM; and 1.142 (95% CI 1.100-1.186) for 1:1:1 PSM (Supplemental Table 6). Those had the same trend even in new user population and were 1.069 (95% CI 1.054-1.085) for before PSM, 1.119 (95% CI 1.084-1.155) for 1:1 PSM, and 1.142 (1.100-1.186) for 1:1:1 PSM (Figure 2 and Supplemental Table 7). However, the risks (95% CI) of any cancer of the eventually NDMA-unexposed period (compared to those for the NDMA-unexposed) were also increased, being biologically implausible.

**Figure 2.**
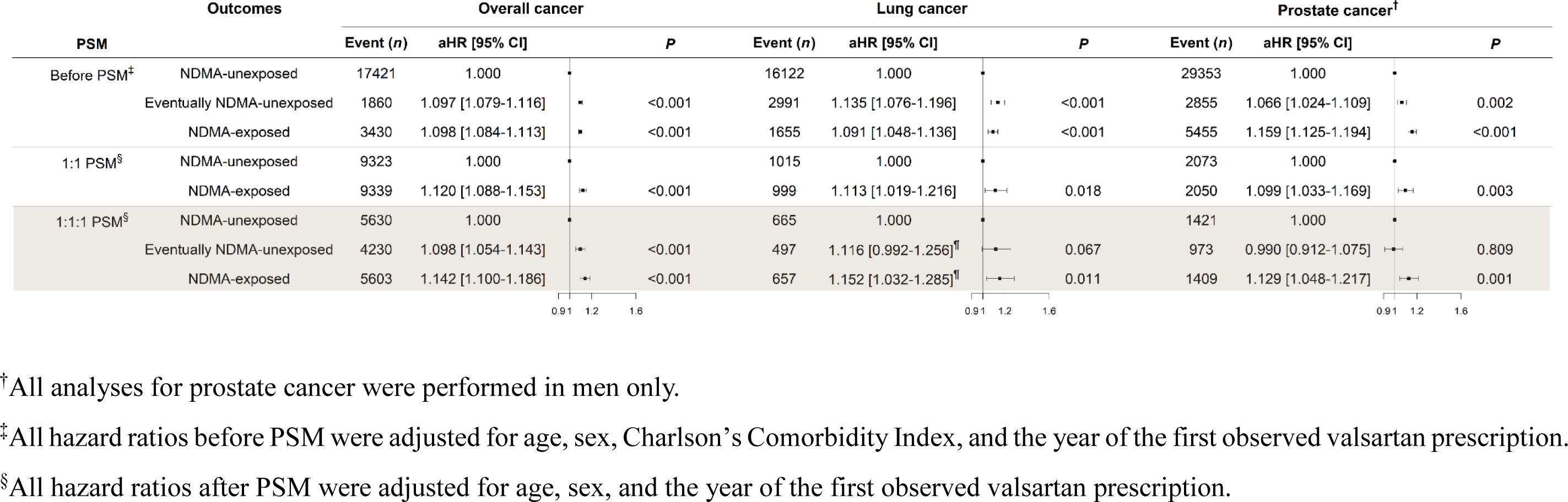
The risks of overall, prostate, and lung cancers in new user population before and after propensity score matching. All the results were estimated excluding patients with the events within 2 years after the first observed valsartan prescription. aHR, adjusted hazard ratio; CI, confidence interval; NDMA, N-Nitrosodimethylamine; PSM, propensity score matching.

However, in the analyses of 12 individual cancer outcomes, lung and prostate cancers showed significantly increased risk in the NDMA-exposed period regardless of PSM, but no increased risk in the eventually NDMA-uncontaminated group after PSM (new user population in Figure2 and Supplemental Table 7; original population in Supplemental Table 6). Colorectal and stomach cancers also showed a significantly increased risk for NDMA-exposed period regardless of PSM. However, both cancers showed an increased risk in the eventually NDMA- uncontaminated group after 1:1:1 PSM, suggesting the false-positive association (Supplemental Table 6).

Next, we evaluated the dose-response relation in original population (Supplemental Table 8). The aHRs of nine organ-specific cancers were significant, but only three organ-specific cancers (lung, prostate, and uterine cancers) showed dose-response relationships before PSM. However, after both PSMs, only lung and prostate cancers showed significantly higher risk in the highest dose. We found similar dose-response trend of overall, lung, and prostate cancers in new user population (Figure 3 and Supplemental Table 9).

**Figure 3.**
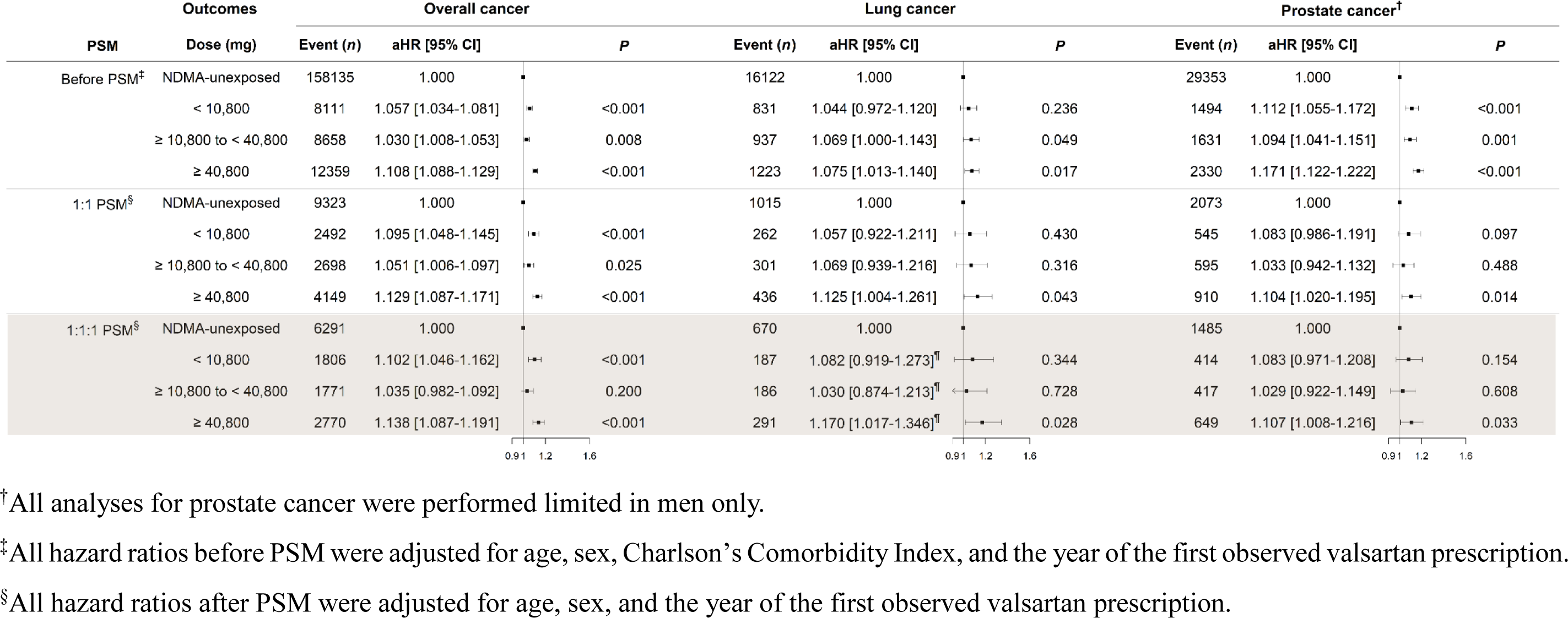
The risks of overall, prostate, and lung cancers in new user population before and after propensity score matching according to NDMA-contaminated valsartan doses. All the results were estimated excluding patients with the events within 2 years after the first observed valsartan prescription. aHR, adjusted hazard ratio; CI, confidence interval; NDMA, N-Nitrosodimethylamine; PSM, propensity score matching.

Then, we made subgroup analyses in lung and prostate cancers as sensitivity analysis. In lung cancer, the increased trend of aHR with NDMA-exposed period were consistent regardless sex or smoking status (Supplemental Table 10). In prostate cancer, the trend was also consistent regardless of age group (Supplemental Table 11). In another sensitivity analysis that excluded patients with any cancer during the first year of follow-up, the results were similar both in overall population and new user only (Supplemental Tables 12 and 13), however, dose-response relation disappeared in prostate cancer (Supplemental Tables 14 and 15). When restriction to patients receiving valsartan since 2015, the clinical significance only remained in any cancer although the trend were similar, which were mainly due to reduced sample numbers (Supplemental Tables 16, 17, 18, and 19).

### Risks of All-cause and Cardiovascular Death Before and After Immediate Banning

The crude all-cause and cardiovascular death rates (case per 1,000 person-years) in the entire study population and their 95% CIs before and after immediate banning were evaluated (Supplemental Table 20). There was no evidence of significantly different risks of all-cause and cardiovascular deaths before and after emergent banning (July 7, 2018) according to exposure to NDMA in the whole study population (*P* = 0.225 for all-cause and *P* = 0.418 for cardiovascular death; Table 2). After 1:1:1 pairwise PSM, the results were also similar (*P* = 0.683 and *P* ≥ 0.999, respectively; Table 2).

**Table 2.** Comparison of risks of all-cause and cardiovascular deaths before and after July 7, 2018 in South Korea.

| PSM |  | Death | All-cause |  |  | Cardiovascular |  |  |
| --- | --- | --- | --- | --- | --- | --- | --- | --- |
|  |  |  | Df | Wald Chi-square | P | Df | Wald Chi-square | P |
| Before | $H_0: \beta_{\text{NDMA-contaminated} \mid \text{Jan 1-Jul 6}} = \beta_{\text{NDMA-contaminated} \mid \text{Jul 7-Dec 31}} \text{ \& } \beta_{\text{Eventually NDMA-uncontaminated} \mid \text{Jan 1-Jul 6}} = \beta_{\text{Eventually NDMA-uncontaminated} \mid \text{Jul 7-Dec 31}}$ | | 2 | 2.9850 | 0.225 | 2 | 1.7454 | 0.418 |
|  | Contrasts (Jan 1-Jul 6 vs Jul 7-Dec 31) |  |  |  |  |  |  |  |
| | $H_0: \beta_{\text{NDMA-contaminated} \mid \text{Jan 1-Jul 6}} = \beta_{\text{NDMA-contaminated} \mid \text{Jul 7-Dec 31}}$ | | 1 | 2.2979 | 0.130 | 1 | 0.2951 | 0.587 |
| | $H_0: \beta_{\text{Eventually NDMA-uncontaminated} \mid \text{Jan 1-Jul 6}} = \beta_{\text{Eventually NDMA-uncontaminated} \mid \text{Jul 7-Dec 31}}$ | | 1 | 0.7082 | 0.400 | 1 | 1.4440 | 0.230 |
| After 1:1:1 | $H_0: \beta_{\text{NDMA-contaminated} \mid \text{Jan 1-Jul 6}} = \beta_{\text{NDMA-contaminated} \mid \text{Jul 7-Dec 31}} \text{ \& } \beta_{\text{Eventually NDMA-uncontaminated} \mid \text{Jan 1-Jul 6}} = \beta_{\text{Eventually NDMA-uncontaminated} \mid \text{Jul 7-Dec 31}}$ | | 2 | 0.7620 | 0.683 | 2 | 0.0001 | $\geq 0.999$ |
|  | Contrasts (Jan 1-Jul 6 vs Jul 7-Dec 31) |  |  |  |  |  |  |  |
| | $H_0: \beta_{\text{NDMA-contaminated} \mid \text{Jan 1-Jul 6}} = \beta_{\text{NDMA-contaminated} \mid \text{Jul 7-Dec 31}}$ | | 1 | 0.4121 | 0.521 | 1 | 0.0001 | 0.992 |
| | $H_0: \beta_{\text{Eventually NDMA-uncontaminated} \mid \text{Jan 1-Jul 6}} = \beta_{\text{Eventually NDMA-uncontaminated} \mid \text{Jul 7-Dec 31}}$ | | 1 | 0.2656 | 0.606 | 1 | 0.0000 | 0.996 |

Abbreviations: Jan, January; Df, degree of freedom; Dec, December; Jul, July; NDMA, N-Nitrosodimethylamine; PSM, propensity score matching.

## Discussion

In this nationwide cohort study, we observed an increased risk of any cancer associated with the use of NDMA-contaminated valsartan products. In the analysis of 12 prevalent cancer outcomes, lung and prostate cancers showed significantly increased risk in the NDMA- exposure period regardless of PSM and no increased risk in the eventually NDMA- uncontaminated group after PSM. In addition, lung and prostate cancers showed significantly higher risk in the highest dose before and after PSM. In contrast, there was no evidence that the risks of all-cause and cardiovascular deaths were increased and after emergent banning. Although NDMA-contaminated valsartan is no longer on the market, the issue of impurities in antihypertensive drugs continues, including recent azide (azidomethyl-biphenyl-tetrazole) issue associated with the angiotensin type 1 receptor antagonist class, namely “sartan’,^12^ and the results of this study can offer valuable evidence for determining future health policy stance.

The principal strength of this study was the use of a high-quality nationwide registry, without selection bias. Because the NHIS in Korea is a single, mandatory medical care system, this study enabled the collection of >3 million patients’ data for >7 years. Additionally, as confirmed by the Korea MFDS, we could provide the three valsartan categories from the NHIS. In particular, patients prescribed with ‘eventually NDMA-uncontaminated’ generic valsartan had a high possibility of the same reaction as the NDMA-contaminated valsartan group. In principle, the Korean NHIS-NHID claim data do not allow the search for product names (only generic names can be searched) to prevent commercial use. However, with the 3-year’s efforts (2020 to 2022), the Korean NHIC accepted that it was a study for public purposes, and offered the data classified into three groups, not according to the individual brand names. Therefore, we minimized possible bias in four ways; by increasing the lag period after exposure, performing PSM, establishing dose-response relationship, and identifying no significant association in the eventually NDMA-uncontaminated valsartan group as an internal control. Lastly, in Korea, the National Plan for Cancer Control was implemented in 1996 and all cancer patients were benefited of reduced insurance rate regarding cancer treatment, which resulted in the near complete and correct registration (98.3%) of all cancers in the Korea National Cancer Incidence Database.^13^

The International Agency for Research on Cancer classified NDMA as “probably carcinogenic to humans” owing to limited evidence of carcinogenicity in humans and sufficient evidence of carcinogenicity in animal studies.^14^ NDMA is suspected to exert both localized and systemic carcinogenic effects due to the induction of DNA-damaging metabolites in the liver and gastrointestinal tract.^15^ Furthermore, tumors in the gastrointestinal tract, lungs, and liver have been observed in animal studies.^16^ Regarding individual cancers, we reported that the lung and prostate cancers satisfactorily fulfilled all the criteria. Gastrointestinal tract tumors and uterine cancer (but not prostate cancer) showed possible association, which were all observed in animal experiment. Although the association between prostate cancer and NDMA has not been reported previously, it is possible that urinary excretion of the carcinogenic substances cause prostate cancer.^17^ According to an animal experimental study, if patients with a body weight of 70 kg and an average body surface area take the highest dose of valsartan (320 mg/day) containing the highest observed level of NDMA (20.19 μg) over a 6-year period, 126 cases of cancer per 100,000 people might occur.^18^ which is about double our overall estimation. The European Medicines Agency estimated that there could be one extra case of cancer for every 5,000 patients exposed to contaminated valsartan at the highest dose.^19^ Therefore, it is not surprising that the patients with NDMA-contaminated valsartan group showed higher cancer incidence in this large-number, long term follow-up data.

The results of this study might be compared with those of the French group in many aspects.^11^ The subjects of this study were about 5 years younger than those in the French study and had a higher proportion of patients without a history of previous hypertension medication use. However, there is a possibility that the proportion of high-risk patients (23% of Charlson’s comorbidity index ≥ 3) could be higher, and the rates of alcohol consumption and smoking were also higher compared with those enrolled in French study. In the case of the study population, the number of subjects in this study was 3.2 million, twice the number of the French study’s 1.5 million. However, the number of patients in the NDMA-contaminated valsartan group was more than twice in the French group. This difference is attributed to the relatively small price difference between the off-patent original valsartan drug and the generic valsartan in Korea, leading to a higher market share for the original valsartan.^20^ Regarding the duration of follow-up observation, this study was slightly longer, but still not considerably long. However, considering the distribution period from the market release to the banning of NDMA- contaminated drugs, it can be considered as the maximum duration possible. In the French study, there was no difference in the overall cancer risk, whereas in this study, the overall cancer risk was 1.142 times higher (Supplemental table 6). There might be several explanations. First, the overall cancer incidence rate, more exactly detection rate, in this study (19.8-21.5 per 1000 people) was higher than that in the French study (12.93-13.09 per 1000 people) (Supplemental table 5). This higher detection rate in Korea might be the Korean Cancer Registration Act, increasing insurance coverage regarding cancer treatment, which resulted in the near complete and correct registration (98.3%) of all cancers in the Korea National Cancer Incidence Database.^13^ Therefore it is possible that these differences in early detection rates might detect significant risk that could not be found by analysis of small numbered, more symptomatic patients during a limited follow-up period. Secondly, in the 1:1:1 PSM comparison in this study, an approximately 12% higher incidence rate was also observed in the eventually NDMA- uncontaminated valsartan group (Supplemental table 6). Although NDMA-contaminated valsartan group showed significantly higher risk compared with the eventually NDMA- uncontaminated valsartan group, some false positive risk might be included in the higher cancer risk in the NDMA-contaminated valsartan group. Indeed, similar finding of abrupt increase in reporting of neoplasms associated with valsartan after medication recall regardless of NDMA contamination was reported.^21^ However, our data showed that the overall cancer risk is still significantly higher even considering the possible false-positive rate. Regarding individual cancers, the French study showed a higher risk of liver cancer and melanoma, but this study did not show a significant increase of them. Melanoma is the most common tumor in Western countries, but its frequency is very low in East-Asian countries,^22^ hence it was not analyzed in this study’s evaluation of the top 12 prevalent cancers in Korea. On the other hand, in the case of liver cancer, the majority causes are hepatis B (74%) and C (9%) viral related, while toxin-related cases are very rare in Korea.^23^ If NDMA acted as a carcinogenesis enhancer, there could make a significantly higher risk in the prevalent liver cancer in Korea. However, this study did not find such an effect, suggesting that NDMA is more likely to function as a direct carcinogen rather than a carcinogenesis enhancing function. In contrast, this study found a significant occurrence of lung and prostate cancers, while the French study did not show a difference. While there is no clear reason for this, such as ethnic/social differences in the cases of melanoma and liver cancer, the higher incidence (early detection) rates of these two tumors might contribute to the significant differences, otherwise not detected analysis of symptomatic patients during the limited 4-year follow-up period cases. Despite the several differences between two studies, we think that both studies add more value when together than alone in that they have confirmed that cancer issue with NDMA-contaminated valsartan is not a hypothetical threat, but is actually likely to increase cancer incidence.

When the results of clinical or basic research are applied to the real-world, it is not uncommon to observe an adverse effect from well-intentioned health behavior. One famous example in the cardiology field is the RALES study-related issue. After the beneficial effect of spironolactone, a potassium-sparing diuretic in heart failure was published in 1999, the prescription of the drug has exploded, however resulted in increased emergency room visits and deaths due to adverse events of hyperkalemia.^24^ After an alert by the European Medicines Agency and an urgent banning announcement by Korean MFDS, there were many patients’ complaints and anxieties. Physicians raised concerns that adherence to antihypertensive drugs might be reduced by immaterial cancer risk, resulting in increased cardiovascular events. Indeed, in Canada, immediate increase in emergency department visits for hypertension and a delayed increase and hospitalizations for stroke/transient ischemic attack following the recall.^25^ However, the findings of this study did not show an increase in all-cause or cardiovascular deaths in the NDMA-contaminated or eventually NDMA-uncontaminated valsartan group, among whom adherence to cardiovascular medication might be reduced due to fear and distrust.

The current study has several limitations. Firstly, in real-world research, there are several confounding factors, which makes it difficult to draw a definitive conclusion beyond hypotheses generation. However, research evaluating the impact of carcinogens, including medications for chronic non-communicable diseases, is virtually impossible to conduct randomized clinical studies for ethical reasons as well as the very small incidence of cancer, demanding long-term follow-up of many patients. Therefore, it might an inherent limitation that only observational studies such as this study can be conducted. Above all, because regulations on drugs containing NDMA have become stricter and the possibility of marketability in the future is further reduced, research on the clinical significance of NDMA contaminants in the antihypertensive drug might be more difficult in the future. The unique advantage of this study is the presence of patients treated with ‘eventually NDMA- uncontaminated’ generic valsartan. Although the overall duration of the suspension of this group was only 3 days from July 7 to July 9, 2018, the claim amount was continuously decreased, which raised the possibility that they showed the same reaction as the NDMA- contaminated valsartan group. Therefore, we evaluated the incidence of cancer, cardiovascular death and overall death in this group as an internal control group, similar to the ‘placebo’ group in clinical trials. We believe that the existence of this group could compensate for the many limitations of observational research. Secondly, incidental cancer diagnosis might have increased due to increased health examinations because of health concerns of patients. However, we could not estimate the number of health examinations in each group because health examinations were not reimbursed by the NHIS, thus records were unavailable in the claim data. However, the incidence of thyroid or breast cancer was not significantly increased in NDMA-contaminated valsartan group, reducing the possibility of bias. Finally, we had hoped to evaluate whether the incidence of cardiovascular events increased due to a decrease in drug adherence. However, we only evaluated the incidence of cardiovascular death and all- cause deaths, and other soft endpoints, such as angina, coronary revascularization, nonfatal myocardial infarction, or heart failure hospitalization were not evaluated. This was because the diagnostic accuracy of these soft endpoints based on the claim data only was very low. However, we could not exclude the possibility that the nonfatal cardiovascular event was actually increased due to decreased adherence to medical treatment.

In conclusion, the use of valsartan products contaminated with NDMA was associated with increased risk of any cancer, especially lung and prostate cancers. However, there was no evidence that all-cause and cardiovascular deaths increased following emergent banning of possible NDMA-contaminated valsartan products. These results support the health policy of the emergent action to potentially hazardous drugs.

## Acknowledgments

This study used data from the National Health Information Database of the NHIS [Research administration No. NHIS-2021-1-774], and the results of the study are not related to the NHIS. We would like to thank Editage for the English language editing.

## Author’s Contributions

LHY conceived and designed the study, had full access to all of the data in the study and take responsibility for the integrity of the data and the accuracy of the data analysis. LHY, AJ, and WS drafted the paper. WS and AJ did the analysis and PJH contributed to data collection and interpretation.

## Transparency Statement

The lead author (HY Lee) affirms that the manuscript is an honest, accurate, and transparent account of the study being reported and no important aspects of the study have been omitted

## Ethical Approval

This study was waived from review by the institutional review board of Seoul National University Hospital (E-2103-057-1203) because of the anonymized data. Informed consent requirement was unattainable, because the NHIS-provided data were de-identified. The NHIS approved the use of the released data in 2022.

## Data availability statement

The entire analysis process of the linked data sources was conducted in the NHIS data analysis office. External exportation of the data and additional extraction of personal information were not permitted. The authors are restricted from sharing the data underlying this study because the Korean NHIS owns the data.

