## Supplemental Figure 1, Supplemental Table 1-20 for "The Medical Impact of Emergent Banning of N-nitrosodimethylamine (NDMA)- contaminated Antihypertensive Drug: A nationwide longitudinal cohort study"

**Supplemental Figure 1. Monthly prescription trend of valsartan in 2013-2020.** The red line presents the potentially estimated NDMA-contaminated valsartan, according to the Ministry of Food and Drug Safety in Korea. Lines indicate the number of tablets while color bars indicate the proportion of each group, among all valsartan prescriptions. NDMA, N-Nitrosodimethylamine.

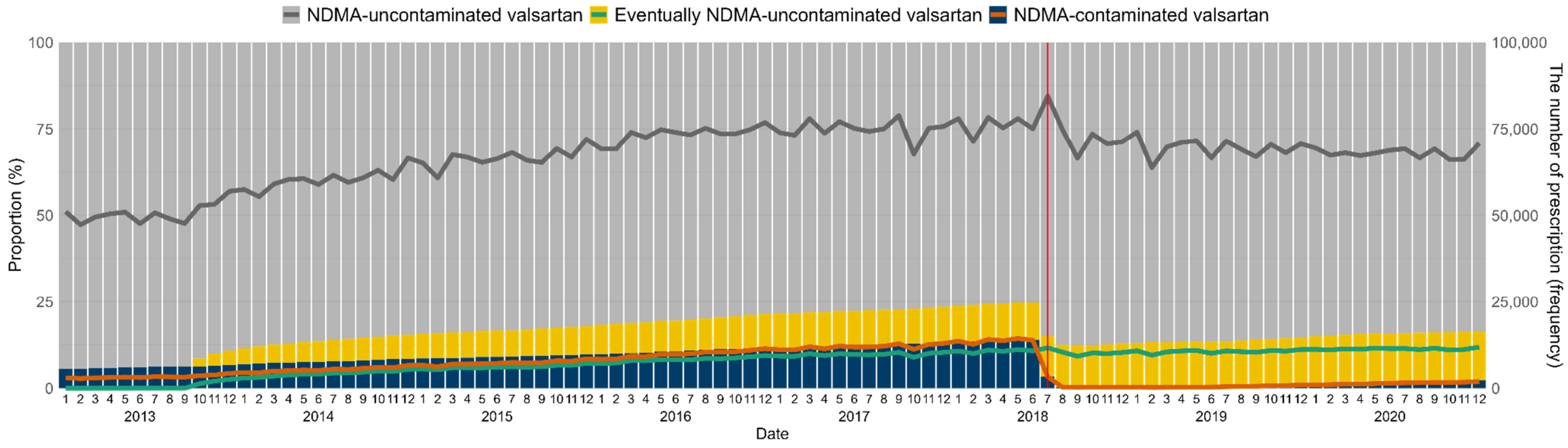

**Supplemental Table 1. Definition of cancers according to ICD-10 codes.**

| Code | Definition |
|---|---|
| C00 - C96 | Any cancer |
| C67 | Bladder |
| C50 | Breast |
| C18, C19, C20 | Colorectal |
| C23 ,C24 | Gallbladder and etc. |
| C64 | Kidney |
| C22 | Liver |
| C33, C34 | Lung |
| C25 | Pancreatic |
| C61 | Prostate |
| C16 | Stomach |
| C73 | Thyroid |
| C54, C55 | Uterine |

Abbreviation: ICD-10, International classification of diseases, tenth revision.

**Supplemental Table 2. Standardized mean differences after propensity score matching.**

| | **1:1 matching** | **1:1:1 matching** | | |
|---|---|---|---|---|
| | NDMA-contaminated valsartan vs original valsartan | Eventually NDMA-uncontaminated valsartan vs original valsartan | NDMA-contaminated valsartan vs original valsartan | Eventually NDMA-uncontaminated valsartan vs NDMA-contaminated valsartan |
| Age | 0.001 | -0.013 | 0.001 | 0.013 |
| Sex | 0.000 | 0.000 | 0.000 | 0.000 |
| CCI | 0.000 | 0.029 | 0.000 | 0.029 |
| Being a prevalent user | 0.001 | 0.000 | 0.000 | 0.017 |
| Drinking status | -0.001 | 0.005 | 0.000 | -0.005 |
| Disability | 0.001 | 0.005 | -0.001 | -0.006 |
| Level of healthcare provider | 0.000 | 0.036 | 0.000 | 0.036 |

Abbreviations: CCI, Charlson's comorbidity index; NDMA, N-Nitrosodimethylamine.

**Supplemental Table 3. Baseline characteristics of study population after 1:1 propensity score matching.**

| Characteristics | Total (*n* = 334,084) | Original valsartan (*n* = 167,042) | NDMA-contained valsartan (*n* = 167,042) | *P* |
|---|---|---|---|---|
| ***Age, years — Mean (SD)*** | 59.1 (11.8) | 59.1 (11.8) | 59.1 (11.8) | 0.740 |
| ***Sex — n (%)*** | | | | 1.000 |
| Male | 186,168 (55.7) | 93,084 (55.7) | 93,084 (55.7) | |
| Female | 147,916 (44.3) | 73,958 (44.3) | 73,958 (44.3) | |
| ***CCI*** | | | | 0.999 |
| *0* | 105,149 (31.5) | 52,581 (31.5) | 52,568 (31.5) | |
| *1* | 91,539 (27.4) | 45,766 (27.4) | 45,773 (27.4) | |
| *2* | 61,339 (18.4) | 30,659 (18.4) | 30,680 (18.4) | |
| ≥ *3* | 76,057 (22.8) | 38,036 (22.8) | 38,021 (22.8) | |
| ***Being a prevalent user — n (%)*** | | | | 0.695 |
| No | 308,792 (92.4) | 154,426 (92.5) | 154,366 (92.4) | |
| Yes | 25,292 (7.6) | 12,616 (7.6) | 12,676 (7.59) | |
| ***Drinking status — n (%)*** | | | | 0.865 |
| No | 178,783 (53.5) | 89,367 (53.5) | 89,416 (53.5) | |
| Yes | 155,301 (49.5) | 77,675 (46.5) | 77,626 (46.5) | |
| ***Smoking status — n (%)*** | | | | <0.001 |
| Never | 197,116 (59.0) | 98,647 (59.1) | 98,469 (59.0) | |

| | | | | |
|---|---|---|---|---|
| Ever | 61,106 (18.3) | 31,386 (18.8) | 29,720 (17.8) | |
| Current | 75,735 (22.7) | 36,937 (22.1) | 38,798 (23.2) | |
| Missing | 127 (0.04) | 72 (0.04) | 55 (0.03) | |
| ***Disability — n (%)*** | | | | 0.744 |
| No | 35,116 (10.5) | 149,513 (89.5) | 149,455 (89.5) | |
| Yes | 298,968 (89.5) | 17,529 (10.5) | 17,587 (10.5) | |
| ***Level of healthcare provider — n (%)*** | | | | 0.998 |
| Tertiary | 1,844 (0.6) | 922 (0.6) | 922 (0.6) | |
| Secondary | 51,067 (15.3) | 25,555 (15.3) | 25,512 (15.3) | |
| Primary | 242,136 (72.5) | 121,076 (72.5) | 121,060 (72.5) | |
| Healthcare center | 37,170 (11.1) | 18,555 (11.1) | 18,615 (11.1) | |
| Others† | 1,867 (0.6) | 934 (0.6) | 933 (0.6) | |
| ***Follow-up duration, year — median (IQR)*** | | 8.0 (8.0-8.0) | 4.4 (3.1-6.0) | |

Abbreviations: CCI, Charlson's comorbidity index; IQR, interquartile range; NDMA, N-Nitrosodimethylamine; SD, standard deviation.

†Others' healthcare providers included dentistry, psychiatry, and oriental medicine.

**Supplemental Table 4. Baseline characteristics of study population after 1:1:1 propensity score matching.**

| Characteristics | Total (*n*=381,159) | Original valsartan (*n* = 127,053) | Eventually NDMA-uncontaminated valsartan (*n* = 127,053) | NDMA-contained valsartan (*n* = 127,053) | *P* |
|---|---|---|---|---|---|
| ***Age, years — Mean (SD)*** | 57.8 (11.8) | 57.9 (11.7l) | 57.7 (11.9) | 57.9 (11.7) | 0.001‡ |
| ***Sex — n (%)*** | | | | | 1.000 |
| Male | 228,381 (59.9) | 76,127 (59.9) | 76,127(59.9) | 76,127(59.9) | |
| Female | 152,778 (40.1) | 50,926 (40.1) | 50,926 (40.1) | 50,926 (40.1) | |
| ***CCI — n (%)*** | | | | | 0.199 |
| *0* | 133,737 (35.1) | 44,396 (34.9) | 44,964 (35.4) | 44,377 (34.9) | |
| *1* | 104,563(27.4) | 34,953 (27.5) | 34,659 (27.3) | 34,951 (27.5) | |
| *2* | 66,229 (17.4) | 22,079 (17.4) | 22,068 (17.4) | 22,082 (17.4) | |
| ≥ *3* | 76,630 (20.1) | 25,625 (20.2) | 25,362 (19.9) | 25,643 (20.2) | |
| ***Being a prevalent user — n (%)*** | | | | | < 0.001 |
| No | 381,037 (99.9) | 127,026 (99.9) | 127,026 (99.9) | 126,985 (99.9) | |
| Yes | 122 (0.1) | 27 (0.1) | 27 (0.1) | 68 (0.1) | |
| ***Drinking status — n (%)*** | | | | | 0.421 |
| No | 187,999 (49.3) | 62,762 (49.4) | 62,475 (49.2) | 62,762 (49.4) | |
| Yes | 193,160 (50.7) | 64,291(50.6) | 64,578 (50.8) | 64,291 (50.6) | |
| ***Smoking status — n (%)*** | | | | | <0.001 |
| Never | 211466 (55.5) | 70687 (55.6) | 70271 (55.3) | 70508 (55.5) | |

| | | | | | |
|---|---|---|---|---|---|
| Ever | 72828 (19.1) | 25247 (19.9) | 23595 (18.6) | 23986 (18.9) | |
| Current | 96713 (25.4) | 31072 (24.5) | 33126 (26.1) | 32515 (25.6) | |
| Missing | 152 (0.04) | 47 (0.04) | 61 (0.04) | 44 (0.03) | |
| ***Disability — n (%)*** | | | | | 0.255 |
| No | 37,006 (9.7) | 12,277 (9.7) | 12,477 (9.8) | 12,252 (9.6) | |
| Yes | 344,153 (90.3) | 114,776 (90.3) | 114,576 (90.2) | 114,801 (90.4) | |
| ***Level of healthcare provider — n (%)*** | | | | | 0.009 |
| Tertiary | 582 (0.2) | 194 (0.2) | 194 (0.2) | 194 (0.2) | |
| Secondary | 53,386 (14.0) | 17,771 (14.0) | 17,867 (14.1) | 17,748 (14.0) | |
| Primary | 292,752 (76.8) | 99,799 (77.0) | 97,167 (76.5) | 97,786 (77.0) | |
| Healthcare center | 31,984 (8.4) | 10,495 (8.3) | 10,953 (8.6) | 10,536 (8.3) | |
| Others† | 2,455 (0.6) | 794 (0.6l | 872 (0.7) | 789 (0.6) | |
| ***Follow-up duration, year — median (IQR)*** | | 8.0 (8.0-8.0) | 3.5 (1.8-5.2) | 4.2 (3.0-5.7) | |

Abbreviations: CCI, Charlson's comorbidity index; IQR, interquartile range; NDMA, N-Nitrosodimethylamine; SD, standard deviation.

†Others' healthcare providers included dentistry, psychiatry, and oriental medicine.

‡Welch's *t*-test was performed for unequal population variances.

**Supplemental Table 5. The number of any cancer and each organ-specific cancer before and after propensity score matching.**

| Cancer | PSM | Before | | | After 1:1:1 | | |
|---|---|---|---|---|---|---|---|
| | | Original (*n* = 2,340,729) | Eventually NDMA-uncontaminated (*n* = 392,604) | NDMA-contaminated (*n* = 497,879) | Original (*n* = 127,053) | Eventually NDMA-uncontaminated (*n* = 127,053) | NDMA-contaminated (*n* = 127,053) |
| **Any cancer** | Event | 361,565 | 50,963 | 71,695 | 18,615 | 17,276 | 18,406 |
| | IR (95% CI)† | 21.54 (21.47-21.61) | 17.49 (17.35-17.65) | 19.77 (19.62-19.91) | 19.87 (19.59-20.16) | 18.27 (18.00-18.54) | 19.61 (19.33-19.89) |
| **Bladder** | Event | 15,869 | 2,076 | 2,990 | 811 | 739 | 793 |
| | IR (95% CI)† | 0.88 (0.87-0.90) | 0.68 (0.65-0.70) | 0.78 (0.75-0.80) | 0.81 (0.76-0.87) | 0.74 (0.69-0.79) | 0.79 (0.74-0.85) |
| **Breast** | Event | 12,696 | 1,697 | 2,301 | 562 | 541 | 520 |
| | IR (95% CI)† | 1.53 (1.50-1.56) | 1.26 (1.20-1.32) | 1.24 (1.19-1.29) | 1.41 (1.29-1.53) | 1.35 (1.24-1.47) | 1.30 (1.19-1.42) |
| **Colorectal** | Event | 41,550 | 6,212 | 8,709 | 2,096 | 1,997 | 2,090 |
| | IR (95% CI)† | 2.33 (2.31-2.35) | 2.03 (1.98-2.08) | 2.27 (2.22-2.32) | 2.08 (2.00-2.18) | 2.00 (1.922-.09) | 2.10 (2.01-2.20) |
| **Gallbladder and etc.** | Event | 8,829 | 1,222 | 1,748 | 424 | 383 | 387 |
| | IR (95% CI)† | 0.49 (0.48-0.50) | 0.40 (0.68-0.42) | 0.45 (0.43-0.47) | 0.42 (0.38-0.47) | 0.38 (0.34-0.42) | 0.39 (0.35-0.43) |
| **Kidney** | Event | 8,295 | 1,104 | 1,511 | 415 | 386 | 386 |
| | IR (95% CI)† | 0.46 (0.45-0.47) | 0.36 (0.34-0.38) | 0.39 (0.37-0.41) | 0.42 (0.38-0.46) | 0.38 (0.35-0.43) | 0.39 (0.35-0.43) |
| **Liver** | Event | 47,690 | 7,084 | 9,161 | 2,506 | 2,396 | 2,314 |
| | IR (95% CI)† | 2.68 (2.65-2.70) | 2.32 (2.26-2.37) | 2.39 (2.34-2.44) | 2.53 (2.43-2.63) | 2.41 (2.31-2.50) | 2.33 (2.24-2.43) |
| **Lung** | Event | 34,351 | 4,561 | 1.6,731 | 1,671 | 1,453 | 1,624 |

| | | | | | | | |
|---|---|---|---|---|---|---|---|
| | IR (95% CI)[†] | 1.92 (1.90-1.94) | 1.49 (1.44-1.53) | 1.75 (1.71-1.79) | 1.68 (1.60-1.76) | 1.45 (1.38-1.53) | 1.63 (1.55-1.71) |
| **Pancreatic** | Event | 26,524 | 3,659 | 5,373 | 1,408 | 1,276 | 1,460 |
| | IR (95% CI)[†] | 1.48 (1.46-1.50) | 1.19 (1.15-1.23) | 1.40 (1.36-1.43) | 1.41 (1.34-1.49) | 1.28 (1.21-1.35) | 1.47 (1.39-1.54) |
| **Prostate** | Event | 65,962 | 9,351 | 13,425 | 3,916 | 3,570 | 4,010 |
| | IR (95% CI)[†] | 7.02 (6.96-7.07) | 5.51 (5.40-5.62) | 6.88 (6.76-6.99) | 6.69 (6.48-6.90) | 6.06 (5.87-6.27) | 6.85 (6.64-7.07) |
| **Stomach** | Event | 28,858 | 4,156 | 6,030 | 1,148 | 1,385 | 1,490 |
| | IR (95% CI)[†] | 1.61 (1.59-1.63) | 1.36 (1.31-1.40) | 1.57 (1.53-1.61) | 1.46 (1.38-1.53) | 1.39 (1.31-1.46) | 1.50 (1.42-1.57) |
| **Thyroid** | Event | 21,928 | 3,221 | 4,283 | 1,206 | 1,170 | 1,219 |
| | IR (95% CI)[†] | 1.22 (1.21-1.24) | 1.05 (1.01-1.09) | 1.11 (1.08-1.15) | 1.21 (1.14-1.28) | 1.17 (1.10-1.24) | 1.22 (1.16-1.29) |
| **Uterine** | Event | 4,238 | 663 | 895 | 109 | 108 | 133 |
| | IR (95% CI)[†] | 0.51 (0.49-0.52) | 0.49 (0.46-0.53) | 0.48 (0.45-0.51) | 0.45 (0.38-0.52) | 0.50 (0.44-0.58) | 0.48 (0.41-0.55) |

Abbreviations: CI, confidence interval; IR, incidence rate; NDMA, N-Nitrosodimethylamine; PSM, propensity score matching.

[†]Incidence rate per 1,000 person-years is calculated for each valsartan group and clinical outcome.

**Supplemental Table 6. The risks of overall and organ-specific cancers in original population before and after propensity score matching according to the status of NDMA.**

| | Cancer | | Any cancer | | | Lung cancer | | | Prostate cancer† | | |
|---|---|---|---|---|---|---|---|---|---|---|---|
| PSM | | | Event | aHR (95% CI) | *P* | Event | aHR (95% CI) | *P* | Event | aHR (95% CI) | *P* |
| Before PSM‡ | Overall | NDMA-unexposed | 410341 | 1.000 | - | 38101 | 1.000 | - | 75234 | 1.000 | - |
| | | Eventually NDMA-unexposed | 27566 | 0.975 (0.963-0.988) | <0.001 | 2824 | 1.002 (0.963-1.042) | 0.933 | 4934 | 0.948 (0.920-0.976) | <0.001 |
| | | NDMA-exposed | 46234 | 0.984 (0.974-0.993) | 0.001 | 4718 | 0.979 (0.949-1.010) | 0.191 | 8570 | 1.024 (1.000-1.048) | 0.045 |
| | 6 mon exc. | NDMA-unexposed | 225149 | 1.000 | - | 22900 | 1.000 | - | 41112 | 1.000 | - |
| | | Eventually NDMA-unexposed | 25709 | 1.348 (1.330-1.366) | <0.001 | 2643 | 1.360 (1.304-1.418) | <0.001 | 4618 | 1.313 (1.272-1.356) | <0.001 |
| | | NDMA-exposed | 43789 | 1.277 (1.264-1.291) | <0.001 | 4491 | 1.258 (1.217-1.300) | <0.001 | 8149 | 1.342 (1.310-1.376) | <0.001 |
| | 1-year exc. | NDMA-unexposed | 22439 | 1.000 | - | 20495 | 1.000 | - | 36959 | 1.000 | - |
| | | Eventually NDMA-unexposed | 2606 | 1.241 (1.223-1.259) | <0.001 | 2296 | 1.269 (1.213-1.327) | <0.001 | 3965 | 1.204 (1.164-1.246) | <0.001 |
| | | NDMA-exposed | 4558 | 1.206 (1.193-1.220) | <0.001 | 3970 | 1.187 (1.146-1.229) | <0.001 | 7281 | 1.275 (1.243-1.308) | <0.001 |
| | 2-year exc. | NDMA-unexposed | 17421 | 1.000 | - | 16122 | 1.000 | - | 29353 | 1.000 | - |
| | | Eventually NDMA-unexposed | 1860 | 1.097 (1.079-1.116) | <0.001 | 2991 | 1.135 (1.076-1.196) | <0.001 | 2855 | 1.066 (1.024-1.109) | 0.002 |

| | | | | | | | | | | | |
|---|---|---|---|---|---|---|---|---|---|---|---|
| | | NDMA-exposed | 3430 | 1.098 (1.084-1.113) | <0.001 | 1655 | 1.091 (1.048-1.136) | <0.001 | 5455 | 1.159 (1.125-1.194) | <0.001 |
| 1:1 PSM§ | Overall | NDMA-unexposed | 34370 | 1.000 | - | 2994 | 1.000 | - | 7072 | 1.000 | - |
| | | NDMA-exposed | 16253 | 1.014 (0.993-1.035) | 0.191 | 1595 | 1.032 (0.966-1.103) | 0.351 | 3375 | 1.023 (0.978-1.071) | 0.325 |
| | 6 mon exc. | NDMA-unexposed | 14179 | 1.000 | - | 1493 | 1.000 | - | 3023 | 1.000 | - |
| | | NDMA-exposed | 14313 | 1.325 (1.294-1.358) | <0.001 | 1513 | 1.337 (1.242-1.440) | <0.001 | 3097 | 1.323 (1.257-1.394) | <0.001 |
| | 1-year exc. | NDMA-unexposed | 12422 | 1.000 | - | 1330 | 1.000 | - | 2664 | 1.000 | - |
| | | NDMA-exposed | 12607 | 1.239 (1.208-1.271) | <0.001 | 1330 | 1.229 (1.137-1.328) | <0.001 | 2772 | 1.253 (1.187-1.323) | <0.001 |
| | 2-year exc. | NDMA-unexposed | 9323 | 1.000 | - | 1015 | 1.000 | - | 2073 | 1.000 | - |
| | | NDMA-exposed | 9339 | 1.120 (1.088-1.153) | <0.001 | 999 | 1.113 (1.019-1.216) | 0.018 | 2050 | 1.099 (1.033-1.169) | 0.003 |
| 1:1:1 PSM§ | Overall | NDMA-unexposed | 33533 | 1.000 | - | 2761 | 1.000 | - | 7109 | 1.000 | - |
| | | Eventually NDMA-unexposed | 9271 | 1.021 (0.995-1.047) | 0.113 | 879 | 1.025 (0.943-1.114) | 0.557 | 1878 | 0.948 (0.896-1.002) | 0.061 |
| | | NDMA-exposed | 11492 | 1.037 (1.012-1.062) | 0.003 | 1108 | 1.062 (0.982-1.148) | 0.132 | 2509 | 1.045 (0.993-1.101) | 0.093 |
| | 6 mon exc. | NDMA-unexposed | 9044 | 1.000 | - | 1017 | 1.000 | - | 2145 | 1.000 | - |
| | | Eventually NDMA-unexposed | 7365 | 1.496 (1.449-1.544) | <0.001 | 810 | 1.479 (1.343-1.628) | <0.001 | 1654 | 1.377 (1.288-1.471) | <0.001 |
| | | NDMA-exposed | 9170 | 1.495 (1.450-1.544) | <0.001 | 1040 | 1.518 (1.387-1.662) | <0.001 | 2205 | 1.474 (1.385-1.569) | <0.001 |

| | | | | | | | | | | | |
|---|---|---|---|---|---|---|---|---|---|---|---|
| | 1-year exc. | NDMA-unexposed | 7805 | 1.000 | - | 889 | 1.000 | - | 1878 | 1.000 | - |
| | | Eventually NDMA-unexposed | 6165 | 1.309 (1.265-1.355) | <0.001 | 701 | 1.326 (1.198-1.468)[¶] | <0.001 | 1398 | 1.208 (1.125-1.297) | <0.001 |
| | | NDMA-exposed | 7912 | 1.333 (1.291-1.377) | <0.001 | 908 | 1.357 (1.233-1.493)[¶] | <0.001 | 1951 | 1.341 (1.256-1.431) | <0.001 |
| | 2-year exc. | NDMA-unexposed | 5630 | 1.000 | - | 665 | 1.000 | - | 1421 | 1.000 | - |
| | | Eventually NDMA-unexposed | 4230 | 1.098 (1.054-1.143) | <0.001 | 497 | 1.116 (0.992-1.256)[¶] | 0.067 | 973 | 0.990 (0.912-1.075) | 0.809 |
| | | NDMA-exposed | 5603 | 1.142 (1.100-1.186) | <0.001 | 657 | 1.152 (1.032-1.285)[¶] | 0.011 | 1409 | 1.129 (1.048-1.217) | 0.001 |

| | | **Cancer** | **Bladder cancer** | | | **Breast cancer[†]** | | | **Colorectal cancer** | | |
|---|---|---|---|---|---|---|---|---|---|---|---|
| **PSM** | | | **Event** | **aHR (95% CI)** | ***P*** | **Event** | **aHR (95% CI)** | ***P*** | **Event** | **aHR (95% CI)** | ***P*** |
| Before PSM[‡] | Overall | NDMA-unexposed | 17768 | 1.000 | - | 14354 | 1.000 | - | 47800 | 1.000 | - |
| | | Eventually NDMA-unexposed | 1146 | 0.868 (0.817-0.924) | <0.001 | 885 | 0.985 (0.918-1.056) | 0.667 | 3233 | 1.048 (1.010-1.087) | 0.013 |
| | | NDMA-exposed | 2021 | 0.921 (0.879-0.966) | 0.001 | 1455 | 0.910 (0.861-0.963) | 0.001 | 5437 | 1.032 (1.003-1.063) | 0.032 |
| | 6 mon exc. | NDMA-unexposed | 10293 | 1.000 | - | 7626 | 1.000 | - | 25211 | 1.000 | - |
| | | Eventually NDMA-unexposed | 1082 | 1.206 (1.130-1.287) | <0.001 | 831 | 1.428 (1.325-1.538) | <0.001 | 3014 | 1.504 (1.446-1.565) | <0.001 |
| | | NDMA-exposed | 1915 | 1.181 (1.123-1.242) | <0.001 | 1378 | 1.211 (1.141-1.284) | <0.001 | 5121 | 1.375 (1.333-1.418) | <0.001 |

| | | | | | | | | | | | |
|---|---|---|---|---|---|---|---|---|---|---|---|
| | 1-year exc. | NDMA-unexposed | 9399 | 1.000 | - | 6867 | 1.000 | - | 22439 | 1.000 | - |
| | | Eventually NDMA-unexposed | 924 | 1.105 (1.030-1.185) | 0.005 | 727 | 1.334 (1.232-1.444) | <0.001 | 2606 | 1.374 (1.318-1.434) | <0.001 |
| | | NDMA-exposed | 1722 | 1.127 (1.069-1.188) | <0.001 | 1234 | 1.152 (1.082-1.226) | <0.001 | 4558 | 1.295 (1.253-1.339) | <0.001 |
| | 2-year exc. | NDMA-unexposed | 7564 | 1.000 | - | 5416 | 1.000 | - | 17421 | 1.000 | - |
| | | Eventually NDMA-unexposed | 691 | 1.008 (0.930-1.093) | 0.850 | 513 | 1.173 (1.068-1.289) | 0.001 | 1860 | 1.203 (1.144-1.264) | <0.001 |
| | | NDMA-exposed | 1320 | 1.036 (0.975-1.100) | 0.255 | 880 | 1.005 (0.934-1.081) | 0.896 | 3430 | 1.184 (1.140-1.230) | <0.001 |
| 1:1 PSM§ | Overall | NDMA-unexposed | 1474 | 1.000 | - | 1065 | 1.000 | - | 3926 | 1.000 | - |
| | | NDMA-exposed | 708 | 0.937 (0.849-1.033) | 0.191 | 499 | 1.008 (0.896-1.134) | 0.893 | 1764 | 1.056 (0.992-1.125) | 0.088 |
| | 6 mon exc. | NDMA-unexposed | 677 | 1.000 | - | 481 | 1.000 | - | 1517 | 1.000 | - |
| | | NDMA-exposed | 668 | 1.234 (1.105-1.379) | <0.001 | 469 | 1.261 (1.105-1.439) | 0.001 | 1636 | 1.449 (1.348-1.558) | <0.001 |
| | 1-year exc. | NDMA-unexposed | 613 | 1.000 | - | 442 | 1.000 | - | 1340 | 1.000 | - |
| | | NDMA-exposed | 608 | 1.173 (1.045-1.315) | 0.007 | 425 | 1.169 (1.019-1.340) | 0.025 | 1462 | 1.348 (1.250-1.455) | <0.001 |
| | 2-year exc. | NDMA-unexposed | 459 | 1.000 | - | 345 | 1.000 | - | 1011 | 1.000 | - |
| | | NDMA-exposed | 474 | 1.120 (0.984-1.276) | 0.087 | 309 | 0.986 (0.843-1.152) | 0.856 | 1107 | 1.229 (1.128-1.340) | <.0001 |
| 1:1:1 PSM§ | Overall | NDMA-unexposed | 1413 | 1.000 | - | 1015 | 1.000 | - | 3919 | 1.000 | - |

| | | | | | | | | | | | |
|---|---|---|---|---|---|---|---|---|---|---|---|
| | | Eventually NDMA-unexposed | 421 | 0.997 (0.884-1.125)[¶] | 0.997 | 283 | 1.064 (0.918-1.233) | 0.411 | 1001 | 1.058 (0.979-1.143) | 0.155 |
| | | NDMA-exposed | 509 | 0.995 (0.888-1.115)[¶] | 0.929 | 325 | 1.001 (0.869-1.154) | 0.984 | 1236 | 1.071 (0.995-1.152) | 0.067 |
| | 6 mon exc. | NDMA-unexposed | 468 | 1.000 | - | 306 | 1.000 | - | 1045 | 1.000 | - |
| | | Eventually NDMA-unexposed | 392 | 1.495 (1.300-1.719)[¶] | <0.001 | 267 | 1.627 (1.370-1.932) | <0.001 | 917 | 1.673 (1.525-1.835) | <0.001 |
| | | NDMA-exposed | 475 | 1.443 (1.262-1.650)[¶] | <0.001 | 304 | 1.451 (1.227-1.715) | <0.001 | 1131 | 1.637 (1.498-1.788) | <0.001 |
| | 1-year exc. | NDMA-unexposed | 424 | 1.000 | - | 281 | 1.000 | - | 907 | 1.000 | - |
| | | Eventually NDMA-unexposed | 324 | 1.238 (1.068-1.437)[¶] | 0.005 | 232 | 1.388 (1.159-1.661) | <0.001 | 785 | 1.466 (1.329-1.618) | <0.001 |
| | | NDMA-exposed | 428 | 1.285 (1.119-1.476)[¶] | <0.001 | 268 | 1.245 (1.046-1.481) | 0.014 | 1004 | 1.470 (1.340-1.613) | <0.001 |
| | 2-year exc. | NDMA-unexposed | 305 | 1.000 | - | 211 | 1.000 | - | 675 | 1.000 | - |
| | | Eventually NDMA-unexposed | 251 | 1.191 (1.005-1.410)[¶] | 0.043 | 173 | 1.194 (0.974-1.465) | 0.088 | 567 | 1.254 (1.120-1.405) | <0.001 |
| | | NDMA-exposed | 330 | 1.220 (1.041-1.428)[¶] | 0.014 | 193 | 1.034 (0.847-1.262) | 0.741 | 754 | 1.297 (1.167-1.442) | <0.001 |

| | | Cancer | Gallbladder and etc. cancer | | | Kidney cancer | | | Liver cancer | | |
|---|---|---|---|---|---|---|---|---|---|---|---|
| **PSM** | | | **Even** | **aHR (95% CI)** | ***P*** | **Event** | **aHR (95% CI)** | ***P*** | **Event** | **aHR (95% CI)** | ***P*** |
| Before PSM[‡] | Overall | NDMA-unexposed | 9725 | 1.000 | - | 9322 | 1.000 | - | 54593 | 1.000 | - |

| | | | | | | | | | | | |
|---|---|---|---|---|---|---|---|---|---|---|---|
| | | Eventually NDMA-unexposed | 770 | 1.051 (0.974-1.134) | 0.201 | 595 | 0.915 (0.840-0.997) | 0.043 | 3703 | 1.027 (0.992-1.063) | 0.135 |
| | | NDMA-exposed | 1304 | 1.024 (0.964-1.087) | 0.447 | 993 | 0.930 (0.869-0.995) | 0.036 | 5638 | 0.954 (0.927-0.981) | 0.001 |
| | 6 mon exc. | NDMA-unexposed | 6129 | 1.000 | - | 5123 | 1.000 | - | 28856 | 1.000 | - |
| | | Eventually NDMA-unexposed | 719 | 1.365 (1.260-1.480) | <0.001 | 551 | 1.291 (1.179-1.415) | <0.001 | 3458 | 1.453 (1.400-1.507) | <0.001 |
| | | NDMA-exposed | 1224 | 1.247 (1.171-1.329) | <0.001 | 937 | 1.236 (1.150-1.328) | <0.001 | 5299 | 1.259 (1.221-1.297) | <0.001 |
| | 1-year exc. | NDMA-unexposed | 5492 | 1.000 | - | 4629 | 1.000 | - | 25439 | 1.000 | - |
| | | Eventually NDMA-unexposed | 615 | 1.259 (1.154-1.373) | <0.001 | 480 | 1.198 (1.087-1.321) | <0.001 | 2947 | 1.321 (1.270-1.374) | <0.001 |
| | | NDMA-exposed | 1104 | 1.203 (1.126-1.286) | <0.001 | 833 | 1.165 (1.080-1.257) | <0.001 | 4657 | 1.179 (1.141-1.217) | <0.001 |
| | 2-year exc. | NDMA-unexposed | 4384 | 1.000 | - | 3698 | 1.000 | - | 19512 | 1.000 | - |
| | | Eventually NDMA-unexposed | 438 | 1.092 (0.986-1.209) | 0.091 | 368 | 1.138 (1.018-1.272) | 0.023 | 2051 | 1.142 (1.089-1.197) | <0.001 |
| | | NDMA-exposed | 829 | 1.079 (1.000-1.165) | 0.050 | 644 | 1.099 (1.008-1.198) | 0.032 | 3406 | 1.058 (1.019-1.098) | 0.003 |
| 1:1 PSM§ | Overall | NDMA-unexposed | 720 | 1.000 | - | 723 | 1.000 | - | 4588 | 1.000 | - |
| | | NDMA-exposed | 402 | 1.010 (0.884-1.153) | 0.888 | 343 | 0.997 (0.864-1.149) | 0.963 | 1952 | 0.974 (0.918-1.032) | 0.372 |
| | 6 mon exc. | NDMA-unexposed | 349 | 1.000 | - | 309 | 1.000 | - | 1927 | 1.000 | - |

| | | | | | | | | | | | |
|---|---|---|---|---|---|---|---|---|---|---|---|
| | | NDMA-exposed | 339 | 1.239 (1.061-1.446)[¶] | 0.007 | 327 | 1.359 (1.157-1.597) | <0.001 | 1821 | 1.288 (1.205-1.377) | <0.001 |
| | 1-year exc. | NDMA-unexposed | 372 | 1.000 | - | 280 | 1.000 | - | 1678 | 1.000 | - |
| | | NDMA-exposed | 350 | 1.142 (0.983-1.326) | 0.082 | 283 | 1.189 (1.004-1.407) | 0.045 | 1595 | 1.185 (1.105-1.272) | <0.001 |
| | 2-year exc. | NDMA-unexposed | 283 | 1.000 | - | 221 | 1.000 | - | 1221 | 1.000 | - |
| | | NDMA-exposed | 260 | 1.019 (0.860-1.209) | 0.824 | 232 | 1.165 (0.969-1.408) | 0.104 | 1182 | 1.080 (0.996-1.171) | 0.063 |
| 1:1:1 PSM[§] | Overall | NDMA-unexposed | 656 | 1.000 | - | 733 | 1.000 | - | 4627 | 1.000 | - |
| | | Eventually NDMA-unexposed | 248 | 1.109 (0.945-1.302)[¶] | 0.205 | 199 | 0.954 (0.802-1.133)[¶] | 0.590 | 1232 | 1.091 (1.017-1.171) | 0.015 |
| | | NDMA-exposed | 290 | 1.067 (0.914-1.245)[¶] | 0.410 | 254 | 1.002 (0.853-1.177)[¶] | 0.981 | 1356 | 0.978 (0.913-1.047) | 0.519 |
| | 6 mon exc. | NDMA-unexposed | 280 | 1.000 | - | 223 | 1.000 | - | 1332 | 1.000 | - |
| | | Eventually NDMA-unexposed | 229 | 1.455 (1.214-1.744)[¶] | <0.001 | 184 | 1.486 (1.214-1.820)[¶] | <0.001 | 1126 | 1.654 (1.522-1.798) | <0.001 |
| | | NDMA-exposed | 272 | 1.383 (1.162-1.647)[¶] | <0.001 | 242 | 1.541 (1.273-1.864)[¶] | <0.001 | 1242 | 1.441 (1.328-1.564) | <0.001 |
| | 1-year exc. | NDMA-unexposed | 253 | 1.000 | - | 203 | 1.000 | - | 1156 | 1.000 | - |
| | | Eventually NDMA-unexposed | 201 | 1.314 (1.086-1.589)[¶] | 0.005 | 158 | 1.260 (1.018-1.558)[¶] | 0.033 | 959 | 1.431 (1.310-1.563) | <0.001 |
| | | NDMA-exposed | 248 | 1.281 (1.069-1.536)[¶] | 0.007 | 213 | 1.325 (1.087-1.614)[¶] | 0.005 | 1074 | 1.255 (1.152-1.368) | <0.001 |
| | 2-year exc. | NDMA-unexposed | 191 | 1.000 | - | 154 | 1.000 | - | 808 | 1.000 | - |

| | | | | | | | | | | |
|---|---|---|---|---|---|---|---|---|---|---|
| | | Eventually NDMA-unexposed | 141 | 1.066 (0.855-1.328)[¶] | 0.572 | 128 | 1.226 (0.967-1.555)[¶] | 0.092 | 667 | 1.205 (1.086-1.337) | <0.001 |
| | | NDMA-exposed | 178 | 1.050 (0.854-1.291)[¶] | 0.645 | 178 | 1.318 (1.059-1.641)[¶] | 0.013 | 777 | 1.093 (0.989-1.208) | 0.081 |

| | | Cancer | Pancreatic cancer | | | Stomach cancer | | | Thyroid cancer | | |
|---|---|---|---|---|---|---|---|---|---|---|---|
| PSM | | | Event | aHR (95% CI) | *P* | Event | aHR (95% CI) | *P* | Event | aHR (95% CI) | *P* |
| Before PSM[‡] | Overall | NDMA-unexposed | 29789 | 1.000 | - | 32955 | 1.000 | - | 25606 | 1.000 | - |
| | | Eventually NDMA-unexposed | 2137 | 0.959 (0.916-1.003) | 0.068 | 2237 | 1.071 (1.024-1.119) | 0.003 | 1439 | 0.918 (0.870-0.970) | 0.002 |
| | | NDMA-exposed | 3630 | 0.973 (0.939-1.008) | 0.132 | 3852 | 1.067 (1.031-1.104) | <0.001 | 2387 | 0.917 (0.878-0.958) | <0.001 |
| | 6 mon exc. | NDMA-unexposed | 17804 | 1.000 | - | 17975 | 1.000 | - | 11950 | 1.000 | - |
| | | Eventually NDMA-unexposed | 2002 | 1.283 (1.223-1.346) | <0.001 | 2067 | 1.478 (1.409-1.550) | <0.001 | 1335 | 1.358 (1.281-1.440) | <0.001 |
| | | NDMA-exposed | 3444 | 1.224 (1.179-1.271) | <0.001 | 3627 | 1.393 (1.343-1.445) | <0.001 | 2237 | 1.262 (1.204-1.322) | <0.001 |
| | 1-year exc. | NDMA-unexposed | 16035 | 1.000 | - | 15901 | 1.000 | - | 10512 | 1.000 | - |
| | | Eventually NDMA-unexposed | 1769 | 1.215 (1.154-1.278) | <0.001 | 1789 | 1.364 (1.297-1.436) | <0.001 | 1152 | 1.235 (1.159-1.315) | <0.001 |
| | | NDMA-exposed | 3053 | 1.154 (1.109-1.201) | <0.001 | 3191 | 1.312 (1.261-1.364) | <0.001 | 1988 | 1.184 (1.127-1.244) | <0.001 |
| | 2-year exc. | NDMA-unexposed | 12923 | 1.000 | - | 12218 | 1.000 | - | 8247 | 1.000 | - |

| | | | | | | | | | | |
|---|---|---|---|---|---|---|---|---|---|---|
| | | Eventually NDMA-unexposed | 1298 | 1.095 (1.032-1.162) | 0.003 | 1235 | 1.168 (1.099-1.242) | <0.001 | 830 | 1.087 (1.009-1.170) | 0.027 |
| | | NDMA-exposed | 2331 | 1.061 (1.014-1.111) | 0.010 | 2337 | 1.182 (1.129-1.237) | <0.001 | 1496 | 1.076 (1.017-1.149) | 0.011 |
| 1:1 PSM§ | Overall | NDMA-unexposed | 2544 | 1.000 | - | 2811 | 1.000 | - | 2392 | 1.000 | - |
| | | NDMA-exposed | 1347 | 1.043 (0.970-1.121) | 0.259 | 1328 | 1.037 (0.964-1.114) | 0.328 | 900 | 0.910 (0.835-0.992) | 0.032 |
| | 6 mon exc. | NDMA-unexposed | 1225 | 1.000 | - | 1237 | 1.000 | - | 855 | 1.000 | - |
| | | NDMA-exposed | 1267 | 1.328 (1.224-1.441) | <0.001 | 1245 | 1.357 (1.251-1.472) | <0.001 | 845 | 1.256 (1.138-1.386) | <0.001 |
| | 1-year exc. | NDMA-unexposed | 1088 | 1.000 | - | 1084 | 1.000 | - | 752 | 1.000 | - |
| | | NDMA-exposed | 1116 | 1.218 (1.118-1.327) | <0.001 | 1109 | 1.286 (1.180-1.401) | <0.001 | 754 | 1.167 (1.052-1.294) | 0.004 |
| | 2-year exc. | NDMA-unexposed | 850 | 1.000 | - | 830 | 1.000 | - | 589 | 1.000 | - |
| | | NDMA-exposed | 853 | 1.092 (0.992-1.202) | 0.072 | 841 | 1.165 (1.057-1.284) | 0.002 | 564 | 1.037 (0.922-1.166) | 0.545 |
| 1:1:1 PSM§ | Overall | NDMA-unexposed | 2451 | 1.000 | - | 2648 | 1.000 | - | 2427 | 1.000 | - |
| | | Eventually NDMA-unexposed | 724 | 0.982 (0.896-1.075) | 0.690 | 754 | 1.086 (0.993-1.189) | 0.071 | 528 | 0.961 (0.864-1.067) | 0.454 |
| | | NDMA-exposed | 969 | 1.076 (0.989-1.171) | 0.087 | 921 | 1.086 (0.997-1.182) | 0.059 | 640 | 0.949 (0.859-1.049) | 0.306 |
| | 6 mon exc. | NDMA-unexposed | 862 | 1.000 | - | 817 | 1.000 | - | 579 | 1.000 | - |
| | | Eventually NDMA-unexposed | 664 | 1.401 (1.261-1.566)¶ | <0.001 | 683 | 1.604 (1.443-1.783) | <0.001 | 476 | 1.486 (1.309-1.686)¶ | <0.001 |

| | | | | | | | | | | | |
|---|---|---|---|---|---|---|---|---|---|---|---|
| | | NDMA-exposed | 902 | 1.502 (1.362-1.657)¶ | <0.001 | 860 | 1.601 (1.448-1.771) | <0.001 | 589 | 1.441 (1.277-1.625)¶ | <0.001 |
| | 1-year exc. | NDMA-unexposed | 761 | 1.000 | - | 715 | 1.000 | - | 508 | 1.000 | - |
| | | Eventually NDMA-unexposed | 575 | 1.237 (1.107-1.382)¶ | <0.001 | 588 | 1.421 (1.260-1.591) | <0.001 | 410 | 1.299 (1.136-1.485)¶ | <0.001 |
| | | NDMA-exposed | 791 | 1.327 (1.197-1.470)¶ | <0.001 | 759 | 1.441 (1.296-1.601) | <0.001 | 525 | 1.290 (1.137-1.463)¶ | <0.001 |
| | 2-year exc. | NDMA-unexposed | 589 | 1.000 | - | 539 | 1.000 | - | 395 | 1.000 | - |
| | | Eventually NDMA-unexposed | 427 | 1.061 (0.935-1.204)¶ | 0.356 | 414 | 1.161 (1.020-1.322) | 0.024 | 296 | 1.080 (0.927-1.258)¶ | 0.324 |
| | | NDMA-exposed | 588 | 1.131 (1.007-1.270)¶ | 0.038 | 563 | 1.237 (1.097-1.395) | 0.001 | 385 | 1.083 (0.939-1.249)¶ | 0.276 |

| PSM | | Cancer | Uterine cancer† | | |
|---|---|---|---|---|---|
| | | | Event | aHR (95% CI) | *P* |
| Before PSM‡ | Overall | NDMA-unexposed | 4793 | 1.000 | - |
| | | Eventually NDMA-unexposed | 355 | 0.933 (0.835-1.042) | 0.220 |
| | | NDMA-exposed | 648 | 1.004 (0.922-1.093) | 0.933 |
| | 6 mon exc. | NDMA-unexposed | 2789 | 1.000 | - |
| | | Eventually NDMA-unexposed | 339 | 1.263 (1.125-1.419) | <0.001 |

| | | | | | |
|---|---|---|---|---|---|
| | | NDMA-exposed | 619 | 1.264 (1.155-1.383) | <0.001 |
| | 1-year exc. | NDMA-unexposed | 2527 | 1.000 | - |
| | | Eventually NDMA-unexposed | 301 | 1.198 (1.059-1.355) | 0.004 |
| | | NDMA-exposed | 558 | 1.206 (1.109-1.326) | <0.001 |
| | 2-year exc. | NDMA-unexposed | 2053 | 1.000 | - |
| | | Eventually NDMA-unexposed | 235 | 1.175 (1.022-1.350) | 0.023 |
| | | NDMA-exposed | 425 | 1.132 (1.017-1.261) | 0.023 |
| 1:1 PSM§ | Overall | NDMA-unexposed | 340 | 1.000 | - |
| | | NDMA-exposed | 202 | 1.007 (0.832-1.218) | 0.943 |
| | 6 mon exc. | NDMA-unexposed | 167 | 1.000 | - |
| | | NDMA-exposed | 194 | 1.334 (1.078-1.652) | 0.008 |
| | 1-year exc. | NDMA-unexposed | 142 | 1.000 | - |
| | | NDMA-exposed | 183 | 1.408 (1.125-1.761) | 0.003 |
| | 2-year exc. | NDMA-unexposed | 121 | 1.000 | - |
| | | NDMA-exposed | 139 | 1.207 (0.943-1.545) | 0.135 |

| | | | | | |
|---|---|---|---|---|---|
| 1:1:1 PSM[§] | Overall | NDMA-unexposed | 333 | 1.000 | - |
| | | Eventually NDMA-unexposed | 108 | 0.902 (0.712-1.144)[¶] | 0.396 |
| | | NDMA-exposed | 133 | 0.887 (0.709-1.110)[¶] | 0.294 |
| | 6 mon exc. | NDMA-unexposed | 106 | 1.000 | - |
| | | Eventually NDMA-unexposed | 104 | 1.515 (1.146-2.001)[¶] | 0.004 |
| | | NDMA-exposed | 127 | 1.422 (1.088-1.857)[¶] | 0.010 |
| | 1-year exc. | NDMA-unexposed | 92 | 1.000 | - |
| | | Eventually NDMA-unexposed | 96 | 1.509 (1.127-2.020)[¶] | 0.006 |
| | | NDMA-exposed | 122 | 1.463 (1.109-1.931)[¶] | 0.007 |
| | 2-year exc. | NDMA-unexposed | 78 | 1.000 | - |
| | | Eventually NDMA-unexposed | 79 | 1.393 (1.015-1.912)[¶] | 0.040 |
| | | NDMA-exposed | 87 | 1.167 (0.857-1.591)[¶] | 0.327 |

Abbreviations: aHR, adjusted hazard ratio; CI, confidence interval; exc., excluded; mon, month; NDMA, N-Nitrosodimethylamine; PSM, propensity score matching.

A shaded area means that each model has the lowest Akaike information criterion.

[†]All analyses for breast and uterine cancer were performed limited to men or women.

[‡]All hazard ratios before PSM were adjusted for age, sex, Charlson's Comorbidity Index, being a prevalent user, and the year of the first observed valsartan

prescription.

[§]All hazard ratios after PSM were adjusted for age, sex, being a prevalent user, and the year of the first observed valsartan prescription.

[¶]All hazard ratios after PSM were adjusted for age, sex, and the year of the first observed valsartan prescription in the population limited to not being a prevalent user.

**Supplemental Table 7. The risks of overall, lung, and prostate cancers in new user population before and after propensity score matching according to the status of NDMA.**

| PSM | | Cancer | All cancer | | | Lung cancer | | | Prostate cancer† | | |
|---|---|---|---|---|---|---|---|---|---|---|---|
| | | | Event | aHR (95% CI) | *P* | Event | aHR (95% CI) | *P* | Event | aHR (95% CI) | *P* |
| Before PSM‡ | Overall | NDMA-unexposed | 306688 | 1.000 | | 27856 | 1.000 | | 55614 | 1.000 | |
| | | Eventually NDMA-unexposed | 27550 | 0.955 (0.943-0.967) | <0.001 | 2823 | 0.968 (0.930-1.007) | 0.105 | 4934 | 0.936 (0.908-0.964) | <0.001 |
| | | NDMA-exposed | 40615 | 0.964 (0.954-0.975) | <0.001 | 4122 | 0.948 (0.916-0.981) | 0.002 | 7497 | 1.012 (0.987-1.038) | 0.351 |
| | 6 mon exc. | NDMA-unexposed | 133428 | 1.000 | | 13675 | 1.000 | | 23781 | 1.000 | |
| | | Eventually NDMA-unexposed | 25704 | 1.234 (1.217-1.251) | <0.001 | 2642 | 1.252 (1.201-1.306) | <0.001 | 4618 | 1.208 (1.170-1.247) | <0.001 |
| | | NDMA-exposed | 38280 | 1.220 (1.206-1.234) | <0.001 | 3916 | 1.207 (1.164-1.252) | <0.001 | 7121 | 1.290 (1.255-1.325) | <0.001 |
| | 1-year exc. | NDMA-unexposed | 116515 | 1.000 | | 11928 | 1.000 | | 20828 | 1.000 | |
| | | Eventually NDMA-unexposed | 22177 | 1.153 (1.136-1.170) | <0.001 | 2295 | 1.181 (1.129-1.236) | <0.001 | 3965 | 1.122 (1.084-1.161) | <0.001 |
| | | NDMA-exposed | 33785 | 1.156 (1.142-1.170) | <0.001 | 3433 | 1.139 (1.096-1.184) | <0.001 | 6318 | 1.228 (1.194-1.264) | <0.001 |
| 1:1 PSM§ | Overall | NDMA-unexposed | 31982 | 1.000 | | 2775 | 1.000 | | 6604 | 1.000 | |
| | | NDMA-exposed | 14198 | 1.014 (0.991-1.036) | 0.231 | 1399 | 1.031 (0.960-1.106) | 0.404 | 2955 | 1.019 (0.970-1.069) | 0.458 |
| | 6 mon exc. | NDMA-unexposed | 12193 | 1.000 | | 1292 | 1.000 | | 2634 | 1.000 | |

| | | | | | | | | | | | |
|---|---|---|---|---|---|---|---|---|---|---|---|
| | | NDMA-exposed | 12349 | 1.323 (1.289-1.358) | <0.001 | 1322 | 1.349 (1.246-1.461) | <0.001 | 2693 | 1.311 (1.240-1.386) | <0.001 |
| | 1-year exc. | NDMA-unexposed | 10595 | 1.000 | | 1137 | 1.000 | | 2304 | 1.000 | |
| | | NDMA-exposed | 10794 | 1.231 (1.198-1.265) | <0.001 | 1150 | 1.233 (1.134-1.342) | <0.001 | 2397 | 1.240 (1.170-1.315) | <0.001 |
| 1:1:1 PSM§ | Overall | NDMA-unexposed | 33518 | 1.000 | | 2760 | 1.000 | | 7106 | 1.000 | |
| | | Eventually NDMA-unexposed | 9265 | 1.021 (0.995-1.047) | 0.111 | 879 | 1.026 (0.944-1.115) | 0.553 | 1878 | 0.949 (0.897-1.003) | 0.066 |
| | | NDMA-exposed | 11487 | 1.037 (1.013-1.063) | 0.003 | 1108 | 1.062 (0.982-1.148) | 0.132 | 2509 | 1.046 (0.994-1.102) | 0.084 |
| | 6 mon exc. | NDMA-unexposed | 9037 | 1.000 | | 1017 | 1.000 | | 2142 | 1.000 | |
| | | Eventually NDMA-unexposed | 7362 | 1.497 (1.449-1.545) | <0.001 | 810 | 1.479 (1.343-1.628) | <0.001 | 1654 | 1.379 (1.290-1.474) | <0.001 |
| | | NDMA-exposed | 9165 | 1.496 (1.451-1.542) | <0.001 | 1040 | 1.518 (1.386-1.662) | <0.001 | 2205 | 1.477 (1.387-1.571) | <0.001 |
| | 1-year exc. | NDMA-unexposed | 7798 | 1.000 | | 889 | 1.000 | | 1875 | 1.000 | |
| | | Eventually NDMA-unexposed | 6162 | 1.310 (1.266-1.355) | <0.001 | 701 | 1.326 (1.198-1.468) | <0.001 | 1398 | 1.210 (1.127-1.299) | <0.001 |
| | | NDMA-exposed | 7907 | 1.334 (1.291-1.377) | <0.001 | 908 | 1.357 (1.233-1.493) | <0.001 | 1951 | 1.344 (1.259-1.434) | <0.001 |

Abbreviations: aHR, adjusted hazard ratio; CI, confidence interval; exc., excluded; mon, month; NDMA, N-Nitrosodimethylamine; PSM, propensity score matching.

A shaded area means that each model has the lowest Akaike information criterion.

†All analyses for uterine cancer were performed limited to men.

‡All hazard ratios before PSM were adjusted for age, sex, Charlson's Comorbidity Index, being a prevalent user, and the year of the first observed valsartan

prescription.

§All hazard ratios after PSM were adjusted for age, sex, being a prevalent user, and the year of the first observed valsartan prescription.

**Supplemental Table 8. The risks of overall and significant organ-specific cancers before and after propensity score matching according to NDMA-contaminated valsartan doses.**

| PSM | | | Any cancer | | | Lung cancer | | | Prostate cancer† | | |
|---|---|---|---|---|---|---|---|---|---|---|---|
| | | | Event | aHR (95% CI) | *P* | Event | aHR (95% CI) | *P* | Event | aHR (95% CI) | *P* |
| Before PSM‡ | Overall | NDMA-unexposed | 383794 | 1.000 | - | 36056 | 1.000 | - | 70289 | 1.000 | - |
| | | < 10800 | 17869 | 1.038 (1.023-1.054) | <0.001 | 1808 | 1.051 (1.002-1.103) | 0.041 | 3218 | 1.059 (1.022-1.098) | 0.002 |
| | | 10800 - 40760 | 16855 | 0.986 (0.970-1.001) | 0.067 | 1761 | 1.017 (0.969-1.067) | 0.501 | 3098 | 1.019 (0.982-1.056) | 0.318 |
| | | 40800 - 525120 | 14740 | 0.948 (0.932-0.964) | <0.001 | 1457 | 0.901 (0.854-0.950) | <0.001 | 2782 | 1.006 (0.967-1.045) | 0.778 |
| | 6 mon exc. | NDMA-unexposed | 225149 | 1.000 | - | 22900 | 1.000 | - | 41112 | 1.000 | - |
| | | < 10800 | 13373 | 1.195 (1.175-1.217) | <0.001 | 1376 | 1.176 (1.113-1.243) | <0.001 | 2451 | 1.252 (1.201-1.304) | <0.001 |
| | | 10800 - 40760 | 15678 | 1.315 (1.293-1.336) | <0.001 | 1659 | 1.331 (1.266-1.400) | <0.001 | 2918 | 1.385 (1.333-1.438) | <0.001 |
| | | 40800 - 525120 | 14738 | 1.101 (1.083-1.120) | <0.001 | 1456 | 1.057 (1.002-1.116) | 0.042 | 2780 | 1.164 (1.119-1.210) | <0.001 |
| | 1-year exc. | NDMA-unexposed | 201123 | 1.000 | - | 20495 | 1.000 | - | 36959 | 1.000 | - |
| | | < 10800 | 11467 | 1.126 (1.105-1.148) | <0.001 | 1180 | 1.113 (1.048-1.181) | <0.001 | 2107 | 1.183 (1.132-1.237) | <0.001 |
| | | 10800 - 40760 | 12927 | 1.186 (1.165-1.208) | <0.001 | 1354 | 1.191 (1.127-1.259) | <0.001 | 2445 | 1.269 (1.217-1.322) | <0.001 |
| | | 40800 - 525120 | 14503 | 1.133 (1.114-1.153) | <0.001 | 1436 | 1.095 (1.037-1.156) | 0.001 | 2729 | 1.195 (1.148-1.243) | <0.001 |
| | 2-year exc. | NDMA-unexposed | 158135 | 1.000 | - | 16122 | 1.000 | - | 29353 | 1.000 | - |

| | | | | | | | | | | | |
|---|---|---|---|---|---|---|---|---|---|---|---|
| | | < 10800 | 8111 | 1.057 (1.034-1.081) | <.0001 | 831 | 1.044 (0.972-1.120) | 0.236 | 1494 | 1.112 (1.055-1.172) | <0.001 |
| | | 10800 - 40760 | 8658 | 1.030 (1.008-1.053) | 0.008 | 937 | 1.069 (1.000-1.143) | 0.049 | 1631 | 1.094 (1.041-1.151) | 0.001 |
| | | 40800 - 525120 | 12359 | 1.108 (1.088-1.129) | <.0001 | 1223 | 1.075 (1.013-1.140) | 0.017 | 2330 | 1.171 (1.122-1.222) | <0.001 |
| 1:1 PSM[§] | Overall | NDMA-unexposed | 33353 | 1.000 | - | 2904 | 1.000 | - | 6879 | 1.000 | - |
| | | < 10800 | 5969 | 1.074 (1.044-1.105) | <0.001 | 567 | 1.078 (0.983-1.182) | 0.113 | 1245 | 1.092 (1.026-1.162) | 0.006 |
| | | 10800 - 40760 | 5890 | 1.027 (0.998-1.057) | 0.068 | 597 | 1.092 (0.996-1.196) | 0.060 | 1191 | 1.004 (0.942-1.070) | 0.900 |
| | | 40800 - 525120 | 5411 | 0.994 (0.963-1.025) | 0.687 | 521 | 0.983 (0.890-1.086) | 0.737 | 1132 | 1.005 (0.940-1.076) | 0.876 |
| | 6 mon exc. | NDMA-unexposed | 14179 | 1.000 | - | 1493 | 1.000 | - | 3023 | 1.000 | - |
| | | < 10800 | 4168 | 1.263 (1.219-1.308) | <0.001 | 445 | 1.261 (1.133-1.403) | <0.001 | 911 | 1.285 (1.193-1.385) | <0.001 |
| | | 10800 - 40760 | 5070 | 1.402 (1.357-1.448) | <0.001 | 552 | 1.441 (1.305-1.591) | <0.001 | 1088 | 1.383 (1.289-1.484) | <0.001 |
| | | 40800 - 525120 | 5075 | 1.153 (1.116-1.192) | <0.001 | 516 | 1.144 (1.031-1.269) | 0.011 | 1098 | 1.150 (1.071-1.236) | <0.001 |
| | 1-year exc. | NDMA-unexposed | 12422 | 1.000 | - | 1330 | 1.000 | - | 2664 | 1.000 | - |
| | | < 10800 | 3544 | 1.176 (1.132-1.221) | <0.001 | 385 | 1.178 (1.051-1.321) | 0.005 | 770 | 1.188 (1.096-1.288) | <0.001 |
| | | 10800 - 40760 | 4121 | 1.241 (1.198-1.286) | <0.001 | 437 | 1.222 (1.096-1.363) | <0.001 | 921 | 1.269 (1.177-1.369) | <0.001 |
| | | 40800 - 525120 | 4942 | 1.169 (1.130-1.209) | <0.001 | 508 | 1.155 (1.040-1.283) | 0.007 | 1081 | 1.175 (1.093-1.263) | <0.001 |
| | 2-year exc. | NDMA-unexposed | 9323 | 1.000 | - | 1015 | 1.000 | - | 2073 | 1.000 | - |

| | | | | | | | | | | | |
|---|---|---|---|---|---|---|---|---|---|---|---|
| | | < 10800 | 2492 | 1.095 (1.048-1.145) | <0.001 | 262 | 1.057 (0.922-1.211) | 0.430 | 545 | 1.083 (0.986-1.191) | 0.097 |
| | | 10800 - 40760 | 2698 | 1.051 (1.006-1.097) | 0.025 | 301 | 1.069 (0.939-1.216) | 0.316 | 595 | 1.033 (0.942-1.132) | 0.488 |
| | | 40800 - 525120 | 4149 | 1.129 (1.087-1.171) | <0.001 | 436 | 1.125 (1.004-1.261) | 0.043 | 910 | 1.104 (1.020-1.195) | 0.014 |
| 1:1:1 PSM[§] | Overall | NDMA-unexposed | 24777 | 1.000 | - | 2113 | 1.000 | - | 5281 | 1.000 | - |
| | | < 10800 | 4420 | 1.083 (1.047-1.119) | <0.001 | 421 | 1.108 (0.995-1.235) | 0.063 | 954 | 1.090 (1.015-1.171) | 0.019 |
| | | 10800 - 40760 | 4137 | 1.040 (1.004-1.076) | 0.027 | 411 | 1.107 (0.991-1.235) | 0.071 | 871 | 1.013 (0.940-1.092) | 0.734 |
| | | 40800 - 525120 | 3686 | 1.003 (0.967-1.042) | 0.858 | 350 | 0.996 (0.883-1.124) | 0.951 | 820 | 1.013 (0.936-1.097) | 0.747 |
| | 6 mon exc. | NDMA-unexposed | 9816 | 1.000 | - | 1020 | 1.000 | - | 2210 | 1.000 | - |
| | | < 10800 | 3081 | 1.256 (1.206-1.309) | <0.001 | 324 | 1.267 (1.117-1.438) | <0.001 | 711 | 1.278 (1.173-1.391) | <0.001 |
| | | 10800 - 40760 | 3511 | 1.408 (1.354-1.465) | <0.001 | 377 | 1.463 (1.298-1.650) | <0.001 | 787 | 1.370 (1.262-1.488) | <0.001 |
| | | 40800 - 525120 | 3420 | 1.153 (1.107-1.201) | <0.001 | 347 | 1.165 (1.027-1.322) | 0.018 | 788 | 1.136 (1.044-1.235) | 0.003 |
| | 1-year exc. | NDMA-unexposed | 8558 | | - | 893 | | - | 1946 | | - |
| | | < 10800 | 2610 | 1.167 (1.117-1.220) | <0.001 | 281 | 1.199 (1.047-1.372)[¶] | 0.009 | 597 | 1.181 (1.077-1.296) | <0.001 |
| | | 10800 - 40760 | 2819 | 1.234 (1.182-1.289) | <0.001 | 290 | 1.220 (1.067-1.395)[¶] | 0.004 | 664 | 1.260 (1.152-1.377) | <0.001 |
| | | 40800 - 525120 | 3330 | 1.169 (1.122-1.218) | <0.001 | 344 | 1.197 (1.053-1.360)[¶] | 0.006 | 775 | 1.162 (1.067-1.266) | 0.001 |
| | 2-year exc. | NDMA-unexposed | 6291 | 1.000 | - | 670 | 1.000 | - | 1485 | 1.000 | - |

| | | | | | | | | | | | |
|---|---|---|---|---|---|---|---|---|---|---|---|
| | | < 10800 | 1806 | 1.102 (1.046-1.162) | <0.001 | 187 | 1.082 (0.919-1.273)[¶] | 0.*344* | 414 | 1.083 (0.971-1.208) | 0.154 |
| | | 10800 - 40760 | 1771 | 1.035 (0.982-1.092) | 0.200 | 186 | 1.030 (0.874-1.213)[¶] | 0.728 | 417 | 1.029 (0.922-1.149) | 0.608 |
| | | 40800 - 525120 | 2770 | 1.138 (1.087-1.191) | <0.001 | 291 | 1.170 (1.017-1.346)[¶] | 0.028 | 649 | 1.107 (1.008-1.216) | 0.033 |

| | | | **Colorectal cancer** | | | **Kidney cancer** | | | **Stomach cancer** | | |
|---|---|---|---|---|---|---|---|---|---|---|---|
| **PSM** | | | **Event** | **aHR (95% CI)** | ***P*** | **Event** | **aHR (95% CI)** | ***P*** | **Event** | **aHR (95% CI)** | ***P*** |
| Before PSM[‡] | Overall | NDMA-unexposed | 44424 | 1.000 | - | 8754 | 1.000 | - | 30760 | 1.000 | - |
| | | < 10800 | 2085 | 1.081 (1.034-1.130) | 0.001 | 325 | 0.841 (0.752-0.940) | 0.002 | 1472 | 1.111 (1.054-1.171) | <0.001 |
| | | 10800 - 40760 | 1967 | 1.023 (0.978-1.071) | 0.320 | 390 | 1.016 (0.917-1.126) | 0.759 | 1390 | 1.046 (0.991-1.105) | 0.102 |
| | | 40800 - 525120 | 1782 | 1.044 (0.995-1.096) | 0.079 | 337 | 0.945 (0.846-1.056) | 0.321 | 1266 | 1.073 (1.013-1.136) | 0.017 |
| | 6 mon exc. | NDMA-unexposed | 25211 | 1.000 | - | 5123 | 1.000 | - | 17975 | 1.000 | - |
| | | < 10800 | 1528 | 1.251 (1.187-1.318) | <0.001 | 242 | 0.979 (0.859-1.115) | 0.750 | 1071 | 1.251 (1.175-1.331) | <0.001 |
| | | 10800 - 40760 | 1811 | 1.388 (1.323-1.457) | <0.001 | 358 | 1.379 (1.237-1.537) | <0.001 | 1290 | 1.400 (1.322-1.482) | <0.001 |
| | | 40800 - 525120 | 1782 | 1.235 (1.176-1.297) | <0.001 | 337 | 1.129 (1.009-1.263) | 0.034 | 1266 | 1.259 (1.188-1.335) | <0.001 |
| | 1-year exc. | NDMA-unexposed | 22439 | 1.000 | - | 4629 | 1.000 | - | 15901 | 1.000 | - |
| | | < 10800 | 1319 | 1.179 (1.114-1.247) | <0.001 | 203 | 0.897 (0.779-1.034) | 0.135 | 917 | 1.184 (1.107-1.267) | <0.001 |
| | | 10800 - 40760 | 1491 | 1.246 (1.182-1.314) | <0.001 | 299 | 1.257 (1.116-1.415) | <0.001 | 1035 | 1.240 (1.164-1.321) | <0.001 |

| | | | | | | | | | | | |
|---|---|---|---|---|---|---|---|---|---|---|---|
| | | 40800 - 525120 | 1748 | 1.264 (1.203-1.328) | <0.001 | 331 | 1.156 (1.032-1.295) | 0.012 | 1239 | 1.296 (1.222-1.374) | <0.001 |
| | 2-year exc. | NDMA-unexposed | 17421 | 1.000 | - | 3698 | 1.000 | - | 12218 | 1.000 | - |
| | | < 10800 | 961 | 1.139 (1.067-1.217) | <0.001 | 156 | 0.914 (0.778-1.075) | 0.277 | 634 | 1.096 (1.011-1.189) | 0.026 |
| | | 10800 - 40760 | 985 | 1.066 (0.999-1.148) | 0.052 | 208 | 1.130 (0.981-1.301) | 0.090 | 671 | 1.055 (0.976-1.141) | 0.180 |
| | | 40800 - 525120 | 1484 | 1.227 (1.163-1.295) | <0.001 | 280 | 1.128 (0.997-1.276) | 0.055 | 1032 | 1.258 (1.179-1.341) | <0.001 |
| 1:1 PSM[§] | Overall | NDMA-unexposed | 3817 | 1.000 | - | | | | | 1.000 | - |
| | | < 10800 | 632 | 1.076(0.986-1.174) | 0.098 | | | | | 1.156 (1.048-1.275) | 0.004 |
| | | 10800 - 40760 | 638 | 1.053 (0.964-1.149) | 0.251 | | | | | 1.035 (0.935-1.145) | 0.504 |
| | | 40800 - 525120 | 603 | 1.093 (0.996-1.200) | 0.061 | | | | | 1.013 (0.909-1.128) | 0.819 |
| | 6 mon exc. | NDMA-unexposed | 1517 | 1.000 | - | | | | | 1.000 | - |
| | | < 10800 | 454 | 1.296 (1.166-1.441) | <0.001 | | | | | 1.325 (1.179-1.490) | <0.001 |
| | | 10800 - 40760 | 588 | 1.537 (1.395-1.693) | <0.001 | | | | | 1.399 (1.252-1.562) | <0.001 |
| | | 40800 - 525120 | 597 | 1.319 (1.195-1.456) | <0.001 | | | | | 1.177 (1.051-1.317) | 0.005 |
| | 1-year exc. | NDMA-unexposed | 1340 | 1.000 | - | | | | | 1.000 | - |
| | | < 10800 | 395 | 1.213 (1.083-1.358) | 0.001 | | | | | 1.303 (1.152-1.474) | <0.001 |
| | | 10800 - 40760 | 480 | 1.343 (1.209-1.492) | <0.001 | | | | | 1.221 (1.081-1.379) | 0.001 |

| | | | | | | | | | | |
|---|---|---|---|---|---|---|---|---|---|---|
| | | 40800 - 525120 | 587 | 1.327 (1.200-1.467) | <0.001 | | | | 1.199 (1.069-1.345) | 0.002 |
| | 2-year exc. | NDMA-unexposed | 1011 | 1.000 | - | | | | 1.000 | - |
| | | < 10800 | 286 | 1.148 (1.007-1.310) | 0.040 | | | | 1.291 (1.121-1.485) | <0.001 |
| | | 10800 - 40760 | 316 | 1.125 (0.991-1.278) | 0.069 | | | | 0.984 (0.847-1.143) | 0.829 |
| | | 40800 - 525120 | 505 | 1.295 (1.161-1.443) | <0.001 | | | | 1.151 (1.016-1.304) | 0.028 |
| 1:1:1 PSM[§] | Overall | NDMA-unexposed | | | | 535 | 1.000 | - | | |
| | | < 10800 | | | | 99 | 1.107 (0.886-1.384)[¶] | 0.370 | | |
| | | 10800 - 40760 | | | | 86 | 0.985 (0.777-1.248)[¶] | 0.898 | | |
| | | 40800 - 525120 | | | | 81 | 0.968 (0.753-1.244)[¶] | 0.799 | | |
| | 6 mon exc. | NDMA-unexposed | | | | 223 | 1.000 | - | | |
| | | < 10800 | | | | 81 | 1.429 (1.105-1.848)[¶] | 0.007 | | |
| | | 10800 - 40760 | | | | 80 | 1.406 (1.085-1.821)[¶] | 0.010 | | |
| | | 40800 - 525120 | | | | 81 | 1.152 (0.885-1.498)[¶] | 0.293 | | |
| | 1-year exc. | NDMA-unexposed | | | | 203 | 1.000 | - | | |
| | | < 10800 | | | | 69 | 1.278 (0.970-1.682)[¶] | 0.081 | | |
| | | 10800 - 40760 | | | | 65 | 1.191 (0.898-1.579)[¶] | 0.224 | | |

| | | | | | | | | |
|---|---|---|---|---|---|---|---|---|
| | | 40800 - 525120 | | | | 79 | 1.117 (0.856-1.458)¶ | 0.416 |
| | 2-year exc. | NDMA-unexposed | | | | 154 | 1.000 | - |
| | | < 10800 | | | | 57 | 1.394 (1.027-1.892)¶ | 0.033 |
| | | 10800 - 40760 | | | | 51 | 1.196 (0.868-1.647)¶ | 0.274 |
| | | 40800 - 525120 | | | | 71 | 1.184 (0.890-1.576)¶ | 0.246 |

| | | | **Liver cancer** | | | **Pancreatic cancer** | | | **Thyroid cancer** | | |
|---|---|---|---|---|---|---|---|---|---|---|---|
| **PSM** | | | **Event** | **aHR (95% CI)** | ***P*** | **Event** | **aHR (95% CI)** | ***P*** | **Event** | **aHR (95% CI)** | ***P*** |
| Before PSM‡ | Overall | NDMA-unexposed | 50793 | 1.000 | - | 28007 | 1.000 | - | 23635 | 1.000 | - |
| | | < 10800 | 2300 | 1.061 (1.017-1.107) | 0.006 | 1423 | 1.075 (1.019-1.135) | 0.009 | 969 | 0.994 (0.931-1.061) | 0.853 |
| | | 10800 - 40760 | 2063 | 0.957 (0.916-1.001) | 0.054 | 1321 | 0.994 (0.940-1.051) | 0.834 | 862 | 0.913 (0.852-0.978) | 0.010 |
| | | 40800 - 525120 | 1694 | 0.874 (0.831-0.918) | <0.001 | 1146 | 0.915 (0.861-0.971) | 0.004 | 745 | 0.903 (0.839-0.973) | 0.008 |
| | 6 mon exc. | NDMA-unexposed | 28856 | 1.000 | - | 17804 | 1.000 | - | 11950 | 1.000 | - |
| | | < 10800 | 1704 | 1.223 (1.164-1.285) | <0.001 | 1062 | 1.163 (1.092-1.238) | <0.001 | 707 | 1.194 (1.105-1.289) | <0.001 |
| | | 10800 - 40760 | 1901 | 1.291 (1.232-1.353) | <0.001 | 1236 | 1.281 (1.208-1.358) | <0.001 | 785 | 1.286 (1.195-1.383) | <0.001 |
| | | 40800 - 525120 | 1694 | 1.035 (0.985-1.088) | 0.172 | 1146 | 1.043 (0.981-1.108) | 0.176 | 745 | 1.094 (1.015-1.180) | 0.019 |
| | 1-year exc. | NDMA-unexposed | 25439 | 1.000 | - | 16035 | 1.000 | - | 10512 | 1.000 | - |

| | | | | | | | | | | | |
|---|---|---|---|---|---|---|---|---|---|---|---|
| | | < 10800 | 1432 | 1.133 (1.073-1.195) | <0.001 | 894 | 1.073 (1.002-1.149) | 0.042 | 614 | 1.129 (1.040-1.226) | 0.004 |
| | | 10800 - 40760 | 1562 | 1.165 (1.106-1.227) | <0.001 | 1027 | 1.163 (1.091-1.240) | <0.001 | 642 | 1.146 (1.057-1.242) | 0.001 |
| | | 40800 - 525120 | 1663 | 1.067 (1.014-1.122) | 0.012 | 1132 | 1.079 (1.015-1.147) | 0.015 | 732 | 1.117 (1.035-1.206) | 0.004 |
| | 2-year exc. | NDMA-unexposed | 19512 | 1.000 | - | 12923 | 1.000 | - | 8247 | 1.000 | - |
| | | < 10800 | 999 | 1.061 (0.995-1.132) | 0.071 | 657 | 1.034 (0.995-1.120) | 0.404 | 452 | 1.097 (0.997-1.207) | 0.059 |
| | | 10800 - 40760 | 1010 | 0.989 (0.927-1.054) | 0.724 | 706 | 1.018 (0.943-1.099) | 0.654 | 425 | 0.974 (0.883-1.075) | 0.604 |
| | | 40800 - 525120 | 1397 | 1.038 (0.983-1.097) | 0.182 | 968 | 1.050 (0.983-1.122) | 0.147 | 619 | 1.075 (0.989-1.168) | 0.088 |

| | | | Uterine cancer† | | |
|---|---|---|---|---|---|
| **PSM** | | | **Event** | **aHR (95% CI)** | ***P*** |
| Before PSM‡ | Overall | NDMA-unexposed | 4444 | 1.000 | - |
| | | < 10800 | 231 | 1.016 (0.889-1.162) | 0.816 |
| | | 10800 - 40760 | 209 | 0.932 (0.809-1.073) | 0.325 |
| | | 40800 - 525120 | 249 | 1.189 (1.043-1.355) | 0.010 |
| | 6 mon exc. | NDMA-unexposed | 2789 | 1.000 | - |
| | | < 10800 | 175 | 1.084 (0.929-1.266) | 0.304 |
| | | 10800 - 40760 | 195 | 1.172 (1.012-1.358) | 0.035 |

| | | | | | |
|---|---|---|---|---|---|
| | | 40800 - 525120 | 249 | 1.345 (1.178-1.535) | <0.001 |
| | 1-year exc. | NDMA-unexposed | 2527 | 1.000 | - |
| | | < 10800 | 153 | 1.037 (0.879-1.223) | <0.001 |
| | | 10800 - 40760 | 159 | 1.038 (0.883-1.221) | <0.001 |
| | | 40800 - 525120 | 246 | 1.377 (1.205-1.574) | <0.001 |
| | 2-year exc. | NDMA-unexposed | 2053 | 1.000 | - |
| | | < 10800 | 103 | 0.941 (0.771-1.150) | 0.554 |
| | | 10800 - 40760 | 108 | 0.933 (0.768-1.135) | 0.490 |
| | | 40800 - 525120 | 214 | 1.356 (1.175-1.565) | <0.001 |

Abbreviations: exc., excluded; aHR, adjusted hazard ratio; mon, month; NDMA, N-Nitrosodimethylamine; PSM, propensity score matching.

A shaded area means that each model has the lowest Akaike information criterion.

†All analyses for uterine cancer were performed limited to women.

‡All hazard ratios before PSM were adjusted for age, sex, Charlson's Comorbidity Index, being a prevalent user, and the year of the first observed valsartan prescription.

§All hazard ratios after PSM were adjusted for age, sex, being a prevalent user, and the year of the first observed valsartan prescription.

¶All hazard ratios after PSM were adjusted for age, sex, and the year of the first observed valsartan prescription in the population limited to not being a prevalent user.

**Supplemental Table 9. The risks of overall, lung, and prostate cancers before and after propensity score matching according to NDMA-contaminated valsartan doses.**

| | | | All cancer | | | Lung cancer | | | Prostate cancer† | | |
|---|---|---|---|---|---|---|---|---|---|---|---|
| PSM | | | Event | aHR (95% CI) | *P* | Event | aHR (95% CI) | *P* | Event | aHR (95% CI) | *P* |
| Before PSM‡ | Overall | NDMA-unexposed | 280370 | 1.000 | | 25824 | 1.000 | | 50719 | 1.000 | |
| | | < 10800 | 16483 | 1.025 (1.009-1.042) | 0.002 | 1673 | 1.033 (0.982-1.085) | 0.210 | 2935 | 1.042 (1.003-1.082) | 0.033 |
| | | 10800 - 40760 | 14584 | 0.967 (0.951-0.983) | <0.001 | 1513 | 0.984 (0.934-1.037) | 0.553 | 2690 | 1.015 (0.976-1.056) | 0.446 |
| | | 40800 - 525120 | 12460 | 0.926 (0.909-0.943) | <0.001 | 1231 | 0.871 (0.822-0.923) | <0.001 | 2350 | 0.991 (0.950-1.034) | 0.666 |
| | 6 mon exc. | NDMA-unexposed | 133428 | 1.000 | | 13675 | 1.000 | | 23781 | 1.000 | |
| | | < 10800 | 12332 | 1.149 (1.128-1.170) | <0.001 | 1267 | 1.135 (1.071-1.203) | <0.001 | 2245 | 1.200 (1.149-1.253) | <0.001 |
| | | 10800 - 40760 | 13490 | 1.275 (1.252-1.298) | <0.001 | 1419 | 1.295 (1.226-1.369) | <0.001 | 2528 | 1.367 (1.312-1.424) | <0.001 |
| | | 40800 - 525120 | 12458 | 1.030 (1.011-1.050) | 0.002 | 1230 | 0.995 (0.938-1.055) | 0.861 | 2348 | 1.091 (1.045-1.139) | <0.001 |
| | 1-year exc. | NDMA-unexposed | 116515 | 1.000 | | 11928 | 1.000 | | 20828 | 1.000 | |
| | | < 10800 | 10525 | 1.092 (1.070-1.114) | <0.001 | 1081 | 1.080 (1.015-1.150) | 0.015 | 1918 | 1.143 (1.090-1.197) | <0.001 |
| | | 10800 - 40760 | 11034 | 1.155 (1.132-1.178) | <0.001 | 1142 | 1.155 (1.087-1.228) | <0.001 | 2102 | 1.258 (1.203-1.316) | <0.001 |
| | | 40800 - 525120 | 12226 | 1.068 (1.048-1.089) | <0.001 | 1210 | 1.037 (0.977-1.101) | 0.237 | 2298 | 1.128 (1.080-1.178) | <0.001 |
| 1:1 PSM§ | Overall | NDMA-unexposed | 31038 | 1.000 | | 2688 | 1.000 | | 6431 | 1.000 | |

| | | | | | | | | | | |
|---|---|---|---|---|---|---|---|---|---|---|
| | | < 10800 | 5496 | 1.077 (1.045-1.109) | <0.001 | 530 | 1.092 (0.991-1.202) | 0.075 | 1134 | 1.077 (1.009-1.150) | 0.027 |
| | | 10800 - 40760 | 5110 | 1.028 (0.997-1.061) | 0.077 | 519 | 1.091 (0.990-1.203) | 0.079 | 1031 | 1.001 (0.935-1.072) | 0.980 |
| | | 40800 - 525120 | 4536 | 0.992 (0.959-1.026) | 0.638 | 437 | 0.978 (0.878-1.090) | 0.687 | 963 | 1.004 (0.933-1.080) | 0.915 |
| | 6 mon exc. | NDMA-unexposed | 12193 | 1.000 | | 1292 | 1.000 | | 2634 | 1.000 | |
| | | < 10800 | 3803 | 1.248 (1.203-1.295) | <0.001 | 412 | 1.266 (1.132-1.416) | <0.001 | 829 | 1.254 (1.159-1.357) | <0.001 |
| | | 10800 - 40760 | 4332 | 1.401 (1.353-1.451) | <0.001 | 478 | 1.458 (1.311-1.622) | <0.001 | 935 | 1.378 (1.278-1.487) | <0.001 |
| | | 40800 - 525120 | 4214 | 1.147 (1.106-1.190) | <0.001 | 432 | 1.143 (1.021-1.280) | 0.020 | 929 | 1.138 (1.054-1.230) | 0.001 |
| | 1-year exc. | NDMA-unexposed | 10595 | 1.000 | | 1137 | 1.000 | | 2304 | 1.000 | |
| | | < 10800 | 3217 | 1.162 (1.117-1.209) | <0.001 | 355 | 1.185 (1.051-1.336) | 0.006 | 694 | 1.156 (1.062-1.260) | <0.001 |
| | | 10800 - 40760 | 3483 | 1.232 (1.186-1.281) | <0.001 | 371 | 1.221 (1.085-1.375) | <0.001 | 790 | 1.270 (1.170-1.378) | <0.001 |
| | | 40800 - 525120 | 4094 | 1.163 (1.120-1.207) | <0.001 | 424 | 1.156 (1.031-1.297) | 0.013 | 913 | 1.163 (1.075-1.259) | <0.001 |
| 1:1:1 PSM§ | Overall | NDMA-unexposed | 24767 | 1.000 | | 2112 | 1.000 | | 5278 | 1.000 | |
| | | < 10800 | 4417 | 1.083 (1.047-1.119) | <0.001 | 421 | 1.108 (0.995-1.235) | 0.063 | 954 | 1.091 (1.016-1.172) | 0.017 |
| | | 10800 - 40760 | 4135 | 1.040 (1.005-1.076) | 0.025 | 411 | 1.106 (0.991-1.235) | 0.071 | 871 | 1.015 (0.941-1.093) | 0.704 |
| | | 40800 - 525120 | 3686 | 1.004 (0.968-1.043) | 0.819 | 350 | 0.996 (0.883-1.125) | 0.952 | 820 | 1.015 (0.937-1.099) | 0.715 |
| | 6 mon exc. | NDMA-unexposed | 9808 | 1.000 | | 1020 | 1.000 | | 2207 | 1.000 | |

| | | | | | | | | | | |
|---|---|---|---|---|---|---|---|---|---|---|
| | < 10800 | 3078 | 1.256 (1.206-1.309) | <0.001 | 324 | 1.267 (1.117-1.438) | <0.001 | 711 | 1.279 (1.175-1.393) | <0.001 |
| | 10800 - 40760 | 3509 | 1.409 (1.355-1.466) | <0.001 | 377 | 1.463 (1.297-1.650) | <0.001 | 787 | 1.373 (1.264-1.491) | <0.001 |
| | 40800 - 525120 | 3420 | 1.154 (1.109-1.202) | <0.001 | 347 | 1.165 (1.027-1.322) | 0.018 | 788 | 1.138 (1.046-1.238) | 0.003 |
| 1-year exc. | NDMA-unexposed | 8551 | 1.000 | | 893 | 1.000 | | 1943 | 1.000 | |
| | < 10800 | 2607 | 1.167 (1.117-1.220) | <0.001 | 281 | 1.199 (1.047-1.372) | 0.009 | 597 | 1.183 (1.079-1.298) | <0.001 |
| | 10800 - 40760 | 2817 | 1.235 (1.183-1.290) | <0.001 | 290 | 1.220 (1.067-1.395) | 0.004 | 664 | 1.263 (1.155-1.380) | <0.001 |
| | 40800 - 525120 | 3330 | 1.170 (1.123-1.220) | <0.001 | 344 | 1.197 (1.053-1.360) | 0.006 | 775 | 1.164 (1.069-1.268) | <0.001 |

Abbreviations: exc., excluded; aHR, adjusted hazard ratio; mon, month; NDMA, N-Nitrosodimethylamine; PSM, propensity score matching.

A shaded area means that each model has the lowest Akaike information criterion.

†All analyses for uterine cancer were performed limited to men.

‡All hazard ratios before PSM were adjusted for age, sex, Charlson's Comorbidity Index, being a prevalent user, and the year of the first observed valsartan prescription.

§All hazard ratios after PSM were adjusted for age, sex, being a prevalent user, and the year of the first observed valsartan prescription.

**Supplemental Table 10. Subgroup analyses in patients with lung cancer according to sex and smoking status.**

| | | aHR (95% CI) | | | | | | | |
|---|---|---|---|---|---|---|---|---|---|
| | | Overall | | 6-mon | | 1-year | | 2-year | |
| | | Eventually NDMA-unexposed | NDMA-exposed | Eventually NDMA-unexposed | NDMA-exposed | Eventually NDMA-unexposed | NDMA-exposed | Eventually NDMA-unexposed | NDMA-exposed |
| **Sex** | | | | | | | | | |
| Before PSM | Male | 1.023 (0.974-1.074) | 0.993 (0.954-1.033) | 1.373 (1.304-1.447)[*] | 1.265 (1.213-1.319)[*] | 1.282 (1.213-1.355)[*] | 1.185 (1.134-1.239)[*] | 1.172 (1.099-1.251)[*] | 1.092 (1.037-1.149)[*] |
| | Female | 0.979 (0.916-1.046) | 0.964 (0.916-1.014) | 1.356 (1.263-1.455)[*] | 1.252 (1.187-1.321)[*] | 1.262 (1.170-1.362)[*] | 1.196 (1.130-1.265)[*] | 1.081 (0.987-1.183) | 1.096 (1.027-1.169)[*] |
| 1:1:1 PSM | Male | 1.007 (0.909-1.114) | 1.058 (0.962-1.164) | 1.453 (1.292-1.634)[*] | 1.519 (1.360-1.697)[*] | 1.284 (1.134-1.454)[†*] | 1.333 (1.187-1.497)[†*] | 1.123 (0.973-1.295)[†] | 1.153 (1.009-1.317)[†*] |
| | Female | 1.062 (0.919-1.228) | 1.068 (0.931-1.225) | 1.531 (1.296-1.809)[†*] | 1.514 (1.292-1.774)[†*] | 1.414 (1.184-1.688)[†*] | 1.404 (1.188-1.659)[†*] | 1.102 (0.895-1.357)[†] | 1.147 (0.948-1.389)[†] |
| 1:1 PSM | Male | - | 1.034 (0.952-1.123) | - | 1.358 (1.239-1.489)[*] | | 1.223 (1.110-1.347)[*] | - | 1.125 (1.007-1.256)[*] |
| | Female | - | 1.028 (0.919-1.150) | - | 1.299 (1.147-1.472)[*] | | 1.239 (1.087-1.411)[*] | - | 1.092 (0.942-1.265) |
| **Smoking Status** | | | | | | | | | |

| | | | | | | | | | |
|---|---|---|---|---|---|---|---|---|---|
| Before PSM | Never | 0.935 (0.844-1.036) | 1.008 (0.935-1.086) | 1.287 (1.154-1.435)* | 1.311 (1.211-1.420)* | 1.211 (1.078-1.362)* | 1.262 (1.161-1.372)* | 1.034 (0.899-1.189) | 1.174 (1.068-1.291)* |
| | Ever | 1.124 (0.970-1.303) | 1.080 (0.961-1.214) | 1.549 (1.326-1.810)* | 1.370 (1.212-1.550)* | 1.371 (1.158-1.623)* | 1.232 (1.079-1.406)* | 1.217 (0.999-1.483) | 1.137 (0.977-1.323) |
| | Current | 0.960 (0.850-1.083) | 0.929 (0.843-1.025) | 1.308 (1.151-1.487)* | 1.194 (1.078-1.322)* | 1.243 (1.086-1.424)* | 1.101 (0.988-1.228) | 1.145 (0.979-1.339) | 0.989 (0.872-1.122) |
| 1:1:1 PSM | Never | 1.008 (0.891-1.140) | 1.129 (1.008-1.264)* | 1.443 (1.251-1.665)†* | 1.617 (1.417-1.846)†* | 1.315 (1.129-1.531)†* | 1.507 (1.312-1.731)†* | 1.068 (0.894-1.276)† | 1.309 (1.118-1.533)†* |
| | Ever | 1.169 (0.979-1.397) | 1.140 (0.962-1.352) | 1.750 (1.423-2.153)* | 1.679 (1.375-2.051)* | 1.444 (1.158-1.800)* | 1.401 (1.135-1.730)* | 1.187 (0.920-1.532)† | 1.105 (0.865-1.412)† |
| | Current | 0.954 (0.824-1.104) | 0.924 (0.803-1.063) | 1.355 (1.147-1.601)* | 1.295 (1.103-1.521)* | 1.263 (1.061-1.504)†* | 1.138 (0.961-1.349)† | 1.129 (0.925-1.378)† | 0.976 (0.803-1.186)† |
| 1:1 PSM | Never | - | 1.064 (0.968-1.169) | - | 1.374 (1.236-1.527)* | - | 1.294 (1.158-1.445)* | - | 1.192 (1.052-1.351)* |
| | Ever | - | 1.102 (0.952-1.275) | - | 1.457 (1.237-1.716)* | - | 1.281 (1.078-1.523)* | - | 1.151 (0.945-1.403) |
| | Current | - | 0.931 (0.825-1.052) | - | 1.198 (1.046-1.371)* | - | 1.086 (0.942-1.252) | - | 0.960 (0.816-1.128) |
| Interaction | | **Wald $\chi^2$** | ***P*** | **Wald $\chi^2$** | ***P*** | **Wald $\chi^2$** | ***P*** | **Wald $\chi^2$** | ***P*** |
| Before PSM | Sex * NDMA | 6.226 | 0.045 | 0.731 | 0.694 | 0.281 | 0.869 | 2.180 | 0.336 |

| | | | | | | | | | |
|---|---|---|---|---|---|---|---|---|---|
| | Smoking * NDMA | 4.712 | 0.318 | 4.316 | 0.365 | 3.074 | 0.546 | 5.553 | 0.235 |
| 1:1:1 PSM | Sex * NDMA | 1.139 | 0.566 | 0.282 | 0.868 | 0.213 | 0.899 | 0.075 | 0.963 |
| | Smoking * NDMA | 3.557 | 0.469 | 6.196 | 0.185 | 7.036 | 0.134 | 9.091 | 0.059 |
| 1:1 PSM | Sex * NDMA | 1.387 | 0.239 | 0.259 | 0.611 | 0.002 | 0.965 | 0.077 | 0.781 |
| | Smoking * NDMA | 1.070 | 0.586 | 3.097 | 0.213 | 2.734 | 0.255 | 4.377 | 0.112 |

Abbreviations: aHR, adjusted hazard ratio; CI, confidence interval; NDMA, mon, month; N-Nitrosodimethylamine; PSM, propensity score matching.

*Statistically significant at 0.05.

†All hazard ratios were estimated in the population limited to not being a prevalent user.

**Supplemental Table 11. Subgroup analyses in patients with prostate cancer according to age.**

| | | aHR (95% CI) | | | | | | | |
|---|---|---|---|---|---|---|---|---|---|
| | | **Overall** | | **6-mon** | | **1-year** | | **2-year** | |
| | | **Eventually NDMA-unexposed** | **NDMA-exposed** | **Eventually NDMA-unexposed** | **NDMA-exposed** | **Eventually NDMA-unexposed** | **NDMA-exposed** | **Eventually NDMA-unexposed** | **NDMA-exposed** |
| **Age (year)** | | | | | | | | | |
| Before PSM | ≤ 65 | 0.924 (0.889-0.961)* | 1.011 (0.980-1.044) | 1.243 (1.193-1.296)* | 1.304 (1.261-1.348)* | 1.155 (1.105-1.208)* | 1.235 (1.192-1.279)* | 1.016 (0.964-1.070) | 1.132 (1.087-1.178)* |
| | > 65 | 0.970 (0.926-1.016) | 1.045 (1.010-1.081)* | 1.395 (1.328-1.465)* | 1.387 (1.338-1.439)* | 1.258 (1.193-1.326)* | 1.323 (1.273-1.375)* | 1.128 (1.060-1.200)* | 1.197 (1.145-1.251)* |
| 1:1:1 PSM | ≤ 65 | 0.955 (0.889-1.025) | 1.031 (0.965-1.101) | 1.351 (1.243-1.469)* | 1.433 (1.327-1.551)* | 1.195 (1.093-1.306)* | 1.329 (1.224-1.442)* | 1 005 (0.906-1.114) | 1.151 (1.048-1.264)* |
| | > 65 | 0.945 (0.863-1.035) | 1.074 (0.989-1.168) | 1.431 (1.281-1.598)* | 1.555 (1.403-1.724)* | 1.238 (1.101-1.393)* | 1.375 (1.234-1.533)* | 0.971 (0.846-1.113) | 1.107 (0.978-1.254) |
| 1:1 PSM | ≤ 65 | - | 1.016 (0.957-1.078) | - | 1.288 (1.204-1.378)* | - | 1.240 (1.155-1.331)* | - | 1.113 (1.027-1.206)* |
| | > 65 | - | 1.039 (0.969-1.114) | - | 1.385 (1.278-1.502)* | - | 1.284 (1.179-1.398)* | - | 1.093 (0.992-1.204) |
| Interaction | | **Wald $\chi^2$** | ***P*** | **Wald $\chi^2$** | ***P*** | **Wald $\chi^2$** | ***P*** | **Wald $\chi^2$** | ***P*** |

| | | | | | | | | | |
|---|---|---|---|---|---|---|---|---|---|
| Before PSM | Age * NDMA | 33.730 | <0.001 | 2.665 | 0.264 | 0.547 | 0.761 | 2.678 | 0.262 |
| 1:1:1 PSM | Age * NDMA | 14.306 | 0.001 | 0.281 | 0.869 | 0.016 | 0.992 | 0.235 | 0.889 |
| 1:1 PSM | Age * NDMA | 7.564 | 0.006 | 0.013 | 0.909 | 0.016 | 0.899 | 0.098 | 0.755 |

Abbreviations: aHR, adjusted hazard ratio; CI, confidence interval; mon, month; NDMA, N-Nitrosodimethylamine; PSM, propensity score matching.

*Statistically significant at 0.05.

†All hazard ratios were estimated in the population limited to not being a prevalent user.

**Supplemental Table 12. The risks of all cancer and organ-specific cancers before and after PSM in study population excluding patients with any cancer during the first year of follow-up.**

| PSM | | Cancer | All cancer | | | Lung cancer | | | Prostate cancer† | | |
|---|---|---|---|---|---|---|---|---|---|---|---|
| | | | Event | aHR (95% CI) | *P* | Event | aHR (95% CI) | *P* | Event | aHR (95% CI) | *P* |
| Before PSM‡ | Overall | NDMA-unexposed | 354043 | 1.000 | | 33971 | 1.000 | | 64878 | 1.000 | |
| | | Eventually NDMA-unexposed | 27554 | 0.962 (0.950-0.974) | <0.001 | 2824 | 0.983 (0.945-1.023) | 0.405 | 4934 | 0.937 (0.910-0.966) | <0.001 |
| | | NDMA-exposed | 45593 | 0.975 (0.965-0.985) | <0.001 | 4651 | 0.967 (0.937-0.998) | 0.039 | 8440 | 1.018 (0.994-1.042) | 0.147 |
| | 6 mon exc. | NDMA-unexposed | 218994 | 1.000 | | 22331 | 1.000 | | 40088 | 1.000 | |
| | | Eventually NDMA-unexposed | 25708 | 1.312 (1.294-1.330) | <0.001 | 2643 | 1.323 (1.269-1.380) | <0.001 | 4618 | 1.279 (1.239-1.320) | <0.001 |
| | | NDMA-exposed | 43373 | 1.251 (1.237-1.264) | <0.001 | 4451 | 1.232 (1.192-1.274) | <0.001 | 8080 | 1.314 (1.282-1.347) | <0.001 |
| | 1-year exc. | NDMA-unexposed | 201123 | 1.000 | | 20495 | 1.000 | | 36959 | 1.000 | |
| | | Eventually NDMA-unexposed | 22181 | 1.241 (1.223-1.259) | <0.001 | 2296 | 1.264 (1.208-1.322) | <0.001 | 3965 | 1.201 (1.161-1.242) | <0.001 |
| | | NDMA-exposed | 38897 | 1.206 (1.193-1.220) | <0.001 | 3970 | 1.184 (1.143-1.226) | <0.001 | 7281 | 1.271 (1.239-1.305) | <0.001 |
| | 2-year exc. | NDMA-unexposed | 158135 | | | 16122 | | | 29353 | | |
| | | Eventually NDMA-unexposed | 15906 | 1.097 (1.079-1.116) | <0.001 | 1655 | 1.130 (1.072-1.191) | <0.001 | 2855 | 1.062 (1.021-1.106) | 0.003 |

| | | | | | | | | | | |
|---|---|---|---|---|---|---|---|---|---|---|
| | | NDMA-exposed | 29128 | 1.098 (1.084-1.113 | <0.001 | 2991 | 1.088 (1.046-1.133) | <0.001 | 5455 | 1.156 (1.122-1.191) | <0.001 |
| 1:1 PSM§ | Overall | NDMA-unexposed | 28674 | 1.000 | | 2608 | 1.000 | | 5818 | 1.000 | |
| | | NDMA-exposed | 15706 | 1.011 (0.990-1.033) | 0.301 | 1536 | 1.023 (0.956-1.095) | 0.514 | 3243 | 1.016 (0.970-1.065) | 0.495 |
| | 6 mon exc. | NDMA-unexposed | 13993 | 1.000 | | 1449 | 1.000 | | 2934 | 1.000 | |
| | | NDMA-exposed | 14159 | 1.309 (1.278-1.341) | <0.001 | 1463 | 1.317 (1.222-1.420) | <0.001 | 3018 | 1.304 (1.237-1.374) | <0.001 |
| | 1-year exc. | NDMA-unexposed | 12422 | 1.000 | | 1304 | 1.000 | | 2627 | 1.000 | |
| | | NDMA-exposed | 12606 | 1.239 (1.208-1.271) | <0.001 | 1295 | 1.220 (1.128-1.320) | <0.001 | 2732 | 1.253 (1.186-1.323) | <0.001 |
| | 2-year exc. | NDMA-unexposed | 9323 | 1.000 | | 999 | 1.000 | | 2043 | 1.000 | |
| | | NDMA-exposed | 9339 | 1.120 (1.088-1.153) | <0.001 | 976 | 1.105 (1.010-1.208) | 0.029 | 2023 | 1.101 (1.035-1.172) | 0.002 |
| 1:1:1 PSM§ | Overall | NDMA-unexposed | 27100 | 1.000 | | 2324 | 1.000 | | 5715 | 1.000 | |
| | | Eventually NDMA-unexposed | 8919 | 1.014 (0.988-1.041) | 0.281 | 844 | 1.026 (0.942-1.117) | 0.560 | 1791 | 0.939 (0.886-0.994) | 0.031 |
| | | NDMA-exposed | 11022 | 1.031 (1.006-1.057) | 0.014 | 1047 | 1.049 (0.968-1.137) | 0.247 | 2393 | 1.039 (0.985-1.096) | 0.157 |
| | 6 mon exc. | NDMA-unexposed | 9006 | 1.000 | | 973 | 1.000 | | 2069 | 1.000 | |
| | | Eventually NDMA-unexposed | 7357 | 1.491 (1.444-1.540) | <0.001 | 780 | 1.475 (1.337-1.626) | <0.001 | 1606 | 1.374 (1.284-1.470) | <0.001 |
| | | NDMA-exposed | 9150 | 1.490 (1.445-1.536) | <0.001 | 985 | 1.492 (1.360-1.638) | <0.001 | 2139 | 1.470 (1.380-1.566) | <0.001 |

| | | | | | | | | | | | |
|---|---|---|---|---|---|---|---|---|---|---|---|
| | 1-year exc. | NDMA-unexposed | 7805 | 1.000 | | 855 | 1.000 | | 1824 | 1.000 | |
| | | Eventually NDMA-unexposed | 6165 | 1.309 (1.265-1.355) | <0.001 | 675 | 1.324 (1.194-1.469) | <0.001 | 1358 | 1.207 (1.123-1.297) | <0.001 |
| | | NDMA-exposed | 7911 | 1.333 (1.290-1.376) | <0.001 | 863 | 1.339 (1.215-1.477) | <0.001 | 1898 | 1.342 (1.256-1.434) | <0.001 |
| | 2-year exc. | NDMA-unexposed | 5630 | 1.000 | | 643 | 1.000 | | 1378 | 1.000 | |
| | | Eventually NDMA-unexposed | 4230 | 1.098 (1.054-1.143) | <0.001 | 475 | 1.099 (0.974-1.239) | 0.125 | 944 | 0.990 (0.911-1.077) | 0.819 |
| | | NDMA-exposed | 5603 | 1.142 (1.100-1.186) | <0.001 | 627 | 1.134 (1.014-1.267) | 0.028 | 1373 | 1.135 (1.052-1.224) | 0.001 |

Abbreviations: aHR, adjusted hazard ratio; CI, confidence interval; exc., excluded; mon, month; NDMA, N-Nitrosodimethylamine; PSM, propensity score matching.

A shaded area means that each model has the lowest Akaike information criterion.

†All analyses for uterine cancer were performed limited to men.

‡All hazard ratios before PSM were adjusted for age, sex, Charlson's Comorbidity Index, being a prevalent user, and the year of the first observed valsartan prescription.

§All hazard ratios after PSM were adjusted for age, sex, being a prevalent user, and the year of the first observed valsartan prescription.

**Supplemental Table 13. The risks of all cancer and organ-specific cancers before and after PSM in valsartan new users excluding patients with any cancer during the first year of follow-up.**

| | | Cancer | All cancer | | | Lung cancer | | | Prostate cancer† | | |
|---|---|---|---|---|---|---|---|---|---|---|---|
| PSM | | | Event | aHR (95% CI) | *P* | Event | aHR (95% CI) | *P* | Event | aHR (95% CI) | *P* |
| Before PSM‡ | Overall | NDMA-unexposed | 26557 6 | 1.000 | | 25050 | 1.000 | | 48017 | 1.000 | |
| | | Eventually NDMA-unexposed | 27550 | 0.950 (0.938-0.962) | <0.001 | 2823 | 0.962 (0.925-1.001) | 0.058 | 4934 | 0.931 (0.903-0.959) | <0.001 |
| | | NDMA-exposed | 40381 | 0.961 (0.951-0.971) | <0.001 | 4102 | 0.944 (0.913-0.977) | 0.001 | 7456 | 1.009 (0.984-1.035) | 0.492 |
| | 6 mon exc. | NDMA-unexposed | 13272 9 | 1.000 | | 13610 | 1.000 | | 23660 | 1.000 | |
| | | Eventually NDMA-unexposed | 25704 | 1.231 (1.214-1.248) | <0.001 | 2642 | 1.245 (1.194-1.299) | <0.001 | 4618 | 1.202 (1.164-1.241) | <0.001 |
| | | NDMA-exposed | 38179 | 1.215 (1.201-1.229) | <0.001 | 3905 | 1.200 (1.157-1.244) | <0.001 | 7100 | 1.281 (1.247-1.316) | <0.001 |
| | 1-year exc. | NDMA-unexposed | 11651 5 | 1.000 | | 11928 | 1.000 | | 20828 | 1.000 | |
| | | Eventually NDMA-unexposed | 22177 | 1.153 (1.136-1.170) | <0.001 | 2295 | 1.176 (1.125-1.231) | <0.001 | 3965 | 1.119 (1.081-1.158) | <0.001 |
| | | NDMA-exposed | 33785 | 1.156 (1.142-1.170) | <0.001 | 3433 | 1.136 (1.094-1.181) | <0.001 | 6318 | 1.225 (1.191-1.260) | <0.001 |
| | 2-year exc. | NDMA-unexposed | 86363 | 1.000 | | 8838 | 1.000 | | 15521 | 1.000 | |
| | | Eventually NDMA-unexposed | 15902 | 1.049 (1.031-1.067) | <0.001 | 1654 | 1.079 (1.024-1.138) | 0.005 | 2855 | 1.019 (0.979-1.060) | 0.365 |

| | | | | | | | | | | | |
|---|---|---|---|---|---|---|---|---|---|---|---|
| | | NDMA-exposed | 24828 | 1.069 (1.054-1.085) | <0.001 | 2530 | 1.055 (1.009-1.103) | 0.018 | 4638 | 1.132 (1.095-1.170) | <0.001 |
| 1:1 PSM[§] | Overall | NDMA-unexposed | 26718 | 1.000 | | 2415 | 1.000 | | 5444 | 1.000 | |
| | | NDMA-exposed | 13854 | 1.010 (0.988-1.033) | 0.363 | 1354 | 1.020 (0.949-1.097) | 0.589 | 2868 | 1.010 (0.962-1.062) | 0.681 |
| | 6 mon exc. | NDMA-unexposed | 12134 | 1.000 | | 1260 | 1.000 | | 2580 | 1.000 | |
| | | NDMA-exposed | 12316 | 1.317 (1.283-1.352) | <0.001 | 1283 | 1.333 (1.230-1.445) | <0.001 | 2645 | 1.300 (1.229-1.375) | <0.001 |
| | 1-year exc. | NDMA-unexposed | 10595 | 1.000 | | 1115 | 1.000 | | 2276 | 1.000 | |
| | | NDMA-exposed | 10793 | 1.231 (1.198-1.265) | <0.001 | 1119 | 1.224 (1.124-1.332) | <0.001 | 2364 | 1.238 (1.168-1.313) | <0.001 |
| | 2-year exc. | NDMA-unexposed | 7802 | 1.000 | | 838 | | | 1737 | | |
| | | NDMA-exposed | 7855 | 1.119 (1.084-1.155) | <0.001 | 821 | 1.099 (0.997-1.211) | 0.059 | 1718 | 1.095 (1.023-1.171) | 0.009 |
| 1:1:1 PSM[§] | Overall | NDMA-unexposed | 27085 | 1.000 | | 2323 | 1.000 | | 5712 | 1.000 | |
| | | Eventually NDMA-unexposed | 8914 | 1.015 (0.989-1.042) | 0.273 | 844 | 1.026 (0.942-1.117) | 0.556 | 1791 | 0.940 (0.887-0.995) | 0.034 |
| | | NDMA-exposed | 11017 | 1.032 (1.007-1.057) | 0.013 | 1047 | 1.049 (0.967-1.137) | 0.248 | 2393 | 1.040 (0.987-1.097) | 0.144 |
| | 6 mon exc. | NDMA-unexposed | 8999 | 1.000 | | 973 | 1.000 | | 2066 | 1.000 | |
| | | Eventually NDMA-unexposed | 7354 | 1.492 (1.445-1.540) | <0.001 | 780 | 1.475 (1.337-1.626) | <0.001 | 1606 | 1.376 (1.286-1.472) | <0.001 |
| | | NDMA-exposed | 9145 | 1.490 (1.445-1.537) | <0.001 | 985 | 1.492 (1.360-1.637) | <0.001 | 2139 | 1.473 (1.382-1.569) | <0.001 |

| | | | | | | | | | | |
|---|---|---|---|---|---|---|---|---|---|---|
| 1-year exc. | NDMA-unexposed | 7798 | 1.000 | | 855 | 1.000 | | 1821 | 1.000 | |
| | Eventually NDMA-unexposed | 6162 | 1.310 (1.265-1.355) | <0.001 | 675 | 1.324 (1.194-1.469) | <0.001 | 1358 | 1.209 (1.125-1.299) | <0.001 |
| | NDMA-exposed | 7906 | 1.333 (1.291-1.377) | <0.001 | 863 | 1.339 (1.215-1.476) | <0.001 | 1898 | 1.345 (1.258-1.437) | <0.001 |
| 2-year exc. | NDMA-unexposed | 5623 | 1.000 | | 643 | 1.000 | | 1375 | 1.000 | |
| | Eventually NDMA-unexposed | 4227 | 1.098 (1.055-1.144) | <0.001 | 475 | 1.099 (0.974-1.239) | 0.125 | 944 | 0.992 (0.913-1.079) | 0.857 |
| | NDMA-exposed | 5598 | 1.142 (1.100-1.186) | <0.001 | 627 | 1.133 (1.014-1.267) | 0.028 | 1373 | 1.138 (1.055-1.227) | 0.001 |

Abbreviations: aHR, adjusted hazard ratio; CI, confidence interval; exc., excluded; mon, month; NDMA, N-Nitrosodimethylamine; PSM, propensity score matching.

A shaded area means that each model has the lowest Akaike information criterion.

†All analyses for uterine cancer were performed limited to men.

‡All hazard ratios before PSM were adjusted for age, sex, Charlson's Comorbidity Index, and the year of the first observed valsartan prescription.

§All hazard ratios after PSM were adjusted for age, sex, and the year of the first observed valsartan prescription.

**Supplemental Table 14. The risks of all cancer and organ-specific cancers before and after PSM in study population excluding patients with any cancer during the first year of follow-up.**

| PSM | | | All cancer | | | Lung cancer | | | Prostate cancer† | | |
|---|---|---|---|---|---|---|---|---|---|---|---|
| | | | Event | aHR (95% CI) | *P* | Event | aHR (95% CI) | *P* | Event | aHR (95% CI) | *P* |
| Before PSM‡ | Overall | NDMA-unexposed | 333150 | 1.000 | | 32258 | 1.000 | | 60963 | 1.000 | |
| | | < 10800 | 17087 | 1.026 (1.010-1.042) | 0.002 | 1742 | 1.034 (0.985-1.086) | 0.174 | 3071 | 1.048 (1.010-1.087) | 0.013 |
| | | 10800 - 40760 | 16437 | 0.978 (0.963-0.994) | 0.006 | 1722 | 1.003 (0.955-1.054) | 0.900 | 3025 | 1.014 (0.977-1.052) | 0.472 |
| | | 40800 - 525120 | 14738 | 0.942 (0.926-0.958) | <0.001 | 1457 | 0.894 (0.847-0.943) | <0.001 | 2782 | 0.999 (0.961-1.038) | 0.945 |
| | 6 mon exc. | NDMA-unexposed | 218994 | 1.000 | | 22331 | 1.000 | | 40088 | 1.000 | |
| | | < 10800 | 13253 | 1.173 (1.152-1.194) | <0.001 | 1364 | 1.155 (1.093-1.221) | <0.001 | 2433 | 1.231 (1.181-1.283) | <0.001 |
| | | 10800 - 40760 | 15384 | 1.284 (1.263-1.306) | <0.001 | 1631 | 1.302 (1.237-1.370) | <0.001 | 2867 | 1.352 (1.301-1.405) | <0.001 |
| | | 40800 - 525120 | 14736 | 1.086 (1.068-1.105) | <0.001 | 1456 | 1.041 (0.986-1.098) | 0.146 | 2780 | 1.145 (1.101-1.190) | <0.001 |
| | 1-year exc. | NDMA-unexposed | 201123 | 1.000 | | 20495 | 1.000 | | 36959 | 1.000 | |
| | | < 10800 | 11467 | 1.126 (1.105-1.148) | <0.001 | 1180 | 1.111 (1.047-1.179) | 0.001 | 2107 | 1.182 (1.131-1.236) | <0.001 |
| | | 10800 - 40760 | 12927 | 1.186 (1.165-1.208) | <0.001 | 1354 | 1.188 (1.124-1.256) | <0.001 | 2445 | 1.265 (1.214-1.319) | <0.001 |
| | | 40800 - 525120 | 14503 | 1.133 (1.114-1.153) | <0.001 | 1436 | 1.091 (1.033-1.152) | 0.002 | 2729 | 1.190 (1.143-1.237) | <0.001 |
| | 2-year exc. | NDMA-unexposed | 158135 | 1.000 | | 16122 | 1.000 | | 29353 | 1.000 | |

| | | | | | | | | | | | |
|---|---|---|---|---|---|---|---|---|---|---|---|
| | | < 10800 | 8111 | 1.057 (1.034-1.081) | <0.001 | 831 | 1.042 (0.971-1.119) | 0.250 | 1494 | 1.111 (1.054-1.171) | <0.001 |
| | | 10800 - 40760 | 8658 | 1.030 (1.008-1.053) | 0.008 | 937 | 1.066 (0.998-1.140) | 0.059 | 1631 | 1.091 (1.038-1.148) | 0.001 |
| | | 40800 - 525120 | 12359 | 1.108 (1.088-1.129) | <0.001 | 1223 | 1.071 (1.009-1.136) | 0.023 | 2330 | 1.165 (1.117-1.216) | <0.001 |
| 1:1 PSM[§] | Overall | NDMA-unexposed | 27826 | 1.000 | | 2528 | 1.000 | | 5668 | 1.000 | |
| | | < 10800 | 5600 | 1.066 (1.035-1.098) | <0.001 | 535 | 1.068 (0.971-1.175) | 0.174 | 1155 | 1.076 (1.008-1.148) | 0.027 |
| | | 10800 - 40760 | 5645 | 1.026 (0.996-1.057) | 0.089 | 572 | 1.083 (0.987-1.189) | 0.093 | 1135 | 0.998 (0.935-1.066) | 0.953 |
| | | 40800 - 525120 | 5309 | 0.991 (0.961-1.023) | 0.576 | 509 | 0.978 (0.884-1.082) | 0.673 | 1103 | 0.991 (0.926-1.062) | 0.808 |
| | 6 mon exc. | NDMA-unexposed | 13993 | 1.000 | | 1449 | 1.000 | | 2934 | 1.000 | |
| | | < 10800 | 4124 | 1.245 (1.203-1.290) | <0.001 | 427 | 1.236 (1.109-1.379) | <0.001 | 892 | 1.277 (1.184-1.377) | <0.001 |
| | | 10800 - 40760 | 4972 | 1.377 (1.333-1.423) | <0.001 | 531 | 1.418 (1.282-1.569) | <0.001 | 1048 | 1.353 (1.260-1.453) | <0.001 |
| | | 40800 - 525120 | 5063 | 1.149 (1.112-1.188) | <0.001 | 505 | 1.136 (1.023-1.262) | 0.017 | 1078 | 1.137 (1.058-1.222) | 0.001 |
| | 1-year exc. | NDMA-unexposed | 12422 | 1.000 | | 1304 | 1.000 | | 2627 | 1.000 | |
| | | < 10800 | 3543 | 1.176 (1.132-1.221) | <0.001 | 370 | 1.160 (1.032-1.303) | 0.013 | 764 | 1.202 (1.108-1.303) | <0.001 |
| | | 10800 - 40760 | 4121 | 1.241 (1.198-1.286) | <0.001 | 428 | 1.223 (1.095-1.366) | <0.001 | 906 | 1.268 (1.175-1.368) | <0.001 |
| | | 40800 - 525120 | 4942 | 1.169 (1.130-1.209) | <0.001 | 497 | 1.148 (1.033-1.277) | 0.011 | 1062 | 1.165 (1.083-1.254) | <0.001 |
| | 2-year exc. | NDMA-unexposed | 9323 | 1.000 | | 999 | 1.000 | | 2043 | 1.000 | |

| | | | | | | | | | | |
|---|---|---|---|---|---|---|---|---|---|---|
| | | < 10800 | 2492 | 1.095 (1.048-1.145) | <0.001 | 254 | 1.045 (0.910-1.200) | 0.532 | 543 | 1.102 (1.002-1.212) | 0.045 |
| | | 10800 - 40760 | 2698 | 1.051 (1.006-1.097) | 0.025 | 295 | 1.066 (0.935-1.215) | 0.341 | 587 | 1.036 (0.945-1.136) | 0.452 |
| | | 40800 - 525120 | 4149 | 1.129 (1.087-1.171) | <0.001 | 427 | 1.115 (0.994-1.251) | 0.063 | 863 | 1.095 (1.011-1.186) | 0.025 |
| 1:1:1 PSM§ | Overall | NDMA-unexposed | 20701 | 1.000 | | 1831 | 1.000 | | 4388 | 1.000 | |
| | | < 10800 | 4236 | 1.075 (1.039-1.112) | <0.001 | 402 | 1.093 (0.978-1.221) | 0.117 | 910 | 1.080 (1.003-1.162) | 0.041 |
| | | 10800 - 40760 | 4044 | 1.039 (1.003-1.076) | 0.033 | 394 | 1.080 (0.966-1.209) | 0.177 | 849 | 1.008 (0.934-1.088) | 0.834 |
| | | 40800 - 525120 | 3618 | 0.997 (0.960-1.036) | 0.885 | 344 | 0.994 (0.880-1.124) | 0.927 | 797 | 0.991 (0.915-1.075) | 0.835 |
| | 6 mon exc. | NDMA-unexposed | 9774 | 1.000 | | 995 | 1.000 | | 2167 | 1.000 | |
| | | < 10800 | 3070 | 1.250 (1.200-1.303) | <0.001 | 313 | 1.249 (1.098-1.420) | 0.001 | 704 | 1.286 (1.181-1.402) | <0.001 |
| | | 10800 - 40760 | 3498 | 1.401 (1.347-1.457) | <0.001 | 362 | 1.427 (1.263-1.612) | <0.001 | 772 | 1.360 (1.251-1.478) | <0.001 |
| | | 40800 - 525120 | 3419 | 1.150 (1.105-1.198) | <0.001 | 341 | 1.157 (1.019-1.314) | 0.025 | 774 | 1.123 (1.031-1.222) | 0.008 |
| | 1-year exc. | NDMA-unexposed | 8558 | 1.000 | | 878 | 1.000 | | 1922 | 1.000 | |
| | | < 10800 | 2609 | 1.167 (1.117-1.220) | <0.001 | 270 | 1.175 (1.024-1.348) | 0.022 | 595 | 1.198 (1.092-1.314) | <0.001 |
| | | 10800 - 40760 | 2819 | 1.234 (1.182-1.289) | <0.001 | 282 | 1.206 (1.053-1.381) | 0.007 | 654 | 1.258 (1.150-1.376) | <0.001 |
| | | 40800 - 525120 | 3330 | 1.169 (1.122-1.218) | <0.001 | 338 | 1.189 (1.045-1.353) | 0.009 | 761 | 1.151 (1.056-1.254) | 0.001 |
| | 2-year exc. | NDMA-unexposed | 6291 | 1.000 | | 662 | 1.000 | | 1468 | 1.000 | |

| | | | | | | | | | |
|---|---|---|---|---|---|---|---|---|---|
| < 10800 | 1806 | 1.102 (1.046-1.162) | <0.001 | 180 | 1.058 (0.897-1.249) | 0.503 | 413 | 1.100 (0.986-1.227) | 0.089 |
| 10800 - 40760 | 1771 | 1.035 (0.982-1.092) | 0.200 | 180 | 1.008 (0.853-1.190) | 0.929 | 413 | 1.033 (0.925-1.154) | 0.563 |
| 40800 - 525120 | 2770 | 1.138 (1.087-1.191) | <0.001 | 287 | 1.161 (1.008-1.337) | 0.038 | 636 | 1.094 (0.995-1.202) | 0.062 |

Abbreviations: exc., excluded; aHR, adjusted hazard ratio; mon, month; NDMA, N-Nitrosodimethylamine; PSM, propensity score matching.

A shaded area means that each model has the lowest Akaike information criterion.

[†]All analyses for uterine cancer were performed limited to men.

[‡]All hazard ratios before PSM were adjusted for age, sex, Charlson's Comorbidity Index, being a prevalent user, and the year of the first observed valsartan prescription.

[§]All hazard ratios after PSM were adjusted for age, sex, being a prevalent user, and the year of the first observed valsartan prescription.

**Supplemental Table 15. The risks of all cancer and organ-specific cancers before and after PSM in valsartan new users excluding patients with any cancer during the first year of follow-up.**

| PSM | | | All cancer | | | Lung cancer | | | Prostate cancer[†] | | |
|---|---|---|---|---|---|---|---|---|---|---|---|
| | | | Event | aHR (95% CI) | *P* | Event | aHR (95% CI) | *P* | Event | aHR (95% CI) | *P* |
| Before PSM[‡] | Overall | NDMA-unexposed | 24470 4 | 1.000 | | 23338 | 1.000 | | 44106 | 1.000 | |
| | | < 10800 | 16094 | 1.019 (1.003-1.035) | 0.024 | 1639 | 1.022 (0.972-1.075) | 0.388 | 2868 | 1.036 (0.998-1.077) | 0.067 |
| | | 10800 - 40760 | 14476 | 0.964 (0.948-0.980) | <0.001 | 1501 | 0.977 (0.927-1.030) | 0.387 | 2672 | 1.013 (0.973-1.053) | 0.537 |
| | | 40800 - 525120 | 12460 | 0.924 (0.907-0.941) | <0.001 | 1231 | 0.869 (0.819-0.921) | <0.001 | 2350 | 0.986 (0.945-1.029) | 0.529 |
| | 6 mon exc. | NDMA-unexposed | 13272 9 | 1.000 | | 13610 | 1.000 | | 23660 | 1.000 | |
| | | < 10800 | 12288 | 1.144 (1.123-1.165) | <0.001 | 1263 | 1.130 (1.066-1.197) | <0.001 | 2238 | 1.194 (1.143-1.247) | <0.001 |
| | | 10800 - 40760 | 13433 | 1.268 (1.245-1.291) | <0.001 | 1412 | 1.285 (1.216-1.357) | <0.001 | 2514 | 1.354 (1.299-1.411) | <0.001 |
| | | 40800 - 525120 | 12458 | 1.029 (1.010-1.048) | 0.003 | 1230 | 0.990 (0.933-1.050) | 0.736 | 2348 | 1.085 (1.040-1.133) | <0.001 |
| | 1-year exc. | NDMA-unexposed | 11651 5 | 1.000 | | 11928 | 1.000 | | 20828 | 1.000 | |
| | | < 10800 | 10525 | 1.091 (1.070-1.114) | <0.001 | 1081 | 1.079 (1.013-1.148) | 0.017 | 1918 | 1.142 (1.089-1.196) | <0.001 |
| | | 10800 - 40760 | 11034 | 1.155 (1.132-1.178) | <0.001 | 1142 | 1.152 (1.084-1.225) | <0.001 | 2102 | 1.255 (1.199-1.313) | <0.001 |
| | | 40800 - 525120 | 12226 | 1.068 (1.048-1.089) | <0.001 | 1210 | 1.033 (0.973-1.096) | 0.290 | 2298 | 1.123 (1.075-1.173) | <0.001 |
| | 2-year exc. | NDMA-unexposed | 86363 | 1.000 | | 8838 | 1.000 | | 15521 | 1.000 | |

| | | | | | | | | | | | |
|---|---|---|---|---|---|---|---|---|---|---|---|
| | | < 10800 | 7333 | 1.049 (1.024-1.074) | <0.001 | 747 | 1.029 (0.955-1.109) | 0.450 | 1336 | 1.096 (1.036-1.159) | 0.001 |
| | | 10800 - 40760 | 7206 | 1.018 (0.994-1.043) | 0.148 | 769 | 1.046 (0.972-1.127) | 0.230 | 1365 | 1.101 (1.041-1.164) | 0.001 |
| | | 40800 - 525120 | 10289 | 1.063 (1.041-1.085) | <0.001 | 1014 | 1.026 (0.961-1.096) | 0.435 | 1937 | 1.121 (1.069-1.176) | <0.001 |
| 1:1 PSM§ | Overall | NDMA-unexposed | 25877 | 1.000 | | 2335 | 1.000 | | 5297 | 1.000 | |
| | | < 10800 | 5255 | 1.067 (1.035-1.100) | <0.001 | 505 | 1.076 (0.974-1.187) | 0.149 | 1076 | 1.062 (0.993-1.137) | 0.079 |
| | | 10800 - 40760 | 4983 | 1.027 (0.995-1.060) | 0.095 | 501 | 1.078 (0.976-1.191) | 0.139 | 1000 | 0.994 (0.927-1.066) | 0.869 |
| | | 40800 - 525120 | 4457 | 0.989 (0.956-1.023) | 0.507 | 428 | 0.975 (0.874-1.088) | 0.656 | 939 | 0.988 (0.917-1.064) | 0.741 |
| | 6 mon exc. | NDMA-unexposed | 12134 | 1.000 | | 1260 | 1.000 | | 2580 | 1.000 | |
| | | < 10800 | 3788 | 1.242 (1.197-1.289) | <0.001 | 397 | 1.247 (1.112-1.397) | <0.001 | 817 | 1.254 (1.159-1.358) | <0.001 |
| | | 10800 - 40760 | 4315 | 1.393 (1.345-1.443) | <0.001 | 462 | 1.438 (1.291-1.603) | <0.001 | 914 | 1.361 (1.261-1.470) | <0.001 |
| | | 40800 - 525120 | 4213 | 1.145 (1.104-1.187) | <0.001 | 424 | 1.139 (1.016-1.277) | 0.025 | 914 | 1.126 (1.042-1.218) | 0.003 |
| | 1-year exc. | NDMA-unexposed | 10595 | 1.000 | | 1115 | 1.000 | | 2276 | 1.000 | |
| | | < 10800 | 3216 | 1.161 (1.116-1.209) | <0.001 | 340 | 1.161 (1.027-1.312) | 0.017 | 690 | 1.169 (1.073-1.273) | <0.001 |
| | | 10800 - 40760 | 3483 | 1.232 (1.186-1.281) | <0.001 | 363 | 1.221 (1.083-1.376) | 0.001 | 776 | 1.264 (1.164-1.373) | <0.001 |
| | | 40800 - 525120 | 4094 | 1.163 (1.120-1.207) | <0.001 | 416 | 1.153 (1.027-1.295) | 0.016 | 898 | 1.154 (1.066-1.250) | <0.001 |
| | 2-year exc. | NDMA-unexposed | 7802 | 1.000 | | 838 | 1.000 | | 1737 | 1.000 | |

| | | | | | | | | | | | |
|---|---|---|---|---|---|---|---|---|---|---|---|
| | | < 10800 | 2225 | 1.093 (1.042-1.146) | <0.001 | 226 | 1.034 (0.892-1.199) | 0.655 | 480 | 1.076 (0.972-1.191) | 0.158 |
| | | 10800 - 40760 | 2218 | 1.045 (0.996-1.096) | 0.070 | 242 | 1.055 (0.913-1.219) | 0.467 | 491 | 1.043 (0.943-1.154) | 0.413 |
| | | 40800 - 525120 | 3412 | 1.132 (1.087-1.179) | <0.001 | 353 | 1.119 (0.986-1.269) | 0.082 | 747 | 1.091 (1.000-1.191) | 0.049 |
| 1:1:1 PSM§ | Overall | NDMA-unexposed | 20691 | 1.000 | | 1830 | 1.000 | | 4385 | 1.000 | |
| | | < 10800 | 4233 | 1.074 (1.038-1.112) | <0.001 | 402 | 1.093 (0.978-1.221) | 0.117 | 910 | 1.081 (1.004-1.163) | 0.038 |
| | | 10800 - 40760 | 4042 | 1.039 (1.004-1.076) | 0.031 | 394 | 1.080 (0.966-1.209) | 0.177 | 849 | 1.010 (0.936-1.089) | 0.803 |
| | | 40800 - 525120 | 3618 | 0.998 (0.961-1.037) | 0.926 | 344 | 0.994 (0.880-1.124) | 0.927 | 797 | 0.993 (0.916-1.077) | 0.870 |
| | 6 mon exc. | NDMA-unexposed | 9766 | 1.000 | | 995 | 1.000 | | 2164 | 1.000 | |
| | | < 10800 | 3067 | 1.250 (1.200-1.302) | <0.001 | 313 | 1.248 (1.098-1.419) | 0.001 | 704 | 1.288 (1.182-1.403) | <0.001 |
| | | 10800 - 40760 | 3496 | 1.402 (1.348-1.458) | <0.001 | 362 | 1.426 (1.262-1.612) | <0.001 | 772 | 1.363 (1.254-1.481) | <0.001 |
| | | 40800 - 525120 | 3419 | 1.152 (1.106-1.199) | <0.001 | 341 | 1.157 (1.018-1.314) | 0.025 | 774 | 1.125 (1.033-1.224) | 0.007 |
| | 1-year exc. | NDMA-unexposed | 8551 | 1.000 | | 878 | 1.000 | | 1919 | 1.000 | |
| | | < 10800 | 2606 | 1.167 (1.116-1.219) | <0.001 | 270 | 1.175 (1.024-1.348) | 0.022 | 595 | 1.200 (1.094-1.316) | <0.001 |
| | | 10800 - 40760 | 2817 | 1.235 (1.183-1.289) | <0.001 | 282 | 1.205 (1.052-1.381) | 0.007 | 654 | 1.261 (1.152-1.379) | <0.001 |
| | | 40800 - 525120 | 3330 | 1.170 (1.123-1.220) | <0.001 | 338 | 1.188 (1.044-1.352) | 0.009 | 761 | 1.153 (1.058-1.257) | 0.001 |
| | 2-year exc. | NDMA-unexposed | 6284 | 1.000 | | 662 | 1.000 | | 1465 | 1.000 | |

| | | | | | | | | | |
|---|---|---|---|---|---|---|---|---|---|
| < 10800 | 1803 | 1.102 (1.045-1.161) | <0.001 | 180 | 1.058 (0.897-1.248) | 0.505 | 413 | 1.102 (0.987-1.230) | 0.083 |
| 10800 - 40760 | 1769 | 1.036 (0.982-1.093) | 0.191 | 180 | 1.007 (0.853-1.190) | 0.930 | 413 | 1.036 (0.928-1.157) | 0.526 |
| 40800 - 525120 | 2770 | 1.139 (1.089-1.192) | <0.001 | 287 | 1.161 (1.008-1.336) | 0.038 | 636 | 1.097 (0.998-1.205) | 0.055 |

Abbreviations: exc., excluded; aHR, adjusted hazard ratio; mon, month; NDMA, N-Nitrosodimethylamine; PSM, propensity score matching.

A shaded area means that each model has the lowest Akaike information criterion.

†All analyses for uterine cancer were performed limited to men.

‡All hazard ratios before PSM were adjusted for age, sex, Charlson's Comorbidity Index, and the year of the first observed valsartan prescription.

§All hazard ratios after PSM were adjusted for age, sex, and the year of the first observed valsartan prescription.

**Supplemental Table 16. The risks of all cancer and organ-specific cancers before and after PSM in study population that received valsartan since 2015.**

| PSM | | Cancer | All cancer | | | Lung cancer | | | Prostate cancer† | | |
|---|---|---|---|---|---|---|---|---|---|---|---|
| | | | Event | aHR (95% CI) | *P* | Event | aHR (95% CI) | *P* | Event | aHR (95% CI) | *P* |
| Before PSM‡ | Overall | NDMA-unexposed | 223140 | 1.000 | | 19307 | 1.000 | | 41017 | 1.000 | |
| | | Eventually NDMA-unexposed | 17497 | 0.920 (0.905-0.935) | <0.001 | 1754 | 0.912 (0.867-0.959) | <0.001 | 3151 | 0.929 (0.895-0.965) | <0.001 |
| | | NDMA-exposed | 25224 | 0.940 (0.928-0.954) | <0.001 | 2505 | 0.908 (0.869-0.949) | <0.001 | 4597 | 0.995 (0.963-1.027) | 0.740 |
| | 6 mon exc. | NDMA-unexposed | 70533 | 1.000 | | 7105 | 1.000 | | 12589 | 1.000 | |
| | | Eventually NDMA-unexposed | 16049 | 1.189 (1.168-1.209) | <0.001 | 1619 | 1.203 (1.139-1.270) | <0.001 | 2898 | 1.195 (1.147-1.245) | <0.001 |
| | | NDMA-exposed | 23507 | 1.170 (1.153-1.188) | <0.001 | 2341 | 1.155 (1.101-1.211) | <0.001 | 4317 | 1.240 (1.197-1.284) | <0.001 |
| | 1-year exc. | NDMA-unexposed | 58608 | 1.000 | | 5881 | 1.000 | | 10443 | 1.000 | |
| | | Eventually NDMA-unexposed | 13265 | 1.098 (1.077-1.119) | <0.001 | 1341 | 1.115 (1.051-1.184) | <0.001 | 2384 | 1.101 (1.053-1.152) | <0.001 |
| | | NDMA-exposed | 20126 | 1.101 (1.083-1.119) | <0.001 | 1980 | 1.075 (1.021-1.132) | 0.006 | 3693 | 1.169 (1.125-1.214) | <0.001 |
| | 2-year exc. | NDMA-unexposed | 38051 | 1.000 | | 3786 | 1.000 | | 6719 | 1.000 | |
| | | Eventually NDMA-unexposed | 8474 | 1.016 (0.992-1.040) | 0.201 | 868 | 1.053 (0.978-1.134) | 0.169 | 1505 | 1.015 (0.960-1.074) | 0.599 |
| | | NDMA-exposed | 13444 | 1.045 (1.025-1.066) | <0.001 | 1314 | 1.021 (0.959-1.088) | 0.511 | 2451 | 1.113 (1.062-1.166) | <0.001 |

| | | | | | | | | | | |
|---|---|---|---|---|---|---|---|---|---|---|
| 1:1 PSM§ | Overall | NDMA-unexposed | 16272 | 1.000 | | 1319 | 1.000 | | 3348 | 1.000 | |
| | | NDMA-exposed | 5876 | 1.003 (0.969-1.039) | 0.865 | 533 | 1.003 (0.893-1.127) | 0.960 | 1187 | 0.983 (0.910-1.062) | 0.667 |
| | 6 mon exc. | NDMA-unexposed | 4343 | 1.000 | | 447 | 1.000 | | 968 | 1.000 | |
| | | NDMA-exposed | 4969 | 1.283 (1.230-1.339) | <0.001 | 487 | 1.273 (1.113-1.457) | <0.001 | 1065 | 1.220 (1.114-1.335) | <0.001 |
| | 1-year exc. | NDMA-unexposed | 3593 | 1.000 | | 372 | 1.000 | | 807 | 1.000 | |
| | | NDMA-exposed | 4221 | 1.183 (1.131-1.238) | <0.001 | 406 | 1.135 (0.982-1.312) | 0.087 | 910 | 1.127 (1.023-1.241) | 0.016 |
| | 2-year exc. | NDMA-unexposed | 2314 | 1.000 | | 226 | 1.000 | | 524 | 1.000 | |
| | | NDMA-exposed | 2800 | 1.123 (1.062-1.187) | <0.001 | 263 | 1.080 (0.902-1.293) | 0.402 | 597 | 1.054 (0.936-1.186) | 0.386 |
| 1:1:1 PSM§ | Overall | NDMA-unexposed | 13976 | 1.000 | | 1062 | 1.000 | | 2886 | 1.000 | |
| | | Eventually NDMA-unexposed | 2959 | 1.014 (0.969-1.060) | 0.560 | 276 | 1.073 (0.924-1.247) | 0.355 | 564 | 0.955 (0.862-1.057) | 0.376 |
| | | NDMA-exposed | 3742 | 1.006 (0.964-1.049) | 0.797 | 328 | 0.994 (0.861-1.149) | 0.940 | 767 | 1.015 (0.924-1.114) | 0.761 |
| | 6 mon exc. | NDMA-unexposed | 2567 | 1.000 | | 281 | 1.000 | | 595 | 1.000 | |
| | | Eventually NDMA-unexposed | 2243 | 1.474 (1.389-1.563) | <0.001 | 251 | 1.559 (1.305-1.861) | <0.001 | 461 | 1.302 (1.148-1.477) | <0.001 |
| | | NDMA-exposed | 2858 | 1.414 (1.337-1.496) | <0.001 | 294 | 1.376 (1.157-1.636) | <0.001 | 664 | 1.390 (1.237-1.562) | <0.001 |
| | 1-year exc. | NDMA-unexposed | 2117 | 1.000 | | 240 | 1.000 | | 498 | 1.000 | |

| | | | | | | | | | | |
|---|---|---|---|---|---|---|---|---|---|---|
| | Eventually NDMA-unexposed | 1777 | 1.235 (1.158-1.317) | <0.001 | 208 | 1.332 (1.101-1.612) | 0.003 | 368 | 1.087 (0.947-1.246) | 0.234 |
| | NDMA-exposed | 2386 | 1.220 (1.148-1.296) | <0.001 | 248 | 1.169 (0.973-1.406) | 0.095 | 558 | 1.203 (1.063-1.362) | 0.004 |
| 2-year exc. | NDMA-unexposed | 1365 | 1.000 | | 137 | 1.000 | | 324 | 1.000 | |
| | Eventually NDMA-unexposed | 1095 | 1.053 (0.972-1.141) | 0.205 | 121 | 1.173 (0.917-1.500) | 0.204 | 221 | 0.890 (0.750-1.057) | 0.185 |
| | NDMA-exposed | 1557 | 1.080 (1.003-1.163) | 0.040 | 160 | 1.112 (0.883-1.402) | 0.366 | 375 | 1.085 (0.933-1.261) | 0.290 |

Abbreviations: aHR, adjusted hazard ratio; CI, confidence interval; exc., excluded; mon, month; NDMA, N-Nitrosodimethylamine; PSM, propensity score matching.

A shaded area means that each model has the lowest Akaike information criterion.

†All analyses for uterine cancer were performed limited to men.

‡All hazard ratios before PSM were adjusted for age, sex, Charlson's Comorbidity Index, being a prevalent user, and the year of the first observed valsartan prescription.

§All hazard ratios after PSM were adjusted for age, sex, being a prevalent user, and the year of the first observed valsartan prescription.

**Supplemental Table 17. The risks of all cancer and organ-specific cancers before and after PSM in valsartan new users observed since 2015.**

| PSM | | Cancer | All cancer | | | Lung cancer | | | Prostate cancer† | | |
|---|---|---|---|---|---|---|---|---|---|---|---|
| | | | Event | aHR (95% CI) | *P* | Event | aHR (95% CI) | *P* | Event | aHR (95% CI) | *P* |
| Before PSM‡ | Overall | NDMA-unexposed | 219151 | 1.000 | | 18955 | 1.000 | | 40243 | 1.000 | |
| | | Eventually NDMA-unexposed | 17497 | 0.919 (0.904-0.934) | <0.001 | 1754 | 0.911 (0.866-0.959) | <0.001 | 3151 | 0.928 (0.894-0.964) | <0.001 |
| | | NDMA-exposed | 25067 | 0.939 (0.926-0.952) | <0.001 | 2491 | 0.908 (0.869-0.949) | <0.001 | 4574 | 0.994 (0.963-1.027) | 0.736 |
| | 6 mon exc. | NDMA-unexposed | 68851 | 1.000 | | 6932 | 1.000 | | 12276 | 1.000 | |
| | | Eventually NDMA-unexposed | 16049 | 1.188 (1.167-1.209) | <0.001 | 1619 | 1.202 (1.138-1.269) | <0.001 | 2896 | 1.195 (1.148-1.245) | <0.001 |
| | | NDMA-exposed | 23360 | 1.170 (1.153-1.188) | <0.001 | 2328 | 1.156 (1.102-1.212) | <0.001 | 4296 | 1.242 (1.199-1.287) | <0.001 |
| | 1-year exc. | NDMA-unexposed | 57189 | 1.000 | | 5736 | 1.000 | | 10175 | 1.000 | |
| | | Eventually NDMA-unexposed | 13265 | 1.097 (1.077-1.118) | <0.001 | 1341 | 1.114 (1.050-1.183) | <0.001 | 2384 | 1.102 (1.053-1.152) | <0.001 |
| | | NDMA-exposed | 20000 | 1.100 (1.082-1.118) | <0.001 | 1969 | 1.075 (1.021-1.132) | 0.006 | 3673 | 1.170 (1.126-1.215) | <0.001 |
| | 2-year exc. | NDMA-unexposed | 37103 | 1.000 | | 3695 | 1.000 | | 6537 | 1.000 | |
| | | Eventually NDMA-unexposed | 8474 | 1.015 (0.992-1.040) | 0.206 | 868 | 1.053 (0.978-1.134) | 0.168 | 1505 | 1.016 (0.960-1.074) | 0.586 |
| | | NDMA-exposed | 13356 | 1.045 (1.025-1.066) | <0.001 | 1307 | 1.023 (0.960-1.090) | 0.490 | 2438 | 1.115 (1.064-1.168) | <0.001 |

| | | | | | | | | | | |
|---|---|---|---|---|---|---|---|---|---|---|
| 1:1 PSM§ | Overall | NDMA-unexposed | 16265 | 1.000 | | 1319 | 1.000 | | 3348 | 1.000 | |
| | | NDMA-exposed | 5873 | 1.003 (0.969-1.039) | 0.868 | 532 | 1.002 (0.892-1.125) | 0.979 | 1187 | 0.983 (0.910-1.062) | 0.667 |
| | 6 mon exc. | NDMA-unexposed | 4342 | 1.000 | | 447 | 1.000 | | 968 | 1.000 | |
| | | NDMA-exposed | 4966 | 1.282 (1.229-1.338) | <0.001 | 486 | 1.271 (1.111-1.455) | 0.001 | 1065 | 1.220 (1.114-1.335) | <0.001 |
| | 1-year exc. | NDMA-unexposed | 3593 | 1.000 | | 372 | 1.000 | | 807 | 1.000 | |
| | | NDMA-exposed | 4219 | 1.183 (1.130-1.238) | <0.001 | 405 | 1.133 (0.980-1.310) | 0.091 | 910 | 1.127 (1.023-1.241) | 0.016 |
| | 2-year exc. | NDMA-unexposed | 2314 | 1.000 | | 226 | 1.000 | | 524 | 1.000 | |
| | | NDMA-exposed | 2806 | 1.123 (1.062-1.187) | <0.001 | 262 | 1.077 (0.899-1.290) | 0.420 | 597 | 1.054 (0.936-1.186) | 0.386 |
| 1:1:1 PSM§ | Overall | NDMA-unexposed | 13975 | 1.000 | | 1062 | 1.000 | | 2886 | 1.000 | |
| | | Eventually NDMA-unexposed | 2959 | 1.014 (0.969-1.060) | 0.556 | 276 | 1.073 (0.924-1.247) | 0.355 | 564 | 0.955 (0.862-1.057) | 0.376 |
| | | NDMA-exposed | 3742 | 1.006 (0.964-1.049) | 0.793 | 328 | 0.994 (0.861-1.149) | 0.940 | 767 | 1.015 (0.924-1.114) | 0.761 |
| | 6 mon exc. | NDMA-unexposed | 2567 | 1.000 | | 281 | 1.000 | | 595 | 1.000 | |
| | | Eventually NDMA-unexposed | 2243 | 1.474 (1.389-1.563) | <0.001 | 251 | 1.559 (1.305-1.861) | <0.001 | 461 | 1.302 (1.148-1.477) | <0.001 |
| | | NDMA-exposed | 2858 | 1.414 (1.337-1.496) | <0.001 | 294 | 1.376 (1.157-1.636) | <0.001 | 664 | 1.390 (1.237-1.562) | <0.001 |
| | 1-year exc. | NDMA-unexposed | 2117 | 1.000 | | 240 | 1.000 | | 498 | 1.000 | |

| | | | | | | | | | | |
|---|---|---|---|---|---|---|---|---|---|---|
| | Eventually NDMA-unexposed | 1777 | 1.235 (1.158-1.317) | <0.001 | 208 | 1.332 (1.101-1.612) | 0.003 | 368 | 1.087 (0.947-1.246) | 0.234 |
| | NDMA-exposed | 2386 | 1.220 (1.148-1.296) | <0.001 | 248 | 1.169 (0.973-1.406) | 0.095 | 558 | 1.203 (1.063-1.362) | 0.004 |
| 2-year exc. | NDMA-unexposed | 1365 | 1.000 | | 137 | 1.000 | | 324 | 1.000 | |
| | Eventually NDMA-unexposed | 1095 | 1.053 (0.972-1.141) | 0.205 | 121 | 1.173 (0.917-1.500) | 0.204 | 221 | 0.890 (0.750-1.057) | 0.185 |
| | NDMA-exposed | 1557 | 1.080 (1.003-1.163) | 0.040 | 160 | 1.112 (0.883-1.402) | 0.366 | 375 | 1.085 (0.933-1.261) | 0.290 |

Abbreviations: aHR, adjusted hazard ratio; CI, confidence interval; exc., excluded; mon, month; NDMA, N-Nitrosodimethylamine; PSM, propensity score matching.

A shaded area means that each model has the lowest Akaike information criterion.

†All analyses for uterine cancer were performed limited to men.

‡All hazard ratios before PSM were adjusted for age, sex, Charlson's Comorbidity Index, and the year of the first observed valsartan prescription.

§All hazard ratios after PSM were adjusted for age, sex, and the year of the first observed valsartan prescription.

**Supplemental Table 18. The risks of all cancer and organ-specific cancers before and after PSM in study population that received valsartan since 2015.**

| | | | All cancer | | | Lung cancer | | | Prostate cancer† | | |
|---|---|---|---|---|---|---|---|---|---|---|---|
| **PSM** | | | **Event** | **aHR (95% CI)** | ***P*** | **Event** | **aHR (95% CI)** | ***P*** | **Event** | **aHR (95% CI)** | ***P*** |
| Before PSM‡ | Overall | NDMA-unexposed | 199499 | 1.000 | | 17498 | 1.000 | | 36607 | 1.000 | |
| | | < 10800 | 11101 | 1.009 (0.990-1.029) | 0.366 | 1124 | 1.014 (0.954-1.078) | 0.658 | 1935 | 1.021 (0.974-1.070) | 0.385 |
| | | 10800 - 40760 | 9704 | 0.942 (0.923-0.962) | <0.001 | 988 | 0.943 (0.883-1.007) | 0.078 | 1777 | 1.000 (0.953-1.050) | 0.988 |
| | | 40800 - 525120 | 6629 | 0.895 (0.872-0.917) | <0.001 | 625 | 0.808 (0.745-0.877) | <0.001 | 1242 | 0.973 (0.918-1.031) | 0.357 |
| | 6 mon exc. | NDMA-unexposed | 70533 | 1.000 | | 7105 | 1.000 | | 12589 | 1.000 | |
| | | < 10800 | 7982 | 1.102 (1.077-1.128) | <0.001 | 805 | 1.092 (1.015-1.175) | 0.018 | 1425 | 1.143 (1.082-1.208) | <0.001 |
| | | 10800 - 40760 | 8898 | 1.219 (1.192-1.247) | <0.001 | 912 | 1.239 (1.156-1.329) | <0.001 | 1652 | 1.307 (1.241-1.377) | <0.001 |
| | | 40800 - 525120 | 6627 | 0.944 (0.920-0.969) | <0.001 | 624 | 0.891 (0.820-0.968) | 0.007 | 1240 | 1.007 (0.949-1.068) | 0.825 |
| | 1-year exc. | NDMA-unexposed | 58608 | 1.000 | | 5881 | 1.000 | | 10443 | 1.000 | |
| | | < 10800 | 6614 | 1.056 (1.029-1.083) | <0.001 | 667 | 1.049 (0.968-1.136) | 0.247 | 1169 | 1.090 (1.026-1.158) | 0.005 |
| | | 10800 - 40760 | 7058 | 1.107 (1.079-1.135) | <0.001 | 705 | 1.095 (1.012-1.185) | 0.023 | 1325 | 1.204 (1.136-1.275) | <0.001 |
| | | 40800 - 525120 | 6454 | 0.986 (0.960-1.012) | 0.273 | 608 | 0.932 (0.856 -1.014) | 0.103 | 1199 | 1.045 (0.983-1.110) | 0.158 |
| | 2-year exc. | NDMA-unexposed | 38051 | 1.000 | | 3786 | 1.000 | | 6719 | 1.000 | |
| | | < 10800 | 4144 | 1.084 (1.050-1.119) | <0.001 | 408 | 1.059 (0.956-1.174) | 0.269 | 715 | 1.105 (1.023-1.194) | 0.011 |

| | | | | | | | | | | | |
|---|---|---|---|---|---|---|---|---|---|---|---|
| | | 10800 - 40760 | 4173 | 1.018 (0.985-1.051) | 0.292 | 422 | 1.025 (0.926-1.135) | 0.634 | 781 | 1.117 (1.036-1.204) | 0.004 |
| | | 40800 - 525120 | 5127 | 0.997 (0.968-1.027) | 0.848 | 484 | 0.949 (0.863-1.044) | 0.283 | 955 | 1.073 (1.002-1.149) | 0.043 |
| 1:1 PSM§ | Overall | NDMA-unexposed | 15799 | 1.000 | | 1268 | 1.000 | | 3275 | 1.000 | |
| | | < 10800 | 2484 | 1.072 (1.025-1.122) | 0.003 | 230 | 1.110 (0.956-1.289) | 0.170 | 478 | 0.995 (0.898-1.101) | 0.917 |
| | | 10800 - 40760 | 2276 | 1.017 (0.971-1.067) | 0.471 | 219 | 1.091 (0.935-1.272) | 0.269 | 445 | 0.955 (0.859-1.061) | 0.391 |
| | | 40800 - 525120 | 1589 | 0.978 (0.924-1.034) | 0.430 | 135 | 0.941 (0.776-1.141) | 0.538 | 337 | 0.984 (0.870-1.112) | 0.793 |
| | 6 mon exc. | NDMA-unexposed | 4343 | 1.000 | | 447 | 1.000 | | 968 | 1.000 | |
| | | < 10800 | 1635 | 1.211 (1.143-1.283) | <0.001 | 161 | 1.159 (0.966-1.392) | 0.113 | 344 | 1.148 (1.014-1.300) | 0.029 |
| | | 10800 - 40760 | 1885 | 1.358 (1.285-1.435) | <0.001 | 196 | 1.385 (1.166-1.646) | <0.001 | 401 | 1.270 (1.128-1.430) | <0.001 |
| | | 40800 - 525120 | 1449 | 1.060 (0.997-1.128) | 0.064 | 130 | 1.014 (0.826-1.244) | 0.895 | 320 | 1.033 (0.906-1.179) | 0.625 |
| | 1-year exc. | NDMA-unexposed | 3593 | 1.000 | | 372 | 1.000 | | 807 | 1.000 | |
| | | < 10800 | 1353 | 1.137 (1.068-1.211) | <0.001 | 133 | 1.073 (0.878-1.310) | 0.492 | 272 | 1.031 (0.898-1.184) | 0.664 |
| | | 10800 - 40760 | 1467 | 1.191 (1.120-1.267) | <0.001 | 148 | 1.157 (0.953-1.405) | 0.141 | 326 | 1.157 (1.016-1.319) | 0.028 |
| | | 40800 - 525120 | 1401 | 1.082 (1.015-1.153) | 0.015 | 125 | 1.015 (0.822-1.252) | 0.892 | 312 | 1.058 (0.925-1.210) | 0.410 |
| | 2-year exc. | NDMA-unexposed | 2314 | 1.000 | | 226 | 1.000 | | 524 | 1.000 | |
| | | < 10800 | 844 | 1.160 (1.072-1.255) | <0.001 | 71 | 0.973 (0.745-1.272) | 0.843 | 164 | 1.037 (0.871-1.235) | 0.682 |

| | | | | | | | | | | | |
|---|---|---|---|---|---|---|---|---|---|---|---|
| | | 10800 - 40760 | 862 | 1.085 (1.002-1.175) | 0.045 | 92 | 1.124 (0.877-1.440) | 0.356 | 186 | 1.031 (0.869-1.222) | 0.730 |
| | | 40800 - 525120 | 1102 | 1.087 (1.010-1.169) | 0.025 | 100 | 1.079 (0.849-1.372) | 0.536 | 244 | 1.051 (0.901-1.226) | 0.529 |
| 1:1:1 PSM§ | Overall | NDMA-unexposed | 12670 | 1.000 | | 1003 | 1.000 | | 2707 | 1.000 | |
| | | < 10800 | 2019 | 1.084 (1.031-1.140) | 0.002 | 191 | 1.162 (0.986-1.370) | 0.074 | 393 | 0.982 (0.878-1.099) | 0.754 |
| | | 10800 - 40760 | 1870 | 1.037 (0.984-1.092) | 0.173 | 181 | 1.137 (0.959-1.347) | 0.139 | 381 | 0.974 (0.868-1.092) | 0.649 |
| | | 40800 - 525120 | 1307 | 0.991 (0.931-1.054) | 0.769 | 108 | 0.939 (0.757-1.165) | 0.567 | 284 | 0.968 (0.847-1.106) | 0.634 |
| | 6 mon exc. | NDMA-unexposed | 3532 | 1.000 | | 358 | 1.000 | | 815 | 1.000 | |
| | | < 10800 | 1343 | 1.228 (1.152-1.308) | <0.001 | 131 | 1.165 (0.951-1.427) | 0.140 | 290 | 1.157 (1.010-1.324) | 0.035 |
| | | 10800 - 40760 | 1535 | 1.364 (1.282-1.450) | <0.001 | 159 | 1.383 (1.142-1.675) | 0.001 | 341 | 1.272 (1.118-1.447) | <0.001 |
| | | 40800 - 525120 | 1195 | 1.068 (0.997-1.144) | 0.060 | 105 | 0.995 (0.793-1.249) | 0.969 | 268 | 0.996 (0.863-1.149) | 0.955 |
| | 1-year exc. | NDMA-unexposed | 2932 | 1.000 | | 300 | 1.000 | | 684 | 1.000 | |
| | | < 10800 | 1113 | 1.149 (1.072-1.232) | <0.001 | 110 | 1.099 (0.882-1.370) | 0.400 | 231 | 1.039 (0.894-1.207) | 0.616 |
| | | 10800 - 40760 | 1191 | 1.188 (1.109-1.272) | <0.001 | 119 | 1.152 (0.928-1.430) | 0.198 | 275 | 1.143 (0.991-1.317) | 0.066 |
| | | 40800 - 525120 | 1157 | 1.088 (1.014-1.167) | 0.018 | 104 | 1.031 (0.818-1.299) | 0.797 | 261 | 1.015 (0.877-1.175) | 0.842 |
| | 2-year exc. | NDMA-unexposed | 1887 | 1.000 | | 181 | 1.000 | | 446 | 1.000 | |
| | | < 10800 | 693 | 1.175 (1.077-1.282) | <0.001 | 61 | 1.059 (0.792-1.417) | 0.700 | 143 | 1.049 (0.868-1.267) | 0.621 |

| | | | | | | | | | |
|---|---|---|---|---|---|---|---|---|---|
| 10800 - 40760 | 696 | 1.079 (0.987-1.179) | 0.093 | 72 | 1.125 (0.851-1.487) | 0.410 | 162 | 1.049 (0.873-1.260) | 0.609 |
| 40800 - 525120 | 905 | 1.086 (1.002-1.177) | 0.045 | 82 | 1.110 (0.850-1.449) | 0.443 | 204 | 1.008 (0.852-1.193) | 0.924 |

Abbreviations: exc., excluded; aHR, adjusted hazard ratio; mon, month; NDMA, N-Nitrosodimethylamine; PSM, propensity score matching.

A shaded area means that each model has the lowest Akaike information criterion.

†All analyses for uterine cancer were performed limited to men.

‡All hazard ratios before PSM were adjusted for age, sex, Charlson's Comorbidity Index, being a prevalent user, and the year of the first observed valsartan prescription.

§All hazard ratios after PSM were adjusted for age, sex, being a prevalent user, and the year of the first observed valsartan prescription.

**Supplemental Table 19. The risks of all cancer and organ-specific cancers before and after PSM in valsartan new users observed since 2015.**

| | | | All cancer | | | Lung cancer | | | Prostate cancer† | | |
|---|---|---|---|---|---|---|---|---|---|---|---|
| PSM | | | Event | aHR (95% CI) | *P* | Event | aHR (95% CI) | *P* | Event | aHR (95% CI) | *P* |
| Before PSM‡ | Overall | NDMA-unexposed | 19552 2 | 1.000 | | 17147 | 1.000 | | 35836 | 1.000 | |
| | | < 10800 | 11039 | 1.009 (0.989-1.029) | 0.379 | 1119 | 1.015 (0.955-1.080) | 0.625 | 1923 | 1.020 (0.973-1.069) | 0.403 |
| | | 10800 - 40760 | 9639 | 0.941 (0.921-0.961) | <0.001 | 981 | 0.941 (0.881-1.005) | 0.070 | 1768 | 1.001 (0.953-1.051) | 0.982 |
| | | 40800 - 525120 | 6588 | 0.893 (0.871-0.916) | <0.001 | 622 | 0.809 (0.745-0.878) | <0.001 | 1237 | 0.973 (0.918-1.032) | 0.358 |
| | 6 mon exc. | NDMA-unexposed | 68851 | 1.000 | | 6932 | 1.000 | | 12276 | 1.000 | |
| | | < 10800 | 7934 | 1.102 (1.077-1.128) | <0.001 | 801 | 1.094 (1.016-1.177) | 0.017 | 1418 | 1.145 (1.083-1.210) | <0.001 |
| | | 10800 - 40760 | 8840 | 1.218 (1.192-1.246) | <0.001 | 906 | 1.238 (1.155-1.328) | <0.001 | 1643 | 1.309 (1.243-1.379) | <0.001 |
| | | 40800 - 525120 | 6586 | 0.944 (0.920-0.968) | <0.001 | 621 | 0.892 (0.820-0.969) | 0.007 | 1235 | 1.009 (0.951-1.071) | 0.766 |
| | 1-year exc. | NDMA-unexposed | 57189 | 1.000 | | 5736 | 1.000 | | 10175 | 1.000 | |
| | | < 10800 | 6575 | 1.056 (1.029-1.083) | <0.001 | 664 | 1.050 (0.969-1.138) | 0.236 | 1162 | 1.091 (1.026-1.159) | 0.005 |
| | | 10800 - 40760 | 7012 | 1.106 (1.079-1.134) | <0.001 | 700 | 1.094 (1.011-1.184) | 0.027 | 1317 | 1.204 (1.137-1.276) | <0.001 |
| | | 40800 - 525120 | 6413 | 0.985 (0.959-1.011) | 0.247 | 605 | 0.933 (0.857-1.015) | 0.108 | 1194 | 1.047 (0.985-1.112) | 0.141 |
| | 2-year exc. | NDMA-unexposed | 37103 | 1.000 | | 3695 | 1.000 | | 6537 | 1.000 | |
| | | < 10800 | 4120 | 1.085 (1.050-1.120) | <0.001 | 407 | 1.064 (0.960-1.179) | 0.237 | 712 | 1.108 (1.026-1.198) | 0.009 |

| | | | | | | | | | | |
|---|---|---|---|---|---|---|---|---|---|---|
| | | 10800 - 40760 | 4146 | 1.017 (0.985-1.051) | 0.296 | 419 | 1.024 (0.925-1.134) | 0.645 | 775 | 1.116 (1.036-1.204) | 0.004 |
| | | 40800 - 525120 | 5090 | 0.996 (0.967-1.026) | 0.784 | 481 | 0.949 (0.863-1.045) | 0.287 | 951 | 1.076 (1.005-1.152) | 0.036 |
| 1:1 PSM§ | Overall | NDMA-unexposed | 15792 | 1.000 | | 1268 | 1.000 | | 3275 | 1.000 | |
| | | < 10800 | 2484 | 1.073 (1.025-1.122) | 0.002 | 230 | 1.110 (0.956-1.289) | 0.170 | 478 | 0.995 (0.898-1.101) | 0.920 |
| | | 10800 - 40760 | 2274 | 1.017 (0.970-1.066) | 0.481 | 219 | 1.091 (0.936-1.273) | 0.267 | 445 | 0.955 (0.859-1.061) | 0.392 |
| | | 40800 - 525120 | 1588 | 0.977 (0.924-1.034) | 0.427 | 134 | 0.935 (0.770-1.134) | 0.492 | 337 | 0.984 (0.871-1.112) | 0.798 |
| | 6 mon exc. | NDMA-unexposed | 4342 | 1.000 | | 447 | 1.000 | | 968 | 1.000 | |
| | | < 10800 | 1635 | 1.211 (1.143-1.283) | <0.001 | 161 | 1.160 (0.966-1.392) | 0.112 | 344 | 1.148 (1.014-1.300) | 0.029 |
| | | 10800 - 40760 | 1883 | 1.357 (1.284-1.434) | <0.001 | 196 | 1.386 (1.167-1.647) | <0.001 | 401 | 1.270 (1.128-1.430) | <0.001 |
| | | 40800 - 525120 | 1448 | 1.060 (0.996-1.128) | 0.066 | 129 | 1.007 (0.820-1.236) | 0.949 | 320 | 1.033 (0.906-1.179) | 0.625 |
| | 1-year exc. | NDMA-unexposed | 3593 | 1.000 | | 372 | 1.000 | | 807 | 1.000 | |
| | | < 10800 | 1353 | 1.137 (1.068-1.211) | <0.001 | 133 | 1.073 (0.878-1.310) | 0.490 | 272 | 1.031 (0.898-1.184) | 0.664 |
| | | 10800 - 40760 | 1466 | 1.191 (1.119-1.266) | <0.001 | 148 | 1.158 (0.954-1.407) | 0.138 | 326 | 1.157 (1.016-1.319) | 0.028 |
| | | 40800 - 525120 | 1400 | 1.081 (1.015-1.153) | 0.016 | 124 | 1.007 (0.816-1.243) | 0.947 | 312 | 1.058 (0.925-1.210) | 0.410 |
| | 2-year exc. | NDMA-unexposed | 2314 | 1.000 | | 226 | 1.000 | | 524 | 1.000 | |
| | | < 10800 | 844 | 1.160 (1.072-1.255) | <0.001 | 71 | 0.974 (0.745-1.272) | 0.845 | 167 | 1.037 (0.871-1.235) | 0.682 |

| | | | | | | | | | | | |
|---|---|---|---|---|---|---|---|---|---|---|---|
| | | 10800 - 40760 | 861 | 1.084 (1.001-1.174) | 0.048 | 92 | 1.126 (0.879-1.442) | 0.349 | 186 | 1.030 (0.869-1.222) | 0.730 |
| | | 40800 - 525120 | 1101 | 1.086 (1.009-1.168) | 0.027 | 99 | 1.069 (0.840-1.360) | 0.586 | 244 | 1.051 (0.901-1.226) | 0.529 |
| 1:1:1 PSM§ | Overall | NDMA-unexposed | 12670 | 1.000 | | 1003 | 1.000 | | 2707 | 1.000 | |
| | | < 10800 | 2019 | 1.084 (1.031-1.140) | 0.002 | 191 | 1.162 (0.986-1.370) | 0.074 | 393 | 0.982 (0.878-1.099) | 0.754 |
| | | 10800 - 40760 | 1870 | 1.037 (0.984-1.092) | 0.173 | 181 | 1.137 (0.959-1.347) | 0.139 | 381 | 0.974 (0.868-1.092) | 0.649 |
| | | 40800 - 525120 | 1307 | 0.991 (0.931-1.054) | 0.769 | 108 | 0.939 (0.757-1.165) | 0.569 | 284 | 0.968 (0.847-1.106) | 0.634 |
| | 6 mon exc. | NDMA-unexposed | 3532 | 1.000 | | 358 | 1.000 | | 815 | 1.000 | |
| | | < 10800 | 1343 | 1.228 (1.152-1.308) | <0.001 | 131 | 1.165 (0.951-1.427) | 0.140 | 290 | 1.157 (1.010-1.324) | 0.035 |
| | | 10800 - 40760 | 1535 | 1.364 (1.282-1.450) | <0.001 | 159 | 1.383 (1.142-1.675) | 0.001 | 341 | 1.272 (1.118-1.447) | <0.001 |
| | | 40800 - 525120 | 1195 | 1.068 (0.997-1.144) | 0.060 | 105 | 0.995 (0.793-1.249) | 0.969 | 268 | 0.996 (0.863-1.149) | 0.955 |
| | 1-year exc. | NDMA-unexposed | 2932 | 1.000 | | 300 | 1.000 | | 684 | 1.000 | |
| | | < 10800 | 1113 | 1.149 (1.072-1.232) | <0.001 | 110 | 1.099 (0.882-1.370) | 0.400 | 231 | 1.039 (0.894-1.207) | 0 616 |
| | | 10800 - 40760 | 1191 | 1.188 (1.109-1.272) | <0.001 | 119 | 1.152 (0.928-1.430) | 0.198 | 275 | 1.143 (0.991-1.317) | 0.066 |
| | | 40800 - 525120 | 1157 | 1.088 (1.014-1.167) | 0.018 | 104 | 1.031 (0.818-1.299) | 0.797 | 261 | 1.015 (0.877-1.175) | 0.842 |
| | 2-year exc. | NDMA-unexposed | 1887 | 1.000 | | 181 | 1.000 | | 446 | 1.000 | |
| | | < 10800 | 693 | 1.175 (1.076-1.282) | <0.001 | 61 | 1.059 (0.792-1.417) | 0.700 | 143 | 1.049 (0.868-1.267) | 0.621 |

| | | | | | | | | | |
|---|---|---|---|---|---|---|---|---|---|
| 10800 - 40760 | 696 | 1.079 (0.987-1.179) | 0.093 | 72 | 1.125 (0.851-1.487) | 0.410 | 162 | 1.049 (0.873-1.260) | 0.609 |
| 40800 - 525120 | 905 | 1.086 (1.002-1.177) | 0.045 | 82 | 1.110 (0.850-1.449) | 0.443 | 204 | 1.008 (0.852-1.193) | 0.924 |

Abbreviations: exc., excluded; aHR, adjusted hazard ratio; mon, month; NDMA, N-Nitrosodimethylamine; PSM, propensity score matching.

A shaded area means that each model has the lowest Akaike information criterion.

[†]All analyses for uterine cancer were performed limited to men.

[‡]All hazard ratios before PSM were adjusted for age, sex, Charlson's Comorbidity Index, and the year of the first observed valsartan prescription.

[§]All hazard ratios after PSM were adjusted for age, sex, and the year of the first observed valsartan prescription.

**Supplemental Table 20. The number of all-cause and cardiovascular death in the whole population in 2018.**

| Death | PSM | From January 1, 2018 to July 6, 2018 | | | From July 7, 2018 to December 31, 2018 | | |
|---|---|---|---|---|---|---|---|
| | | Original | Eventually NDMA-uncontaminated | NDMA-contaminated | Original | Eventually NDMA-uncontaminated | NDMA-contaminated |
| **All-cause** | Event | 18,963 | 2,134 | 4,317 | 17,942 | 2,208 | 4,059 |
| | IR (95% CI)† | 17.48 (17.24-17.73) | 11.29 (10.82-11.78) | 18.47 (17.92-19.03) | 17.38 (17.12-17.63) | 12.27 (11.77-12.79) | 18.24 (17.68-18.81) |
| **Cardiovascular** | Event | 5,056 | 519 | 1,062 | 4,596 | 497 | 1,012 |
| | IR (95% CI)† | 4.66 (4.53-4.79) | 2.75 (2.52-2.99) | 4.54 (4.27-4.83) | 4.45 (4.32-4.58) | 2.76 (2.53-3.02) | 4.55 (4.27-4.84) |

Abbreviations: CI, confidence interval; IR, incidence rate; NDMA, N-Nitrosodimethylamine; PSM, propensity score matching.

†Incidence rate per 1,000 person-years is calculated for each valsartan group and clinical outcome.
